# Genetic drivers of hippocampal atrophy highlight the role of *APOE* functional variants and AD polygenicity in Mild Cognitive Impairment

**DOI:** 10.1101/2025.03.25.25324619

**Authors:** Natalia Vilor-Tejedor, Albert Rodrigo, Patricia Genius, Blanca Rodríguez-Fernández, Federica Anastasi, Wiesje Pelkmans, Arcadi Navarro, Hieab H. Adams, Laura Wisse, Juan D. Gispert, Tavia E. Evans, the ADNI

## Abstract

The hippocampus, crucial in cognitive aging and Alzheimer’s disease (AD), shows early atrophy during disease progression. This study investigates 5-year trajectories of hippocampal volumes across AD stages, focusing on genetic predisposition to AD and their impact on hippocampal atrophy. Analyzing data from 1,051 participants in the ADNI, we found that higher genetic predisposition to AD accelerates hippocampal atrophy, particularly in individuals with mild cognitive impairment (MCI). Effects were primarily driven by cerebrospinal fluid protein quantitative trait loci of APOE. Excluding the *APOE* region from analyses negated these effects, underscoring its critical role. This stage-specific effect suggests that AD genetic factors, particularly the *APOE* region, exert their influence primarily before or during the transition to MCI, highlighting the hippocampus’s increased vulnerability during this period. These findings underscore the importance of targeting the MCI stage for early detection and intervention in AD. Further research is warranted to elucidate these dynamics and their potential clinical applications.

## INTRODUCTION

The hippocampus plays a crucial role in learning and memory, and abnormalities are observed in many neurological and psychiatric disorders, including Alzheimer’s disease (AD). Hippocampal atrophy, a hallmark of AD, is highly sensitive to neurodegeneration and can be observed years before the onset of clinical symptoms (Brickman et al., 2011; Evans et al., 2018)even in the preclinical stages (Bartsch & Wulff, 2015; Yang & Yu, 2017). This early vulnerability highlights the hippocampus as one of the first brain regions affected by AD, making it a crucial target for early detection and intervention.

Studies exploring genetic influences of hippocampal atrophy offer a unique window into the biological pathways that may predispose individuals to AD before clinical symptoms emerge. By identifying specific genetic factors associated with hippocampal structure, these studies deepen our understanding of the mechanisms that drive early vulnerability to AD, potentially guiding future strategies for prevention and intervention. However, genetic bases of hippocampal atrophy remain incompletely understood. Few studies have shown associations between genetic factors and hippocampal volume across various neurological pathologies, establishing the hippocampus as a primary area of interest for these conditions (Cai et al., 2024; Vilor-Tejedor et al., 2021; Xu et al., 2020). The ε4 allele of *APOE*, a well-known genetic risk factor for AD, has been implicated in both AD disease development and hippocampal structure changes. Carriers of this allele have a higher risk of developing AD, and the allele’s impact on hippocampal volume may provide insights into potential mechanistic pathways by which it exerts its influence. Studies have pointed out the associations between the *APOE*-ε4 allele and AD polygenic risk score with hippocampal volume in cognitively healthy individuals (Foo et al., 2021; Walhovd et al., 2020), although others suggested an effect only driven by *APOE*-ε4 carriers (Håglin et al., 2023).

Alongside traditional *APOE* alleles, the investigation of *APOE*-related quantitative trait loci (QTLs), has gained prominence in recent genetic studies as a means to better understand genetic predisposition to AD and its structural impact on the hippocampus. Previous studies demonstrated that these *APOE* QTLs can modulate Apolipoprotein E (ApoE) protein expression differently than classical *APOE* alleles (ε2,ε3,ε4) and have been associated with variations in ApoE plasma levels, potentially impacting the brain’s vulnerability to neurodegeneration and introducing another layer of genetic complexity (Liu et al., 2013; Raulin et al., 2022).

In addition to the *APOE-*ε4 allele, the broader genetic predisposition to AD has gained increasing attention in recent genetic studies. By identifying specific genetic factors associated with hippocampal structure, these studies offer a unique window into the biological pathways that may predispose individuals to AD before clinical symptoms emerge. This connection underscores the complex interplay between genetics and neurodegeneration, suggesting that the risk of developing AD might be linked to genetic factors that influence hippocampal integrity.

Sex-specific effects on hippocampal atrophy have also gained attention, as accumulating evidence suggests that men and women may experience different patterns of hippocampal decline, particularly in the context of AD (Yagi & Galea, 2018). For instance, women with the *APOE*-ε4 allele have shown a heightened risk of AD and may experience faster hippocampal atrophy compared to men with the same genetic profile (Sundermann et al., 2018). Investigating these sex-specific genetic effects is crucial, as it may help clarify the biological mechanisms driving differential vulnerability to AD and could lead to more personalized approaches in prevention, early detection, and treatment strategies.

The majority of studies on the genetic influence on hippocampal structure have been cross-sectional, yield non-significant findings at a single time point (Vilor-Tejedor et al., 2021). The only longitudinal study examining the effects of *APOE* (over 5 time points spanning 1.5 years) revealed that ε4 carriers exhibited a greater rate of volume loss in the right hippocampal subfields compared to non-ε4 carriers (Reiter et al., 2017). However, a significant gap remains in understanding how these genetic risks interact with disease groups and sex, to influence hippocampal atrophy over time. To address this, the objectives of the current study were (i) to investigate whether hippocampal trajectories over a 5-year period exhibit variations across different AD groups (CU amyloid-beta (A)-/A+, MCI A+, AD A+) (ii) to evaluate whether there is a distinct sex-related impact on the rate of hippocampal volume decline, and (iii) to determine whether genetic predisposition to AD and *APOE* functional genetic variants contributes to the rate of hippocampal volume atrophy. Exploratory analyses into hippocampal subfields were performed.

## METHODS

### Participants

This study included participants from the Alzheimer’s Disease Neuroimaging Initiative (ADNI) cohort, consisting of CU, A+ MCI and individuals clinically diagnosed with AD dementia (A+), for whom complete information on demographic, genetic, and MRI scans and at least two longitudinal visits was available. Clinical assessment closest in time to Aβ PET imaging was used to define diagnostic groups. Further details are available elsewhere (Petersen et al., 2010). Of the 2,146 ADNI participants with demographic information, 246 were excluded due to a lack of genetic information, and 158 were excluded due to discrepancies between reported amyloid status. Among the remaining participants, 615 individuals were excluded due to incomplete MRI sequences. Additionally, 32 participants were excluded due to changes in scanner parameters during their follow-up for brain imaging acquisition. Finally, individuals with less than two visits were excluded from the analysis, as well as 2 individuals presenting technical errors in the segmentation. The final sample size included in this study comprised 1,051 participants (CU=363; A+ positive MCI=474, A+ AD patients, AD=214) [**Figure 1**].

**Figure 1.**
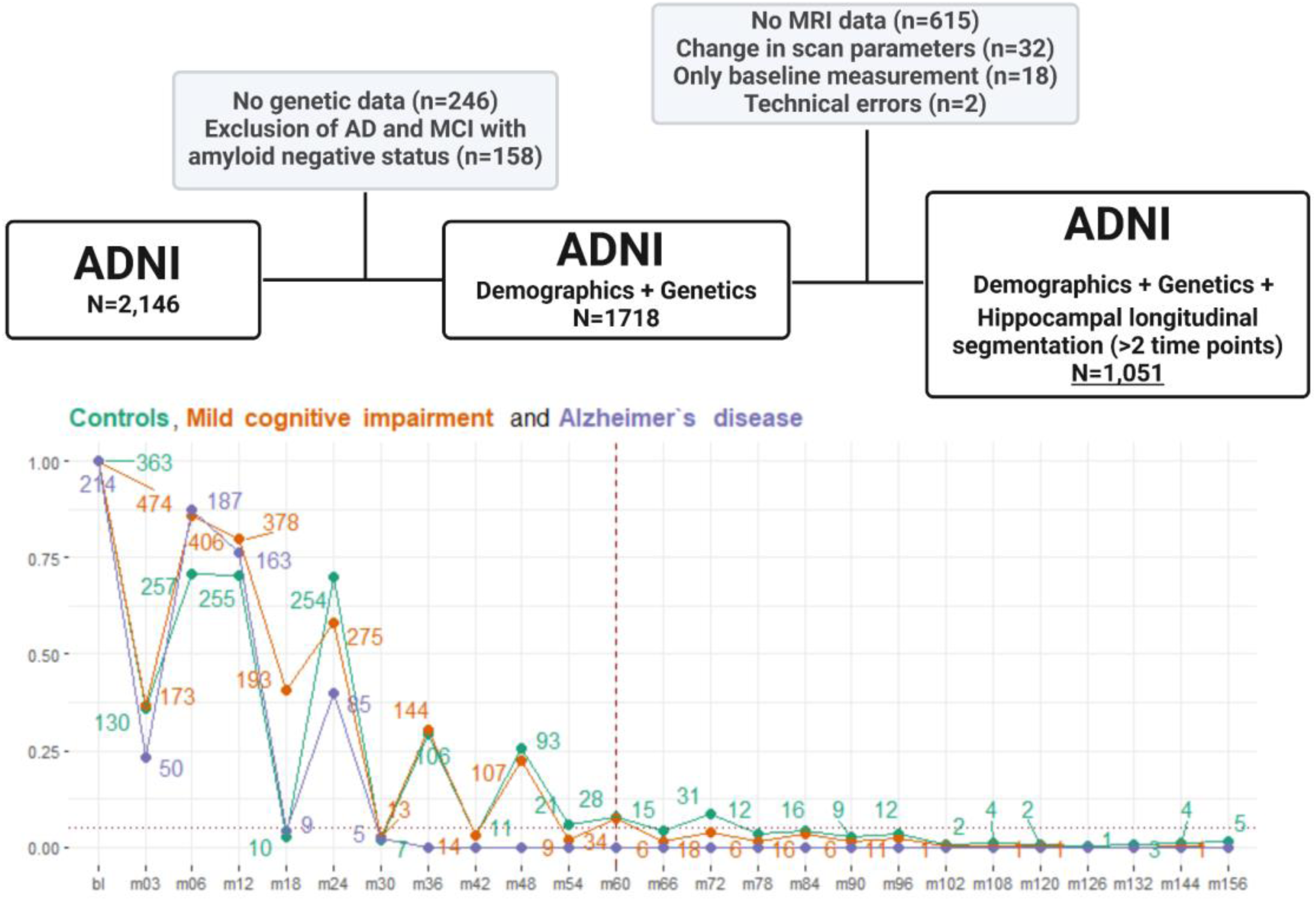
Flowchart study.

### Magnetic resonance imaging acquisition

The data consisted of standard T1-weighted images obtained through volumetric 3D sagittal MPRAGE or equivalent protocols with varying resolution. The typical protocol parameters included a repetition time (TR) of 2,400 ms, inversion time (TI) of 1,000 ms, flip angle of 8 degrees, and a field of view (FOV) of 24 cm. The acquisition matrix was set to 256 × 256 × 170 in the x-, y-, and z-dimensions, resulting in a voxel size of 1.25 × 1.26 × 1.2 mm. The MRI data underwent preprocessing steps, including correction for anterior commissure and posterior commissure, skull stripping, cerebellum removal, intensity inhomogeneity correction, and registration. Finally, the data were down-sampled to a resolution of 128 × 128 × 128. Further specific details can be found in (Jack et al., 2024). Hippocampal volumes were extracted using the longitudinal processing method within FreeSurfer (version 6.0). The method utilizes Bayesian inference jointly computing different timepoints (Iglesias et al., 2016). To avoid a processing bias it treats all time points the same creating an unbiased subject specific T1-weighted template. Multiple assessments agree that the test-retest reliability for this method is good, and additionally it is reported that hippocampal volumetry had a high intraclass coefficient across field strengths (1.5 and 3 tesla) (Sämann et al., 2020; Whelan et al., 2016). Hippocampal subfield analyses of hippocampal volumes were considered exploratory, given the limited visibility of the stratum radiatum lacunosum moleculare (SRLM; or SLM in case of the subiculum) – an important feature for discerning and labelling the different hippocampal subfields (Wisse et al., 2021). Our analyses focused on subiculum subregions (subiciulum proper, presubiculum, parasubiculum) because these regions can be relatively well discerned on T1-weighted MRI, as most of the borders consist of white matter in the parahippocampal gyrus and cerebrospinal fluid in the hippocampal sulcus and ambient cistern, and accurate labelling is in only a small portion dependent on adequate visualization of SRLM. Additionally, the subiculum regions harbor relatively early and prominent tau pathology and are also an early site for amyloid-beta pathology within the medial temporal lobe in AD (Denning et al., 2024; Jellinger, 2018; Murray et al., 2011; Thal et al., 2000).

### Genotyping

ADNI participants were genotyped using Illumina’s Human 610-Quad BeadChip (ADNI-1) or HumanOmniExpress BeadChip. Quality control (QC) employed PLINK software, excluding samples with call rates below 98%, sex mismatches, excessive heterozygosity, or significant genetic relatedness (IBD>0.185) (Purcell et al., 2007). Post-QC, variants failing criteria for minor allele frequency (MAF<1%), Hardy-Weinberg equilibrium (p<10^-6^), or having high missingness (>5%) were removed. Genetic imputation used the Michigan Imputation Server with the HRC r1.1 2016 panel, following the protocols reported in (Vilor-Tejedor et al., 2024), including pre-imputation filtering and phasing via EAGLE v.2.4, with a quality threshold of >0.2 and MAF >1%.

### Genetic predisposition to Alzheimer’s disease

The genetic predisposition to AD (PRS-AD) was calculated by summing the alleles of single nucleotide polymorphisms (SNPs) carried by participants, weighting them according to the SNP allele effect sizes estimated from a previous genome-wide association study (Bellenguez et al., 2022), and normalizing the scores by the total number of included SNPs using PRSice version 2 (Choi & O’Reilly, 2019). PRS-AD was calculated using representative genetic variants per linkage disequilibrium block (LD) (referred to as clumped variants), with an LD cutoff of r^2 > 0.1 within a 250-kb window. PRS-AD was also calculated excluding the *APOE* region (chr19:45,409,011-45,412,650; GRCh37/hg19) (PRS-AD_noAPOE_). The individual effect of ApoE CSF protein associated QTLs (pQTLs, n=4), and *APOE* brain gene expression associated QTLS (eQTLs, n=6) (Aguet et al., 2020) [**Supplemental Table 1**], was assessed as well as the effect of *APOE*-ε4 carriership.

### Statistical analysis

Demographic and genetic characteristics of the sample are summarized in **Table 1** by diagnostic group, and **Table 2** by sex. The significance of any observed differences according to diagnostic group or sex was determined using either χ^2^ tests for categorical variables or parametric tests (t-test or ANOVA) and non-parametric tests (Wilcoxon rank-sum or Kruskal-Wallis) for normally distributed and non-normally distributed numerical variables, respectively. Subsequently, pairwise comparisons between diagnostic groups were conducted using t-tests with Tukey’s correction to explore the nature of these unadjusted differences in hippocampal volumes (response variables) and polygenic risk scores (explanatory variables). The normality of the distribution for hippocampal volumes was assessed through visual inspection of histograms [**Supplemental Figure 1**]. In addition, for the longitudinal volume measurements, the number of participants with measurements at each time point of the study (baseline, 3-and 6-month visits, and annual visits from the first to the fifth year of follow-up) was reported for each diagnostic group [**Figure 1**, **Supplemental Table 2**]. Observations with less than 5% of the sample in follow-up for each diagnostic group were excluded. Baseline associations were assessed using generalized linear regression models adjusted by age, sex, years of education, diagnostic group, parameter for different scannes and total intracranial volume. The baseline hippocampal measures corresponded to those obtained in visit 1 when participants completed the MRI for the first time. Additionally, we assessed whether these effects differed according to diagnostic group and sex by including an interaction term between sex and/or disease status and the genetic features. We analyzed the association between genetic predisposition to AD and changes in hippocampal volumes repeatedly assessed over a 5-years period using linear mixed-effect models with random-time slope and intercept for individuals and including age, sex, years of education, diagnostic group, tesla parameter and total intracranial volume as fixed effects. Hippocampal trajectories were also assessed by including a three-way interaction term in the model, where the annual average volume variation was calculated according to the diagnostic group (CU, MCI, AD) and AD genetic predisposition (Low-Intermediate/High) [*see details of the models in* **[Supplemental methods](https://docs.google.com/document/d/1aTT-Rfkgx20V9TcLFeH__Q5f_AmKxIb9-Ess2qNrplA/edit?tab=t.0)**]. In all models, a linear change in volumes over time was assumed. To verify if a nonlinear trajectory could be more appropriate, as suggested by several studies (Vilor-Tejedor et al., 2020; Vinke et al., 2018), a quadratic term for the time variable was included in post hoc analyses, both as a main fixed effect and in interaction with the diagnostic group. The null hypothesis of all coefficients associated with this term was tested using a Wald test for simultaneous contrasts. Finally, exploratory analyses were run with subiculum subregions instead of hippocampal volumes. Multiple comparison correction was applied using the False Discovery Rate (FDR) method to adjust for multiple statistical tests conducted in the study.

**Table 1.** Baseline characteristics by diagnostic group. Note: Results are reported as mean (standard deviation) or median [Q1, Q3] quartile for normal and non-normal numeric variables, respectively; and as the number of cases and proportion n (p%) for categorical variables. Differences between diagnostic groups were tested using ANOVA or the Kruskal-Wallis test for numeric normal or non-normal variables, respectively; and using a Chi-squared test for categorical variables. P-values are reported.

|  | Total<br>N = 1051 | Controls<br>N = 363 | Mild Cognitively Impairment<br>N = 474 | Alzheimer's Disease<br>N = 214 | Pvalue |
| --- | --- | --- | --- | --- | --- |
| **Demographic characteristics** |  |  |  |  |  |
| Age at baseline (years; mean, sd) | 74.2 (6.9) | 74.3 (6.0) | 73.8 (7.1) | 74.6 (7.8) | 0.322 |
| Sex (n, %) |  |  |  |  | **<0.001** |
| Female | 473 (45.0%) | 189 (52.1%) | 185 (39.0%) | 99 (46.3%) |  |
| Male | 578 (55.0%) | 174 (47.9%) | 289 (61.0%) | 115 (53.7%) |  |
| Education level (years; median, range) | 16.00 [14.00, 18.00] | 16.00 [14.00, 18.00] | 16.00 [14.00, 18.00] | 16.00 [12.00, 17.00] | **<0.001** |
| APOE-ε4 carriership (n, %) |  |  |  |  | **<0.001** |
| Non carriers | 471 (46.8%) | 242 (71.2%) | 171 (37.3%) | 58 (28.0%) |  |
| 1 allele carriers | 421 (41.8%) | 89 (26.2%) | 224 (48.8%) | 108 (52.2%) |  |
| 2 allele carriers | 114 (11.3%) | 9 (2.6%) | 64 (13.9%) | 41 (19.8%) |  |
| Intracranial volume (cm<sup>3</sup>; mean, sd) | 1539 (178) | 1523 (166) | 1554 (181) | 1534 (187) | **0.037** |
| **Genetic Scores (N at high risk, %)** |  |  |  |  |  |
| AD (High predisposition) | 208 (19.8%) | 32 (8.8%) | 120 (25.3%) | 56 (26.2%) | **<0.001** |
| AD excluding APOE region (High predisposition) | 203 (19.3%) | 64 (17.6%) | 99 (20.9%) | 40 (18.7%) | 0.481 |
| **SNPs (N, %)** |  |  |  |  |  |
| eQTL Brain - rs157584 (**minor allele: C**) |  |  |  |  | 0.173 |
| Non carriers | 318 (30.3%) | 95 (26.2%) | 152 (32.1%) | 71 (33.2%) |  |
| 1 minor allele carriers | 527 (50.1%) | 193 (53.2%) | 225 (47.5%) | 109 (50.9%) |  |
| 2 minor allele carriers | 206 (19.6%) | 75 (20.7%) | 97 (20.5%) | 34 (15.9%) |  |
| eQTL Brain - rs157588 (**minor allele: T**) |  |  |  |  | 0.078 |
| Non carriers | 333 (31.7%) | 98 (27.0%) | 159 (33.5%) | 76 (35.5%) |  |
| 1 minor allele carriers | 518 (49.3%) | 190 (52.3%) | 221 (46.6%) | 107 (50.0%) |  |
| 2 minor allele carriers | 200 (19.0%) | 75 (20.7%) | 94 (19.8%) | 31 (14.5%) |  |
| eQTL Brain - rs157590 (**minor allele: C**) |  |  |  |  | **0.047** |
| Non carriers | 330 (31.4%) | 95 (26.2%) | 158 (33.3%) | 77 (36.0%) |  |
| 1 minor allele carriers | 522 (49.7%) | 193 (53.2%) | 223 (47.0%) | 106 (49.5%) |  |
| 2 minor allele carriers | 199 (18.9%) | 75 (20.7%) | 93 (19.6%) | 31 (14.5%) |  |
| eQTL Brain rs34878901 (**minor allele: T**) |  |  |  |  | **0.003** |
| Non carriers | 482 (45.9%) | 146 (40.2%) | 223 (47.0%) | 113 (52.8%) |  |
| 1 minor allele carriers | 467 (44.4%) | 170 (46.8%) | 205 (43.2%) | 92 (43.0%) |  |
| 2 minor allele carriers | 102 (9.7%) | 47 (12.9%) | 46 (9.7%) | 9 (4.2%) |  |
| eQTL Brain rs1305062 (**minor allele: C**) |  |  |  |  | **0.003** |
| Non carriers | 486 (46.2%) | 149 (41.0%) | 223 (47.0%) | 114 (53.3%) |  |
| 1 minor allele carriers | 464 (44.1%) | 166 (45.7%) | 207 (43.7%) | 91 (42.5%) |  |
| 2 minor allele carriers | 101 (9.6%) | 48 (13.2%) | 44 (9.3%) | 9 (4.2%) |  |
| eQTL Brain rs769450 (**minor allele: A**) |  |  |  |  | **0.003** |
| Non carriers | 492 (46.8%) | 150 (41.3%) | 226 (47.7%) | 116 (54.2%) |  |
| 1 minor allele carriers | 458 (43.6%) | 165 (45.5%) | 205 (43.2%) | 88 (41.1%) |  |
| 2 minor allele carriers | 101 (9.6%) | 48 (13.2%) | 43 (9.1%) | 10 (4.7%) |  |
| pQTL CSF rs769449 (**minor allele: A**) |  |  |  |  | **<0.001** |
| Non carriers | 624 (59.4%) | 284 (78.2%) | 251 (53.0%) | 89 (41.6%) |  |
| 1 minor allele carriers | 353 (33.6%) | 73 (20.1%) | 179 (37.8%) | 101 (47.2%) |  |
| 2 minor allele carriers | 74 (7.0%) | 6 (1.7%) | 44 (9.3%) | 24 (11.2%) |  |
| pQTL CSF rs429358 (**minor allele: C**) |  |  |  |  | **<0.001** |
| Non carriers | 509 (48.4%) | 256 (70.5%) | 192 (40.5%) | 61 (28.5%) |  |
| 1 minor allele carriers | 433 (41.2%) | 96 (26.4%) | 225 (47.5%) | 112 (52.3%) |  |
| 2 minor allele carriers | 109 (10.4%) | 11 (3.0%) | 57 (12.0%) | 41 (19.2%) |  |
| pQTL CSF rs7412 (**minor allele: T**) |  |  |  |  | **0.010** |
| Non carriers | 968 (92.1%) | 322 (88.7%) | 444 (93.7%) | 202 (94.4%) |  |
| 1 allele carriers | 81 (7.7%) | 40 (11.0%) | 30 (6.3%) | 11 (5.1%) |  |
| 2 allele carriers | 2 (0.2%) | 1 (0.3%) | 0 (0.0%) | 1 (0.5%) |  |
| pQTL CSF rs75627662 (**minor allele: T**) |  |  |  |  | **<0.001** |
| Non carriers | 557 (53.0%) | 246 (67.8%) | 229 (48.3%) | 82 (38.3%) |  |
| 1 minor allele carriers | 397 (37.8%) | 108 (29.8%) | 188 (39.7%) | 101 (47.2%) |  |
| 2 minor allele carriers | 97 (9.2%) | 9 (2.5%) | 57 (12.0%) | 31 (14.5%) |  |
| **Hippocampal volumes (cm3, mean, sd)** |  |  |  |  |  |
| Whole hippocampus (cm3) | 5873 (1044) | 6525 (736) | 5771 (993) | 4995 (865) | **<0.001** |
| subiculum (cm3) | 663 (136) | 745 (102) | 651 (128) | 553 (116) | **<0.001** |
| presubiculum (cm3) | 481 (111) | 547 (87) | 470 (107) | 393 (84) | **<0.001** |
| parasubiculum (cm3) | 105.2 (27.6) | 113.9 (23.6) | 103.4 (28.7) | 94.3 (27.1) | **<0.001** |

**Table 2.** Baseline characteristics by sex. . Note: Results are reported as mean (standard deviation) or median [Q1, Q3] quartile for normal and non-normal numeric variables, respectively; and as the number of cases and proportion n (p%) for categorical variables. Differences between diagnostic groups were tested using ANOVA or the Kruskal-Wallis test for numeric normal or non-normal variables, respectively; and using a Chi-squared test for categorical variables. P-values are reported.

|  | Total<br>N = 1051 | Female<br>N = 473 | Male<br>N = 578 | Pvalue |
| --- | --- | --- | --- | --- |
| **Demographic characteristics** |  |  |  |  |
| Age at baseline (years; mean, sd) | 74.2 (6.9) | 73.2 (7.0) | 74.9 (6.7) | **<0.001** |
| Diagnostic group (N, %) |  |  |  | **<0.001** |
| *Cognitively healthy controls* | 363 (34.5%) | 189 (40.0%) | 174 (30.1%) |  |
| *Mild Cognitive Impairment* | 474 (45.1%) | 185 (39.1%) | 289 (50.0%) |  |
| *Alzheimer's Disease* | 214 (20.4%) | 99 (20.9%) | 115 (19.9%) |  |
| Education level (years; median, range) | 16.00 [14.00, 18.00] | 16.00 [13.00, 18.00] | 16.00 [15.00, 18.00] | **<0.001** |
| APOE-ε4 carriership (n, %) |  |  |  | 0.508 |
| *Non carriers* | 471 (46.8%) | 218 (48.3%) | 253 (45.6%) |  |
| *1 allele carriers* | 421 (41.8%) | 187 (41.5%) | 234 (42.2%) |  |
| *2 allele carriers* | 114 (11.3%) | 46 (10.2%) | 68 (12.3%) |  |
| Intracranial volume (cm<sup>3</sup>; mean, sd) | 1539 (178) | 1421 (133) | 1636 (149) | **<0.001** |
| **Genetic Scores (N at high risk, %)** |  |  |  |  |
| *AD (High predisposition)* | 208 (19.8%) | 87 (18.4%) | 121 (20.9%) | 0.342 |
| *AD excluding APOE region (High predisposition)* | 203 (19.3%) | 95 (20.1%) | 108 (18.7%) | 0.622 |
| **SNPs (N, %)** |  |  |  |  |
| eQTL Brain - rs157584 (*minor allele: C*) |  |  |  | 0.414 |
| *Non carriers* | 318 (30.3%) | 147 (31.1%) | 171 (29.6%) |  |
| *1 minor allele carriers* | 527 (50.1%) | 227 (48.0%) | 300 (51.9%) |  |
| *2 minor allele carriers* | 206 (19.6%) | 99 (20.9%) | 107 (18.5%) |  |
| eQTL Brain - rs157588 (*minor allele: T*) |  |  |  | 0.716 |
| *Non carriers* | 333 (31.7%) | 152 (32.1%) | 181 (31.3%) |  |
| *1 minor allele carriers* | 518 (49.3%) | 227 (48.0%) | 291 (50.3%) |  |
| *2 minor allele carriers* | 200 (19.0%) | 94 (19.9%) | 106 (18.3%) |  |
| eQTL Brain - rs157590 (*minor allele: C*) |  |  |  | 0.684 |
| *Non carriers* | 330 (31.4%) | 152 (32.1%) | 178 (30.8%) |  |
| *1 minor allele carriers* | 522 (49.7%) | 228 (48.2%) | 294 (50.9%) |  |
| *2 minor allele carriers* | 199 (18.9%) | 93 (19.7%) | 106 (18.3%) |  |
| eQTL Brain rs34878901 (*minor allele: T*) |  |  |  | 0.882 |
| *Non carriers* | 482 (45.9%) | 214 (45.2%) | 268 (46.4%) |  |
| *1 minor allele carriers* | 467 (44.4%) | 211 (44.6%) | 256 (44.3%) |  |
| *2 minor allele carriers* | 102 (9.7%) | 48 (10.1%) | 54 (9.3%) |  |
| eQTL Brain rs1305062 (*minor allele: C*) |  |  |  | 0.864 |
| *Non carriers* | 486 (46.2%) | 218 (46.1%) | 268 (46.4%) |  |
| *1 minor allele carriers* | 464 (44.1%) | 207 (43.8%) | 257 (44.5%) |  |
| *2 minor allele carriers* | 101 (9.6%) | 48 (10.1%) | 53 (9.2%) |  |
| eQTL Brain rs769450 (*minor allele: A*) |  |  |  | 0.756 |
| *Non carriers* | 492 (46.8%) | 220 (46.5%) | 272 (47.1%) |  |
| 1 minor allele carriers | 458 (43.6%) | 204 (43.1%) | 254 (43.9%) |  |
| 2 minor allele carriers | 101 (9.6%) | 49 (10.4%) | 52 (9.0%) |  |
| pQTL CSF rs769449 (**minor allele: A**) |  |  |  | 0.906 |
| Non carriers | 624 (59.4%) | 284 (60.0%) | 340 (58.8%) |  |
| 1 minor allele carriers | 353 (33.6%) | 157 (33.2%) | 196 (33.9%) |  |
| 2 minor allele carriers | 74 (7.0%) | 32 (6.8%) | 42 (7.3%) |  |
| pQTL CSF rs429358 (**minor allele: C**) |  |  |  | 0.586 |
| Non carriers | 509 (48.4%) | 231 (48.8%) | 278 (48.1%) |  |
| 1 minor allele carriers | 433 (41.2%) | 198 (41.9%) | 235 (40.7%) |  |
| 2 minor allele carriers | 109 (10.4%) | 44 (9.3%) | 65 (11.2%) |  |
| pQTL CSF rs7412 (**minor allele: T**) |  |  |  | 0.862 |
| Non carriers | 968 (92.1%) | 434 (91.8%) | 534 (92.4%) |  |
| 1 allele carriers | 81 (7.7%) | 38 (8.0%) | 43 (7.4%) |  |
| 2 allele carriers | 2 (0.2%) | 1 (0.2%) | 1 (0.2%) |  |
| pQTL CSF rs75627662 (**minor allele: T**) |  |  |  | 0.607 |
| Non carriers | 557 (53.0%) | 254 (53.7%) | 303 (52.4%) |  |
| 1 minor allele carriers | 397 (37.8%) | 172 (36.4%) | 225 (38.9%) |  |
| 2 minor allele carriers | 97 (9.2%) | 47 (9.9%) | 50 (8.7%) |  |
| **Hippocampal volumes (cm3, mean, sd)** |  |  |  |  |
| Whole hippocampus (mm3) | 5873 (1044) | 5647 (1045) | 6058 (1008) | **<0.001** |
| subiculum (mm3) | 663 (136) | 636 (135) | 686 (133) | **<0.001** |
| presubiculum (mm3) | 481 (111) | 468 (110) | 492 (111) | **<0.001** |
| parasubiculum (mm3) | 105.2 (27.6) | 99.3 (25.6) | 110.0 (28.3) | **<0.001** |

## RESULTS

### Descriptives

The sample was characterized by participants in three different diagnostic groups: CU (N=363), MCI due to AD (N=474), and individuals diagnosed with AD dementia (N=214). **Table 1** presents the baseline characteristics of participants by diagnostic group. Although the average age did not differ significantly across diagnosis groups (p=0.322), sex distributions showed significant differences, with a higher proportion of women among CU individuals (52.1%) compared to those with MCI (39%) and AD (46.3%). Years of education also varied significantly, with the median education level slightly lower in the AD group than in the CU and MCI groups. Genetic factors also revealed significant differences among diagnostic groups. *APOE*-ε4 carriership increased with disease severity: while 71.2% of CU are non-carriers, this percentage decreased to 37.3% in MCI and 28.0% in AD. In contrast, the proportion of individuals with one or two *APOE*-ε4 alleles was substantially higher in the AD group (52.2% and 19.8%, respectively) compared to CU (26.2% and 2.6%, respectively). Similarly, high genetic predisposition to AD was more frequent in MCI (25.3%) and AD (26.2%) groups compared to CU (8.8%).

**Table 2** presents the baseline characteristics of the study participants by sex. Age at baseline differed slightly between sexes, with men being slightly older on average (74.9 years) compared to women (73.2 years) (p<0.001). Diagnostic group distribution also varied by sex, with a higher percentage of CU among women (40.0%) than men (30.1%), while men were more frequently diagnosed with MCI (50.0%) compared to women (39.1%). Years of education showed a slight difference, with men having a higher median education level compared to women. However, there is no significant difference between sexes in *APOE*-ε4 carriership, with a similar distribution of non-carriers, one allele carrier, and two allele carriers across both sexes (p=0.508). Regarding AD genetic risk, no significant sex difference was observed, with 18.4% of women and 20.9% of men classified as high risk (p=0.342). Similarly, genetic scores excluding the *APOE* region showed no significant differences between sexes (p=0.622) [**Supplemental Figure 2**]. Moreover, we observed significant differences in the mean hippocampal volumes at the baseline visit, both among diagnostic groups and between sexes [**Supplemental Table 3**]. For all hippocampal regions, AD and MCI exhibited, on average, greater estimated annual volume loss compared to controls [**Supplemental Table 4**].

### Baseline analysis

At baseline, significant associations were observed between *APOE*-ε4 allele carriers and genetically predisposed individuals with reduced hippocampal and subiculum volumes, as well as CSF pQTLs [**Figure 2, Supplemental Table 5**]. When stratified by disease status, we found that the significant genetic associations were predominantly observed in the MCI and AD groups [**Supplementary Tables 6-8**]. Although no sex-specific or diagnosis-group specific interactions were observed at baseline, subsequent analysis stratified by sex revealed significant associations. In particular, genetic predisposition to AD was primarily associated with hippocampal volume, and reduced subiculum volume, in men classified with MCI, and in whole hippocampal volume in women diagnosed with AD [**Figure 3, Supplementary Table 9**]. No significant interactions were found with brain eQTLs [**Supplementary Figure 3**].

**Figure 2.**
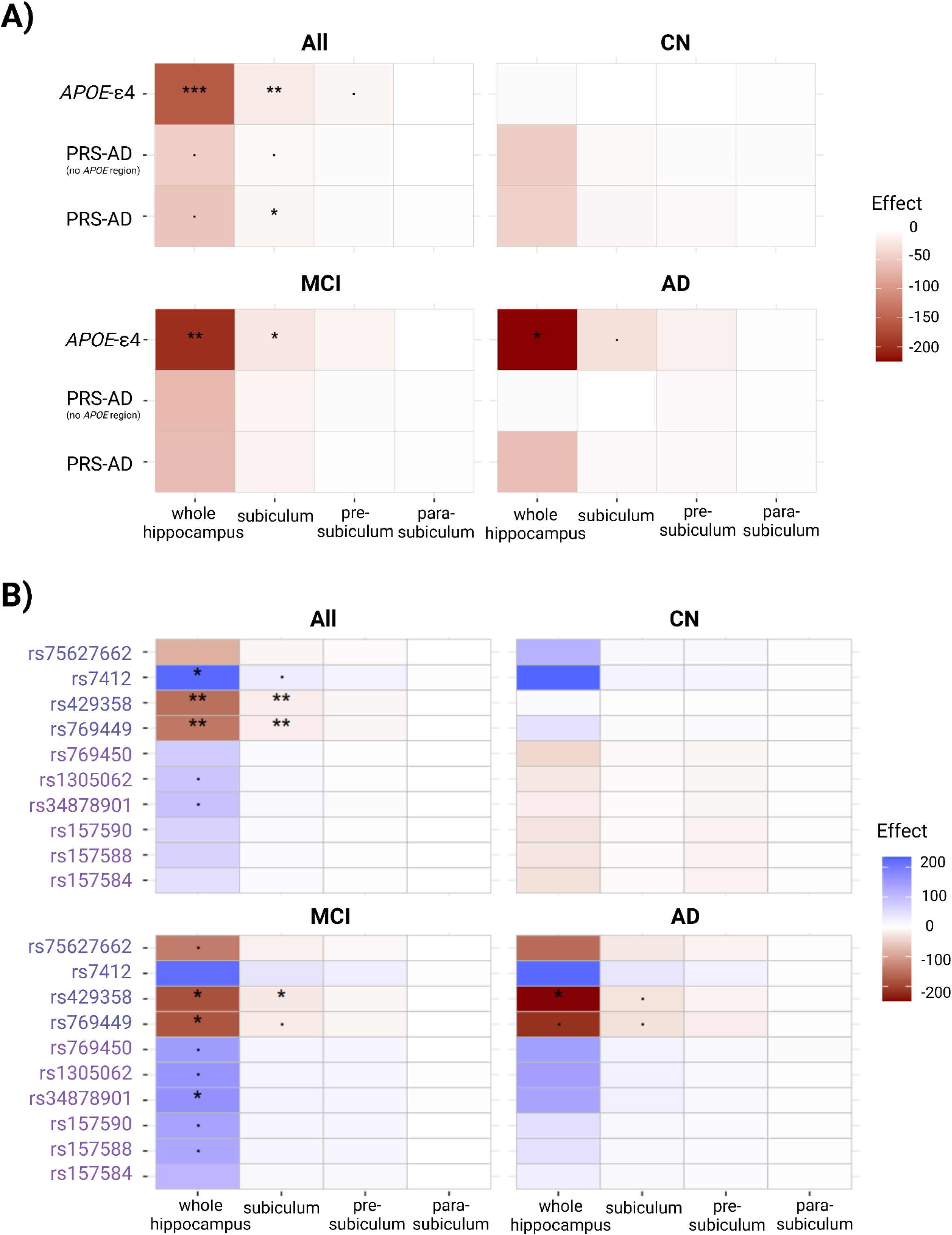
Cross-sectional global and stratified by disease stage models at baseline. P-values are reported as *** <.001, ** <.01, and * <.05 after correcting for multiple comparisons. Effects adjusted for age at baseline, intracranial volume, sex, diagnostic status, years of education and tesla scan parameter. A) Adjusted effect of PRS-AD, PRS-AD_noAPOE_, and *APOE*-ε4 carriership. Heatmap displays genetic main effect lineal coefficients. B) Adjusted effect of ApoE pQTLs and *APOE* brain eQTLs.

**Figure 3.**
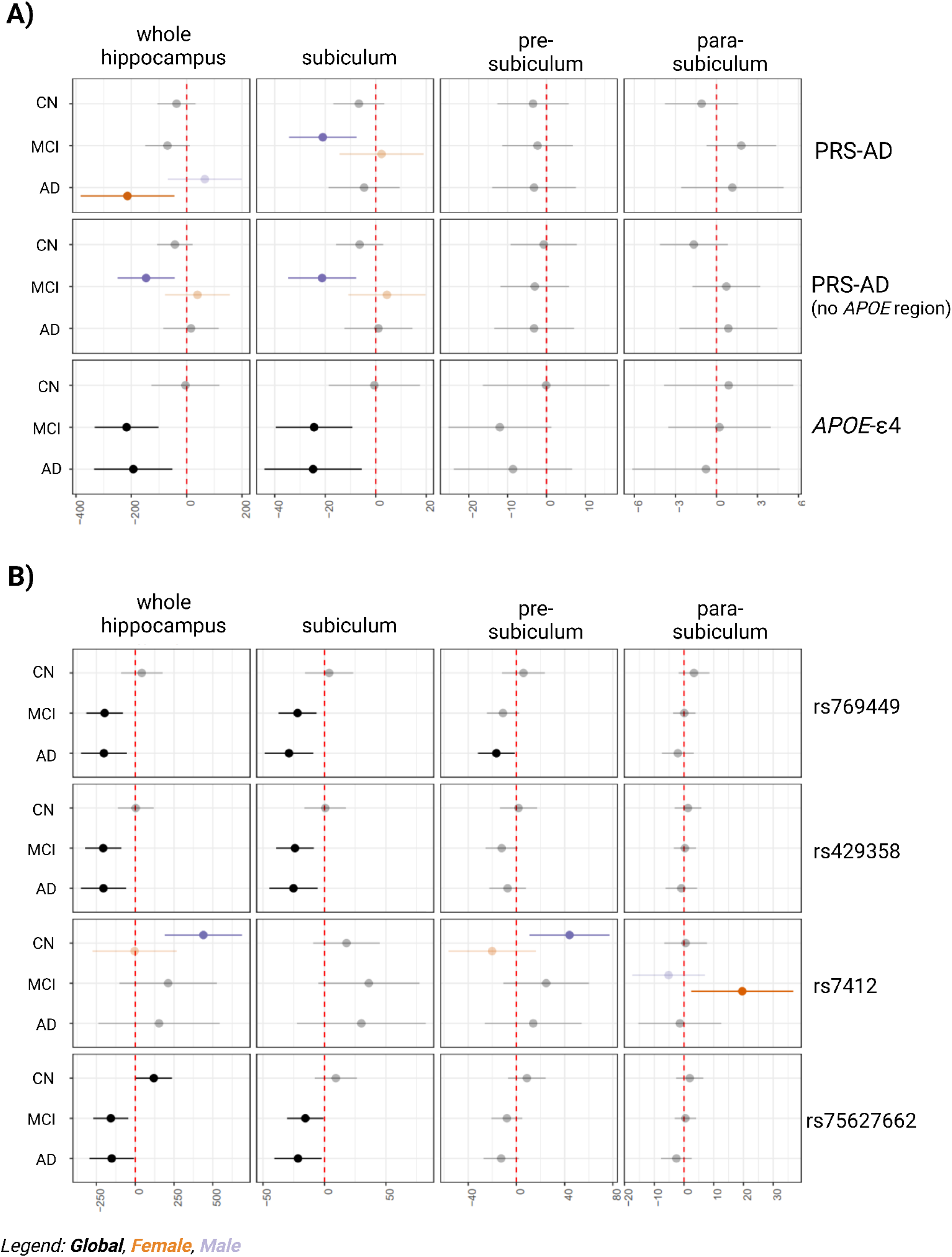
Cross-sectional model at baseline with disease status interaction. A) Adjusted effect of PRSs and *APOE*-ε4 carriership on hippocampal volumes at baseline, estimated for each diagnosis groups by adding interaction term PRS*Disease status. A) Heatmap shows the PRS effect for each disease group as its linear combination of main effect + interaction term. B) Heatmap shows the pQTL and eQTL effect for each disease group as its linear combination of main effect + interaction term.

### Longitudinal analysis of hippocampal trajectories

#### Characterization of annual volume changes in the hippocampus

We observed a statistically significant reduction in hippocampal volumes over time in all groups. This reduction was estimated as the expected volume change for each year of follow-up. This atrophy was notably more pronounced in participants with MCI and AD compared to CU individuals. Both the atrophy trajectories and the differences between CU, MCI and AD were consistent with exploratory graphs of the average volume at each measurement point, smoothed using *Loess* methodology [**Figure 4]**.

**Figure 4.**
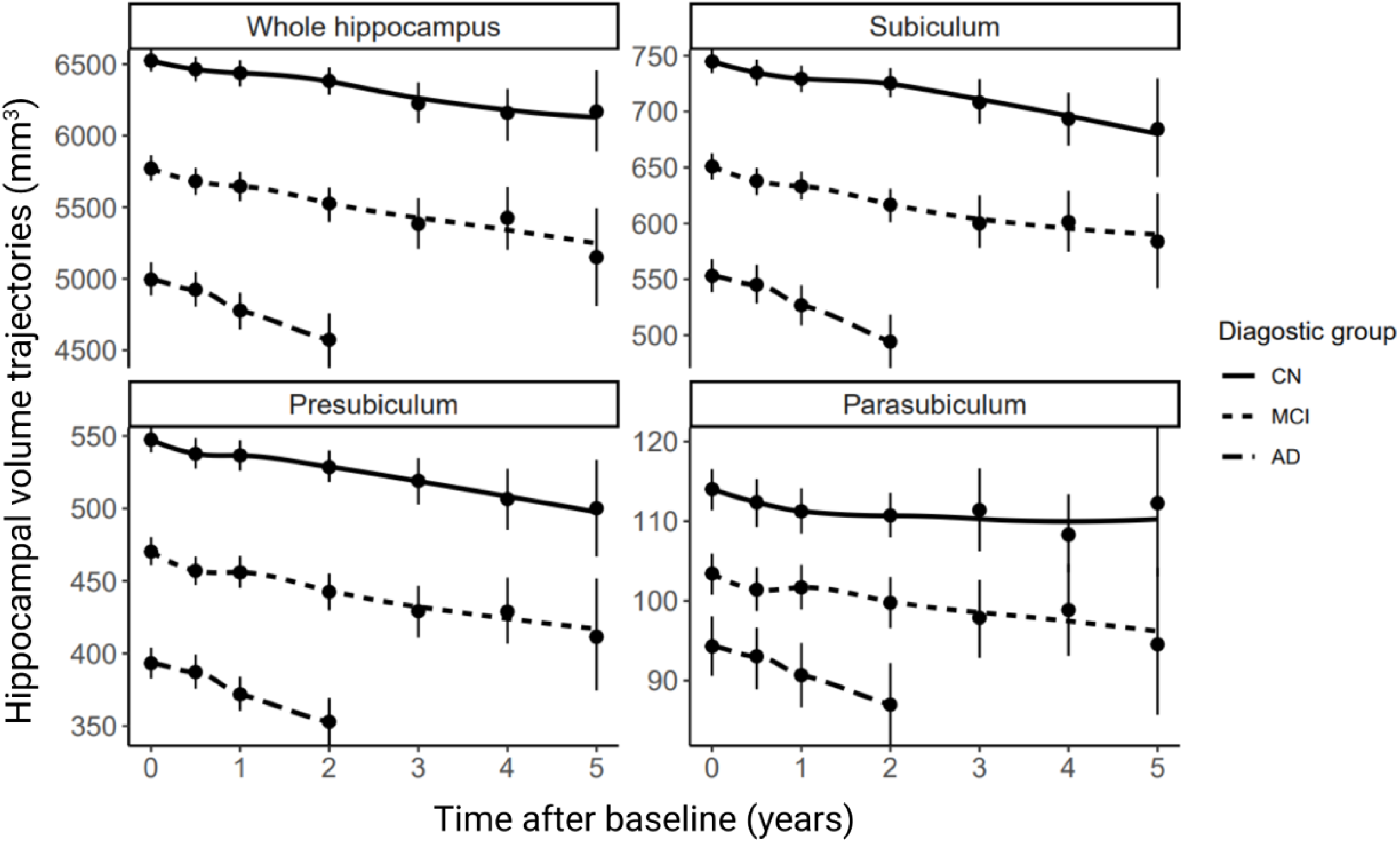
Smoothed trajectories of hippocampal mean volumes at each visit stratified by disease stage.

#### Impact of genetic predisposition to AD on hippocampal trajectories

High genetic predisposition to AD was associated with increased hippocampal atrophy rates. However, when excluding variants in the *APOE* region from the PRS, this genetic influence on volume trajectories was no longer significant, suggesting that the *APOE* region plays a central role in hippocampal atrophy in individuals with a genetic predisposition to AD [**Figure 5A**]. Further analysis of *APOE*’s role revealed that *APOE*-ε4 status, as well as CSF pQTLs were driving the association with hippocampal volume decline over time [**Figure 5B,Supplemental Table 10**]. For the explored subfields, the effects were largest for the subiculum and presubiculum, and smaller, but still significant, for the parasubiculum.

**Figure 5.**
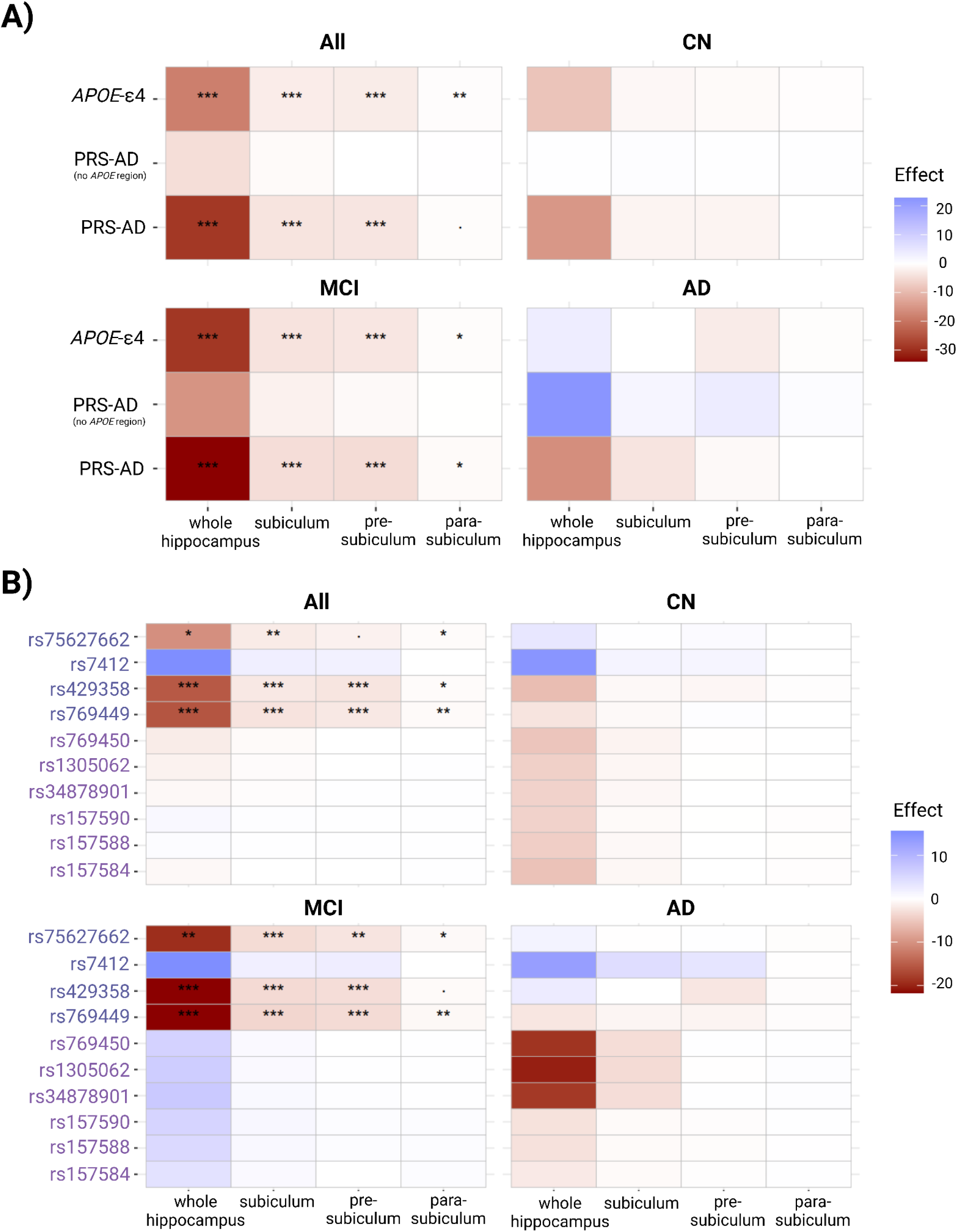
Longitudinal models stratified by disease status, with time*genetic interaction. A) Adjusted PRS effect (binary; high-low predisposition) or allele effect (categorical; 0,1,2), on hippocampal volume’s rate of change over time. Heatmaps display interaction terms A) *time*PRS* and *time*APOE-ε4carriership* (B) *time*QTL-alleles*; for global and stratified models.

#### Diagnostic group-related differences in genetic effect on hippocampal trajectories

The effect of AD genetic predisposition on hippocampal atrophy depended on the diagnostic group, as indicated by a significant interaction term [**Figure 6A**]. Individuals with MCI with a high genetic predisposition to AD and/or *APOE*-ε4 carriers exhibited a more severe atrophy rate (nominal significance) compared to those with a lower genetic predisposition. We additionally found significant associations between CSF pQTLs with accelerated volume decrease only in MCI. Models stratified by the diagnostic group revealed that the *APOE*-related effects were predominantly linked to accelerated atrophy in MCIs, with CU and AD showing similar but reduced and nonsignificant differences [**Figure 6B, Supplemental Table 11**]. Similar effects were observed for the hippocampus, subiculum and presubiculum. No significant interactions were found with brain eQTLs [**Supplementary Figure 4**].

**Figure 6.**
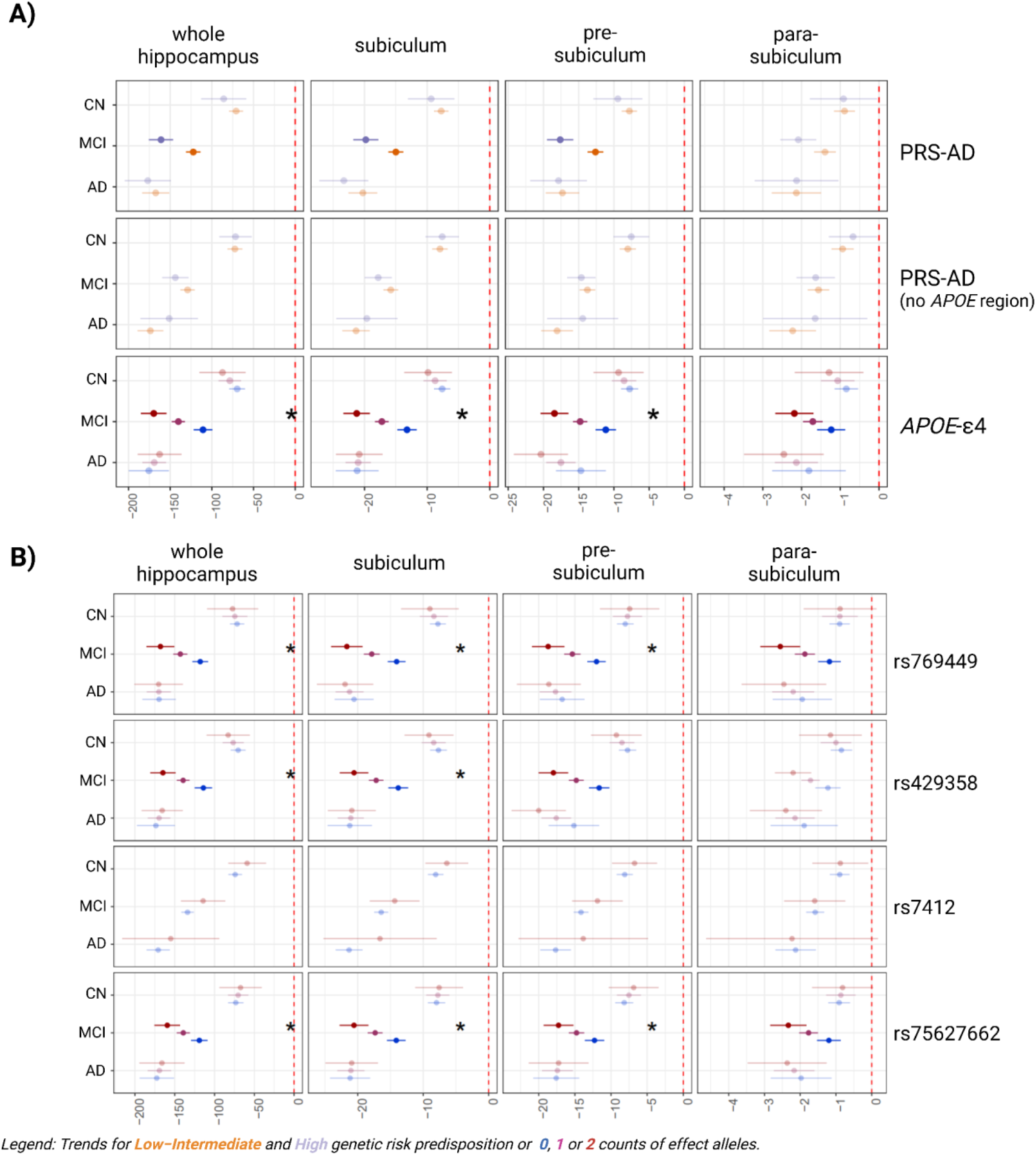
Longitudinal models with disease status interaction. Adjusted PRS effect (binary, high-low genetic predisposition) on hippocampal volume’s rate of change over time, global and stratified models by disease stage. A) Heatmap displays effects stratified by diagnostic group as well as significant interaction term *time*Disease status*PRS* (*), which estimates differences in rate of change associated with being at high predisposition for a given PRS or *APOE*-ε4 carriership. B) Heatmap displays effects stratified by diagnostic group as well as significant interaction term *time*Disease status*pQTLs* (*).

#### Sex-related differences in genetic effect on hippocampal trajectories

Accelerated hippocampal atrophy associated with a higher genetic predisposition to AD were observed irrespective of sex, driven by CSF pQTL variants. These genetic effects were significant in both men and women [**Supplemental Figure 5, Supplemental Table 12**].

## DISCUSSION

Our study revealed that the impact of genetic predisposition to AD, particularly driven by the *APOE* region, significantly accelerates the atrophy of the hippocampal region in individuals with MCI but not in individuals diagnosed with AD or healthy controls. This finding suggests a stage-specific effect of genetic factors on disease progression, which is critical for understanding the temporal dynamics of AD pathology and for designing targeted interventions. The exclusive observation in individuals with MCI might be attributed to several factors. MCI is often considered an intermediate condition between normal aging and AD, where neurodegenerative changes are underway but not yet pronounced enough to impair daily functioning significantly. It is plausible that the hippocampus, being a primary site of early AD pathology, is more susceptible to genetic influences during this phase. The specific acceleration of atrophy might thus reflect an increased vulnerability or a heightened response to pathological processes in individuals at higher AD genetic predisposition. Moreover, it is plausible that the bulk of hippocampal atrophy driven by genetic predisposition, particularly factors associated with the *APOE* region, occurs before or during the transition to MCI. By the time an individual is diagnosed with AD dementia, significant atrophy may already have taken place, especially in the hippocampus affecting memory. Although AD progression is mainly driven by a cascade of neuropathological events, genetic predispositions, particularly *APOE*-ε4 carriership, may modulate this process by facilitating the accumulation of amyloid-beta in the medial temporal lobe and hippocampus. Studies have shown that *APOE*-ε4 is associated with increased tau pathology, potentially even without amyloid-beta presence (Therriault et al., 2022; Young et al., 2023). Therefore, while direct genetic predisposition alone may not entirely drive AD progression, it can contribute to neurodegeneration by accelerating neuropathological changes in areas crucial for memory and cognition. Interestingly, strongest associations of *APOE*-ε4 carriership were observed with the subiculum, in both cross-sectional and longitudinal analyses. As the subiculum is a relatively prominent site of both tau and amyloid beta deposits within the hippocampus, it is possible that the observed effect of *APOE*-ε4 carriership on subiculum atrophy, may occur through AD neuropathological processes (Bouras et al., 1994; Denning et al., 2024; Murray et al., 2011; Thal et al., 2000). Finally, *APOE*-ε4 is associated with heightened neuroinflammation, which further accelerates the progression of neurodegeneration. These pathological processes appear to be most pronounced in individuals with MCI, which may represent a critical tipping point in the disease trajectory. During this stage, the combined effects of Aβ accumulation, tau deposition, and inflammation may cross a threshold, leading to pronounced neuronal loss and cognitive decline. This underscores the importance of the MCI stage as a window for intervention, as the pathological processes underlying neurodegeneration are more susceptible to genetic influence at this juncture.

In controls, the intact cognitive and hippocampal function may exhibit resilience to genetic predispositions linked to AD. The lack of cognitive impairment and neurodegeneration in this group provides little context in which the subtle impacts of genetic risk factors on hippocampal atrophy can manifest, highlighting the role of these genetic factors primarily though disease pathology.

A critical aspect of our analysis involved dissecting the influence of the *APOE* region on hippocampal atrophy. Upon removing the *APOE* region from our genetic predisposition scores, we observed a loss of statistical significance. This result underscores the *APOE* region’s substantial contribution to the effect of genetic predisposition of AD on the hippocampus, aligning with previous literature (Håglin et al., 2023; Saeed et al., 2021). Furthermore, our investigation extended into the functional effects of the *APOE* region on the hippocampal trajectories. However, the stronger effect observed for PRS-AD compared to *APOE*-ε4 highlights the importance of polygenic influences in hippocampal atrophy, suggesting that genetic risk beyond *APOE* also plays a crucial role in shaping neurodegenerative trajectories, specially in MCI stages.

A surprising result from our study was the absence of sex interactions in the genetic effects on hippocampal volumetric changes. However, when our data were stratified by sex, nuanced differences emerged. Specifically, the genetic predisposition for AD as well as *APOE*-ε4 carriership were predominantly associated with hippocampal atrophy in women diagnosed with AD dementia (Duarte-Guterman et al., 2021; Y. T. T. Wang et al., 2021). This suggests that while genetic factors influencing hippocampal volume may operate similarly across sexes, their impact could be more pronounced or detectable in the context of AD dementia in women. Supportive of this hypothesis is the observation of higher levels of amyloid-beta and tau deposits in cornu ammonis 1 and the subiculum in AD dementia in women, compared to men (Liesinger et al., 2018). In contrast, in men with MCI, similar genetic associations were observed. This may indicate that the AD genetic predisposition affecting hippocampal trajectories manifests before the full clinical presentation of AD dementia in men, perhaps indicating a different trajectories or timelines for disease progression between sexes.

Our research stands out due to its longitudinal assessment, which encompasses multiple diagnostic groups. This approach is crucial in highlighting the significant role genetics play in hippocampal atrophy, particularly in the prodromal or MCI stage of AD. This insight is particularly valuable for the stratification and classification of clinical trial groups within AD research, ensuring that genetic factors are appropriately considered in the development and testing of new treatments.

Moreover, our study investigated the functional effects of genetic variants associated with ApoE protein and *APOE* expression on hippocampal volume trajectories. We emphasized the role of protein QTLs, particularly those linked to *APOE* isoforms, in modulating hippocampal atrophy. By examining the influence of *APOE*-QTLs on hippocampal structure over time, we provided new insights into AD pathophysiology and a more nuanced understanding of how genetic predisposition affects disease progression. In particular, our QTL analysis highlighted the importance of pQTLs linked to the ApoE isoform-defining SNPs, which was strongly associated with hippocampal trajectories. These SNPs alter the amino acid sequence and 3D structure of ApoE and influence its final concentration, likely through differences in protein affinity with receptors and other interacting molecules (Aslam et al., 2023). While these SNPs define the APOE isoform, the pQTLs measured in CSF reflect the actual circulating protein levels. The association of these pQTLs with hippocampal atrophy suggested that the effect is driven primarily by differences in APOE levels rather than isoform-specific structural properties. Notably, *APOE*-ε4 carriers exhibited lower circulating ApoE levels, possibly due to its increased tendency to aggregate and interact with other proteins, affecting its stability and clearance. The two well-known SNPs defining *APOE* isoforms (rs429358 and rs7412) do not act as brain eQTLs, reinforcing the idea that their impact on protein levels is mediated by post-transcriptional mechanisms rather than direct regulation of brain gene expression (Wang et al., 2024). However, a key challenge remains in disentangling whether the observed effects on hippocampal atrophy are driven primarily by differences in protein concentration, isoform-specific properties, or a combination of both. Further studies integrating multimodal datasets, including proteomic and functional assays, will be essential to clarify these mechanisms.

Further studies are additionally needed to explore the broader landscape of genetic factors influencing hippocampal atrophy and their interactions with environmental and lifestyle factors. Additionally, the functional roles of other genetic regions in AD pathology warrant comprehensive investigation. Longitudinal studies incorporating larger cohorts and diverse populations will be crucial in validating our findings and understanding the variability in AD progression at the individual level over time, especially as individuals transition from MCI to AD. Moreover, integrating biomarker data, particularly amyloid-beta, tau and other neurodegeneration markers, will be essential to better disentangle whether higher genetic predisposition (e.g., *APOE*-ε4) leads to accelerated hippocampal atrophy through faster amyloid-beta and/or tau accumulation (Cicognola et al., 2025). Exploring the therapeutic potential of targeting specific pQTLs related to *APOE* could offer promising avenues for treatment development.

In conclusion, our study highlights the major influence of AD genetic predisposition in hippocampal atrophy, especially among A+ MCI individuals. This highlights the critical role of early-stage-specific effects and the influence of *APOE* CSF-pQTLs. These insights not only advance our understanding of AD pathology but also pave the way for innovative approaches to early detection, risk assessment, and targeted intervention.

## Data Availability

Data used in preparation of this article was obtained from the Alzheimer's Disease Neuroimaging Initiative (ADNI) database (adni.loni.usc.edu).
Codes for all analyses are available online through the GitHub platform: https://github.com/GeneticNeuroStats/Mixed-effect-model-brain-genetic-trajectories
Main scripts can be also found in Supplementary materials.

## Data and Code Availability (mandatory unless there is no data or code used)

Data used in preparation of this article was obtained from the Alzheimer’s Disease Neuroimaging Initiative (ADNI) database (adni.loni.usc.edu).

Codes for all analyses are available online through the GitHub platform: https://github.com/GeneticNeuroStats/Mixed-effect-model-brain-genetic-trajectories

Main scripts can be also found in Supplementary materials.

## Author contributions

NV-T and TE conceived and designed the study, supervised the analyses and drafted the manuscript. AR performed all the analyses and generated all the available code. TE performed MRI segmentation and quantification. NV-T, AR, PG, BR-F, FA, WP, LW, TE provided interpretation. All authors helped write and/or revise the manuscript. All authors have read and approved the final version of the manuscript.

## Acknowledgements

Data used in preparation of this article were obtained from the Alzheimer’s Disease Neuroimaging Initiative (ADNI) database (adni.loni.usc.edu). As such, the investigators within the ADNI contributed to the design and implementation of ADNI and/or provided data but did not participate in analysis or writing of this report. A complete listing of ADNI investigators can be found at: http://adni.loni.usc.edu/wp-content/uploads/how_to_apply/ADNI_Acknowledgement_List.pdf

## Supplementary material

The Supplementary Material section includes additional tables that enhance the understanding of the findings of the present study. Description of [Supplementary Figures](https://drive.google.com/drive/u/1/folders/1LcpJkIgFwjlQ-PEHJzyk1XlBJ6lM4PmH) (SF). SF1. Histogram showing the distribution of hippocampal volumes. Normality was assessed through visual inspection. SF2. Comparison of Alzheimer’s disease genetic risk between disease stages and sex. SF3. Cross-sectional model at baseline with disease status interaction. Heatmap shows eQTL effect for each disease group as its linear combination of main effect and interaction term. SF4. Longitudinal models with disease status interaction. Heatmap displays effects stratified by diagnostic group as well as significant interaction term time*Disease status*eQTLs (*). SF5. Longitudinal models with sex interaction. Heatmaps display effects stratified by sex as well as significant interaction term A) *time*Sex*PRS* and *time*Sex*APOE-ε4* carriership (B) *time*QTL-alleles*; for global and stratified models. Description of <u>Supplemental Tables</u> (ST). ST1. Functional genetic variants included in the study. ST2. Number and timing of scans per time point by diagnostic group (Controls, N=363; Mild Cognitive Impairment, N=474; Alzheimer’s disease, N=214). ST3. Adjusted marginal means for hippocampal volumes at baseline by diagnostic group and sex. ST4. Estimated mean annual volume changes by diagnostic group. ST5. Results for the PRS/QTL effect on hippocampal volumes shown in Figure 2. ST6. Results for the PRS/QTL effect on hippocampal volumes shown in Figures 2 for CU individuals. ST7. Results for the PRS/QTL effect on hippocampal volumes shown in Figures 2 for MCI. ST8. Results for the PRS/QTL effect on hippocampal volumes shown in Figures 2 for AD. ST9. Results for the PRS/QTL effect on hippocampal volumes stratified by sex. ST10. Results for the PRS/QTL*time effect on hippocampal volumes shown in Figure 5. ST11. Results for atrophy rates on hippocampal volumes by diagnostic stage shown in Figure 6. ST12. Results for atrophy rates on hippocampal volumes by sex shown in Supplemental Figure 5.

