## Supplementary material for "Genetic drivers of hippocampal atrophy highlight the role of *APOE* functional variants and AD polygenicity in Mild Cognitive Impairment": modelling_auxFunctions.R

```
getModelParameters <- function(model){
  # Get fixed effects paramenters
  params <- extract_fixed_effects(model)
  
  # Get model name. volume:PRS
  aux <- sapply(c("volume","PRS"),function(x)attributes(model)[[x]])
  modName <- paste(aux,collapse=":")
  
  # Result
  res <- data.frame(model = modName,
                    Volume = aux[["volume"]],
                    PRS = aux[["PRS"]],
                    params)
}

ensambleParamsTable <- function(modelList,data=result){
  # CONTROL
  if(!(modelList %in% c("modGlobal","mod_3wayIntDX","mod_3wayIntSx"))){
    stop("Wrong model list name")
  }
  
  # Get models
  modelParams <- lapply(data,"[[",modelList)
  
  # Get parameter estimates
  modelParams <- lapply(modelParams,getModelParameters)
  
  # Merge dataframe for all models in a final table
  modelParams <- do.call(rbind,modelParams)
  
  rownames(modelParams) <- NULL
  return(modelParams)
}

get_XsecModelParams <- function(model){
  smod <- summary(model)$coefficients
  terms <- rownames(smod)
  ci <- confint.default(model)
  att <- Reduce("c",attributes(model)[c("volume","PRS")])
  res <- data.frame(model = paste(att,collapse=":"),
                    Volume = att[1],
                    PRS = att[2],
                    terms,
                    smod,
                    ci)
  names(res) <- c("model","Volume","PRS","term","value","SE","t","p_value",
                  "lower_2.5","upper_97.5")
  rownames(res) <- NULL
  
  res[sapply(res,is.numeric)] <- lapply(res[sapply(res,is.numeric)],round,4)
  
  return(res)
}

enambleTable_xsecModels <- function(modelsList){
  
  # Get params
  params <- lapply(modelsList,FUN=get_XsecModelParams)
  
  # Ensamble
  res <- do.call(rbind,params)
  return(res)
}
```
