## Supplementary material for "Genetic drivers of hippocampal atrophy highlight the role of *APOE* functional variants and AD polygenicity in Mild Cognitive Impairment": modelling_crossSectional_baselineVolumes_all.R

```
###############################################################################
### MODELLING                                                                 #
###   Cross sectional (baseline) analyses                                     #
###                                                                           #
###############################################################################
rm(list=ls())
source("Rscripts/libraries.R")
source(here(RscriptsDir,"3_modelling","modelling_auxFunctions.R"))

#·························································
load(here(cleandataDir,"dataForAnalysis_mixed.RData"))
paramT <- "All"

# Error in brain measurements
basal <- basal[!basal$PTID%in%c("4337","0558"),]

#·························································
#===============================================================================
# 1) Establish differences between diagnostic groups or 
# biological sex in hippocampal volumes at baseline, 
# adjusting for age, sex/group, and total intracranial volume.
#===============================================================================
volVars <- sort(names(basal)[27:39])
PRSVars <- names(basal)[7:12]
RiskVars <- names(basal)[40:45]
snpVars <- names(basal)[13:23]
basal$tesla <- as.factor(basal$tesla)

# Select subset to analyse
# volVars <- volVars[volVars %in% c("Whole_hippocampus","subiculum","presubiculum",
#                                   "parasubiculum")]
volVars <- c("Whole_hippocampus","subiculum","presubiculum","parasubiculum")
PRSVars <- PRSVars[grep("_AD",PRSVars)]
RiskVars <- RiskVars[grep("^AD",RiskVars)]
snpVars <- snpVars

#------------------------------------------------------------------------------
# FUNCTION adjustedGroupDifferences 
#   INPUT:
#     - volume: variablE hippocamp volume
#
#   TASKS
#     - A) Adjust a linear model to explain hippocampal volume at baseline 
#          based on AGE, ICV, and PTEDUCAT, including an interaction 
#          between DX.bl and PTGENDER.
#     - B) Extract significance of volume differences by diagnostic group, 
#          storing the result in the object 'res_signif'.
#     - C) Calculate marginal means based on DX.bl and PTGENDER, 
#          fixing AGE, ICV, and PTEDUCAT at their mean values.
#     - D) Pairwise contrasts: differences between PTGENDER within each 
#          diagnostic group. Store the result in the object 'contrast'
#     - E) Present results of marginal means for volume in the 
#          object 'res_emmeans'.
#     - F) Generate a diagnostic plot of residuals and save it to the directory.
#
#   OUTPUT
#     - List with 3 objects: res_signif, contrast, res_emmeans
#------------------------------------------------------------------------------
adjustedGroupDifferences <- function(volume){
  
  # Lineal Model including interaction term sex-diagnosis
  expr <- paste(volume,"AGE + ICV + PTEDUCAT + PTGENDER * DX.bl + tesla",
                sep = " ~ ")
  mod <- lm(formula = expr, data = basal)
  smod <- summary(mod)
  anovaTab <- anova(mod)

  pvals_DXbl <- c(anovaTab["DX.bl","Pr(>F)"])
  pvals_DXbl_formatted <- ifelse(pvals_DXbl<0.001,"<0.001",
                                 sprintf("%.3f",pvals_DXbl))
  
  res_signif <- data.frame(Volume = volume,
                           Group = "DX.bl",
                           pval = pvals_DXbl_formatted,
                           test = "FTest")
  
  
  # Marginal means
  RG <- ref_grid(mod,at = list(tesla ="3"))
  
  emm_conjunt <- emmeans(RG,spec=~PTGENDER|DX.bl) # with interaction
  emm_conjuntDF <- as.data.frame(emm_conjunt)
  
  # Pairwaise contrasts: for each DX.bl level, are the sex difference 
  contrast <- as.data.frame(pairs(emm_conjunt))
  contrast <- data.frame(Volume=rep(volume,3),
                         DX.bl = contrast$DX.bl,
                         group1 = rep("Female",3),
                         group2 = rep("Male",3),
                         statistic = contrast$t.ratio,
                         df = contrast$df,
                         p.adj = contrast$p.value)
  
  res_emmeans <- rbind(expand.grid(levels(basal$PTGENDER),levels(basal$DX.bl)))
  res_emmeans <- cbind(rep(volume,nrow(res_emmeans)),
                       res_emmeans)
  
  res_emmeans[c("emmean","lwr","upr")] <- 
    emm_conjuntDF[,c("emmean","lower.CL","upper.CL")]

  res_emmeans$value <- with(res_emmeans,
                            sprintf("%.f [%.f;%.f]",emmean,lwr,upr))
  
  colnames(res_emmeans)[1:3] <- c("Volume","PTGENDER","DX.bl")
  
  
  # Diagnostic residual plots
  file <- paste(volume,"diagnosticPlots.png",sep="_")
  path <- here("results",paramT,"2_Modelling",
               "1_baselineVolumes_groupAdjustedDifferences",
               "residualDiagnosticPlots")
  obj_is_vector = function(x){TRUE}
  
  if(!(any(grepl(file,list.files(path))))){
    resid_panel(mod,qqbands = T,plots = "all")
    ggsave(file=file, path=path ,width = 8, height = 6)
  }
  
  return(list(significance = res_signif , 
              marginal_means = res_emmeans,
              contrast = contrast))
}
#------------------------------------------------------------------------------

#-----------------------------------------------------------------------------
# a) Apply adjustedGroupDifferences() function - all volumes
#-----------------------------------------------------------------------------
res <- lapply(volVars, FUN=adjustedGroupDifferences)

# Extract objects: marginal means, significance between diagnostic groups, 
# and sex differences within each diagnostic group.
res_significance <- do.call(rbind,lapply(res,"[[",1))
res_emmeans <- do.call(rbind,lapply(res,"[[",2))
res_contrast <- do.call(rbind,lapply(res,"[[",3))


#-----------------------------------------------------------------------------
# b) Generate a results table: marginal means of volumes by DX.bl 
#  and PTGENDER, with AGE, ICV, and PTEDUCAT fixed.
#-----------------------------------------------------------------------------

# Generate result tables
emmeansData <- subset(res_emmeans,select=c(1,2,3,7))
emmeansData$class <- with(emmeansData, paste(DX.bl,PTGENDER,sep="_"))
emmeansData <- emmeansData[,c("Volume","class","value")]

# Contrasts of sex differences within each diagnostic groups
cont <- split(res_contrast,res_contrast$DX.bl)
cont <- lapply(cont, "[[","p.adj")
cont <- lapply(cont,function(x)ifelse(x<0.001,"<0.001",sprintf("%.3f",x)))

taula <- reshape(emmeansData, direction = "wide", idvar = "Volume",
                 timevar = "class")
taula[paste(c("CN","MCI","AD"),"sexContrast",sep="_")] <- cont

taulaDef <- taula[c(1:3,8,4,5,9,6,7,10)]
pvalData <- subset(res_significance,Group=="DX.bl",select=c(Volume,pval))
taulaDef <- merge(taulaDef,pvalData,by="Volume",sort = F)

#Sample size
x <- with(basal,table(PTGENDER,DX.bl))
n <- sprintf("N = %.i",x)

# HTML format
tab <- kbl(taulaDef,align = "lcccccccccr",
           col.names = c("",n[1:2],"",n[3:4],"",n[5:6],"",""),
           caption = "Adjusted marginal means for hippocampal volumes at baseline
           by diagnostic group") 

tab <- add_header_above(tab,c("",
                              rep(c(levels(basal$PTGENDER),"P"),3),"Pval_groups"),
                        line = F,bold=T)

tab <- add_header_above(tab,c(" " = 1,"CN"=3,"MCI"=3,"AD"=3,""),
                        bold = T,line = T,)

#tab <- column_spec(tab,2:10,width = "2in")
tab <- kable_classic(tab,full_width=F,font="Cambria")

formattedTab <- footnote(tab,
                         general = c("Results are reported as estimated marginal mean volume and 95% confidence interval by diagnostic group and sex, adjusted by Age, Intracraneal total volume and tesla parameter.",
                                     "Differences between diagnostic groups were tested using F-test for global significance. P-values are reported as Pval_groups.",
                                     "Differences between sex within each diagnostic group were tested pairwise t-test. P-values are reported as P in every diagnostic group",
                                     "Diagnostic groups: Controls (CN), Mild Cognitive Impairment (MCI) and Alzheimer's Disease (AD) "),
                         title_format = c("bold")) 

file <- "result_emmeansVolume.html"
path <- here("results",paramT,"2_Modelling",
             "1_baselineVolumes_groupAdjustedDifferences")

if(!(any(grepl(file,list.files(path))))){
  save_kable(formattedTab,file=here(path,file),zoom=2)
}

formattedTab

#-----------------------------------------------------------------------------
# c) Graph: Hippocampal volumes by DX.bl and PTGENDER, adjusted for AGE, ICV, 
# and PTEDUCAT.
# Marginal means calculated at the mean value of AGE, the mean value of ICV, 
# and the median of PTEDUCAT.
#-----------------------------------------------------------------------------

emmeansData <- subset(res_emmeans,Volume != "Whole_hippocampus",
                      select=-value)

res_contrast$p.adj.signif <- cut(res_contrast$p.adj,
                                 breaks=c(-Inf,0.001,0.01,0.05,0.1,1),
                                 labels=c("***","**","*","·",""))

emmeansData <- merge(emmeansData,
                     res_contrast[,c("Volume","DX.bl","group1","p.adj.signif")],
                     by.x=c("Volume","DX.bl","PTGENDER"),
                     by.y=c("Volume","DX.bl","group1"),all.x=T)

emmeansData$p.adj.signif[is.na(emmeansData$p.adj.signif)] <- ""

pos_dodge <- position_dodge(width=0.2)

ggplot(emmeansData, aes(x=DX.bl)) + 
  geom_pointrange(aes(y=emmean,ymin=lwr,ymax=upr,color=PTGENDER),
                  position = pos_dodge,show.legend = FALSE,size=0.3) +
  geom_text(aes(y=upr,label=p.adj.signif),vjust=-1) +
  scale_y_continuous(expand = expansion(mult = c(0,0.2))) + 
  scale_color_manual(values = c("#D95F02","#7570B3")) +
  facet_wrap(~Volume,scales = "free_y") + 
  ggstatsplot::theme_ggstatsplot() +
  theme(strip.background = element_blank(),
        plot.margin = margin(1,1,.5,1,"cm"),
        axis.title = element_blank()) +
  
  ggtitle("Estimated volume means at baseline by biological sex and diagnostic group.") +
  labs(subtitle = "Adjusted for Age at baseline, sex, intracranial volume, diagnostic status, years of education and tesla parameter. <br/>
       <span style = 'color: #D95F02;'>**Female**</span> and <span style = 'color: #7570B3;'>**Male**</span>" ,
       caption="**Source**: ADNI project | **Units** Volume measurements in mm3 | Differences between sex are reported as significance level of t-test with Tukey's correction, sliced by diagnostic group, <br/>
       at levels \\*** <.001, \\** <.01, \\* <.05 and ·<.1. Non-significant differences are not shown.") +
  theme(plot.caption = element_markdown(hjust = 0, lineheight = 1.5),
        plot.subtitle = element_markdown(size = 12, hjust = 0),
        plot.title = element_text(size = 16, hjust = -.05))

ggsave(file="emmeansAdjustedVolumes.png",
       path = here("results",paramT,"2_Modelling",
                   "1_baselineVolumes_groupAdjustedDifferences"),
       width = 25,height = 15,units = "cm")


#==============================================================================
# 2) ASSOCIATION ANALYSIS BETWEEN PRS AND HIPPOCAMPAL VOLUMES
#==============================================================================

#------------------------------------------------------------------------------
# FUNCTION: effectPRS_strat 
#   INPUT:
#     - Hippocampal volume
#     - PRS
#     - group: "all", or stratified by "CN","MCI","AD"
#
#   TASKS
#     - A) Subset the data by group, if aplicable
#     - B) Fit the additive linear model with covariates and PRS
#     - C) Retrieve the effect of PRS on volume, confidence interval and p-value
#
#   OUTPUT
#     - Dataframe with 4 variables: Effect, lower.ci, upper.ci and Significance
#       corresponding to the effect of PRS on volume for each "group".
#------------------------------------------------------------------------------

effectPRS_strat <- function(volume,PRS,group="All"){
  data <- basal
  covars <- c("AGE","ICV","PTEDUCAT","PTGENDER","DX.bl","tesla")

    if(group!="All"){
    data <- subset(basal,DX.bl == group)
    covars <- covars[covars!="DX.bl"]
    }
  
  # Model
  covars <- paste(covars,collapse = " + ")
  expr <- paste(volume,
                paste(c(covars,PRS), collapse=" + "),
                sep = " ~ ")
  
  # Model: fit, summary and confidence interval
  mod <- lm(formula = expr, data = data)        
  smod <- summary(mod)$coefficients
  CIs <- confint.default(mod)
  df <- summary(mod)$df[2]
  
  pos_beta <- grep(PRS,rownames(smod))
  beta_PRS <- smod[pos_beta,]
  CI_PRS <- CIs[pos_beta,,drop=T]
  res <- data.frame(Effect = beta_PRS[1],
                    SE = beta_PRS[2],
                    df = df,
                    t.ratio = beta_PRS[3],
                    lower.ci = CI_PRS[1],
                    upper.ci = CI_PRS[2],
                    pvalue = beta_PRS[4])

  
  
  rownames(res) <- expr
  attributes(mod)$PRS <- PRS
  attributes(mod)$volume <- volume
  return(list("PRSEffect" = res,
              "ModelGlobal" = mod))
}
#------------------------------------------------------------------------------


#---------------------------------------------------------------------------
# a) Apply effectPRS_strat() function on "modGrid" matrix
#---------------------------------------------------------------------------

modGrid <- expand.grid(PRS = c(PRSVars,snpVars),
                       Volume = volVars,
                       DX.bl = c("All",levels(basal$DX.bl)),
                       stringsAsFactors = F)

modGrid$analysis[modGrid$PRS %in% c(PRSVars,"APOE4")] <- "Primary"
modGrid$analysis[modGrid$PRS %in% snpVars[-1]] <- "Secondary"

# modGrid$analysis[modGrid$PRS %in% PRSVars[1:4]] <- "Primary"
# modGrid$analysis[modGrid$PRS %in% c(PRSVars[5:6],"APOE4")] <- "Secondary"
# modGrid$analysis[modGrid$PRS %in% snpVars[2:7]] <- "Univariate.brain"
# modGrid$analysis[modGrid$PRS %in% snpVars[8:11]] <- "Univariate.CSF"

auxQ1 <- mapply(FUN = effectPRS_strat,
                volume = modGrid$Volume,
                PRS = modGrid$PRS,
                group = modGrid$DX.bl,
                SIMPLIFY = F,USE.NAMES = F)
names(auxQ1) <- sapply(auxQ1,function(x)
  paste(attributes(x$ModelGlobal)$PRS,
         attributes(x$ModelGlobal)$volume,
         sep="|")
)

result_strat <- lapply(auxQ1,"[[","PRSEffect")
result_strat <- do.call(rbind,result_strat)

modGrid <- cbind(modGrid,result_strat)
rownames(modGrid) <- NULL

modGrid$PRS <- factor(modGrid$PRS,levels=c(PRSVars,snpVars))
modGrid$DX.bl <- factor(modGrid$DX.bl, levels=c("All","CN","MCI","AD"))

# modGrid$strat <- modGrid$DX.bl
# levels(modGrid$strat) <- c("All",rep("strat",3))
# res <- split(modGrid,modGrid$strat)
res <- split(modGrid,modGrid$DX.bl)


lr <- lapply(res, function(l){
  l$prsVol <- paste(l$PRS,l$Volume,sep="|")
  aux <- lapply(split(l,l$analysis), function(x){
    x$pvalue_corrected <- p.adjust(x$pvalue, method = "fdr")
    return(x[c("prsVol","pvalue_corrected")])
  })
  aux <- do.call(rbind,aux)
  pos <- match(l$prsVol,aux$prsVol)
  l$pvalue_corrected <- aux[pos,"pvalue_corrected"]
  l$prsVol <- NULL
  return(l)
})
res <- do.call(rbind,lr)
#res$strat <- NULL

res[paste(names(res)[11:12],"symbol",sep="_")] <-
  lapply(res[names(res[11:12])], FUN=cut,
         breaks = c(-Inf,0.001,0.01,0.05,0.1,1),
         labels=c("***","**","*","·",""))
 
significant_effects <- subset(res, pvalue_corrected_symbol!="")


#-----------------------------------------------------------------------------
# b) Q1: Associació between genetic predisposition and volumetric differences?
#-----------------------------------------------------------------------------

# plotData <- subset(res,Volume!="Whole_hippocampus" & DX.bl=="All")
# plotData <- subset(res,& DX.bl=="All")
# names(plotData)[3] <- "Group"
plotData <- subset(res,DX.bl=="All")
names(plotData)[3] <- "Group"


heatmapT1 <- ggplot(data=plotData,aes(x=Volume,y=PRS,fill=Effect)) + 
  geom_tile() +
  geom_text(aes(label=pvalue_corrected_symbol),size=6)+
  scale_fill_gradient2(low="red4",mid = "white", high="#0066FF", midpoint = 0)+
  facet_wrap(~Group)+
  ggstatsplot::theme_ggstatsplot() +
  theme(strip.background = element_blank(),
        plot.margin = margin(1,1,.5,1,"cm"),
        axis.title = element_blank(),
        axis.text = element_text(face="bold",size = 8),
        axis.text.x = element_text(angle=90,vjust = 0.6,hjust = 1)) +
  
  ggtitle("Corrected significant associations between PRS or individual SNPs and
          hippocampal subfield volumes") +
  labs(caption="PRS linear effects on hippocampal volumes, which corresponds to beta1 coefficient of T.1 models. Pvalues are reported as \\*** <.01\\** <.01, \\* <.05 and<br/>· <.1.
  after correcting for multiple comparisons. Effects adjusted for Age at baseline, intracranial volume, sex, diagnostic status, years of education and<br/>
  tesla scan parameter. **Source**: ADNI project") +
  theme(plot.caption = element_markdown(hjust = 0, lineheight = 1.5),
        plot.subtitle = element_markdown(size = 12, hjust = 0),
        plot.title = element_text(size = 16, hjust = 0))


heatmapT1
subset(res, pvalue_symbol !="" & DX.bl=="All",
       c(Volume,PRS,Effect,pvalue_symbol,pvalue_corrected_symbol))


### All+Groups: primary analysis
plotData <- subset(res,analysis=="Primary")
names(plotData)[3] <- "Group"

upper.th <- quantile(plotData$Effect,1)
lower.th <- quantile(plotData$Effect,0)
plotData$Volume <- factor(plotData$Volume,levels=unique(plotData$Volume),
                          labels=c("Whole\nhippocampus","Subiculum","Pre-\nsubiculum","Para-\nsubiculum"))


heatmap <- ggplot(data=plotData,aes(x=Volume,y=PRS,
                                    fill=pmax(pmin(Effect,upper.th),lower.th))) + 
  geom_tile(color="grey") +
  geom_text(aes(label=pvalue_corrected_symbol),size=4)+
  scale_fill_gradient2(low="red4",mid = "white", high="#0066FF", midpoint = 0,
                       name="Effect")+
  facet_wrap(~Group)+
  ggstatsplot::theme_ggstatsplot() +
  theme(strip.background = element_blank(),
        plot.margin = margin(1,1,.5,1,"cm"),
        axis.title = element_blank(),
        axis.text = element_text(face="bold",size = 8)) +
  
  # ggtitle("Corrected significant associations between PRS or individual SNPs and
  #         hippocampal subfield volumes") +
  labs(caption="Genetic predictor linear main effects on hippocampal volumes. Pvalues are reported as \\*** <.01\\** <.01, \\* <.05 and ·<.1.<br/>
  after correcting for multiple comparisons. Effects adjusted for Age at baseline, intracranial volume, sex, diagnostic status,<br/>
       years of education and tesla scan parameter.") +
  theme(plot.caption = element_markdown(hjust = 0, lineheight = 1.5),
        plot.subtitle = element_markdown(size = 12, hjust = 0),
        plot.title = element_text(size = 16, hjust = 0))

heatmap.primary <- heatmap
heatmap.primary
#8.68x5.45 in  export pdf


### All: secondary analysis
plotData <- subset(res,analysis=="Secondary")
names(plotData)[3] <- "Group"

upper.th <- quantile(plotData$Effect,1)
lower.th <- quantile(plotData$Effect,0)
plotData$Volume <- factor(plotData$Volume,levels=unique(plotData$Volume),
                          labels=c("Whole\nhippocampus","Subiculum","Pre-\nsubiculum","Para-\nsubiculum"))


heatmap <- ggplot(data=plotData,aes(x=Volume,y=PRS,
                                    fill=pmax(pmin(Effect,upper.th),lower.th))) + 
  geom_tile(color="grey") +
  geom_text(aes(label=pvalue_corrected_symbol),size=4)+
  scale_fill_gradient2(low="red4",mid = "white", high="#0066FF", midpoint = 0,
                       name="beta")+
  facet_wrap(~Group)+
  ggstatsplot::theme_ggstatsplot() +
  theme(strip.background = element_blank(),
        plot.margin = margin(1,1,.5,1,"cm"),
        axis.title = element_blank(),
        axis.text = element_text(face="bold",size = 8)) +
  
  # ggtitle("Corrected significant associations between PRS or individual SNPs and
  #         hippocampal subfield volumes") +
  labs(caption="Genetic predictor linear main effects on hippocampal volumes. Pvalues are reported as \\*** <.01\\** <.01, \\* <.05 and ·<.1.<br/>
  after correcting for multiple comparisons. Effects adjusted for Age at baseline, intracranial volume, sex, diagnostic status,<br/>
       years of education and tesla scan parameter.") +
  theme(plot.caption = element_markdown(hjust = 0, lineheight = 1.5),
        plot.subtitle = element_markdown(size = 12, hjust = 0),
        plot.title = element_text(size = 16, hjust = 0))

heatmap.secondary <- heatmap
heatmap.secondary
#8.68x6 in  export pdf


### All + groups, primary analysis, only Whole hippocampus
plotData <- subset(res,analysis=="Primary" & Volume == "Whole_hippocampus")
names(plotData)[3] <- "Group"

upper.th <- quantile(plotData$Effect,1)
lower.th <- quantile(plotData$Effect,0)


heatmap <- ggplot(data=plotData,aes(x=Group,y=PRS,
                                    fill=pmax(pmin(Effect,upper.th),lower.th))) + 
  geom_tile(color="grey") +
  geom_text(aes(label=pvalue_corrected_symbol),size=4)+
  scale_fill_gradient2(low="red4",mid = "white", high="#0066FF", midpoint = 0,
                       name="Effect")+
  ggstatsplot::theme_ggstatsplot() +
  theme(strip.background = element_blank(),
        plot.margin = margin(1,1,.5,1,"cm"),
        axis.title = element_blank(),
        axis.text = element_text(face="bold",size = 8)) +
  
  # ggtitle("Corrected significant associations between PRS or individual SNPs and
  #         hippocampal subfield volumes") +
  # labs(caption="Genetic predictor linear main effects on hippocampal volumes. Pvalues are reported as \\*** <.01\\** <.01, \\* <.05 and ·<.1.<br/>
  # after correcting for multiple comparisons. Effects adjusted for Age at baseline, intracranial volume, sex, diagnostic status,<br/>
  #      years of education and tesla scan parameter.") +
  theme(plot.caption = element_markdown(hjust = 0, lineheight = 1.5),
        plot.subtitle = element_markdown(size = 12, hjust = 0),
        plot.title = element_text(size = 16, hjust = 0))

heatmap.primary.WH <- heatmap
heatmap.primary.WH


#-----------------------------------------------------------------------------
# b) Q2: Does the PRS effect on hippocampal volume differences at baseline 
#    depend on the diagnostic status of individuals?
#     Contrast: All interaction terms PRS:DXbl are equal to 0
#-----------------------------------------------------------------------------
volume <- "CA1"; PRS <- "AD_predisposition"; interact <- "DX.bl"

models_interact <- function(volume,PRS,interact){
  data <- basal
  covars <- c("AGE","ICV","PTEDUCAT","PTGENDER","DX.bl","tesla")
  
  covars <- paste(covars <- covars[covars!=interact],collapse = " + ")
  prs_interact <- paste(PRS,interact,sep = " * ")
  expr <- paste(volume,
                paste(c(covars,prs_interact), collapse=" + "),
                sep = " ~ ")
  
  mod <- lm(formula = expr, data = data)
  attributes(mod)$PRS <- PRS
  attributes(mod)$volume <- volume
  
  specs <- as.formula(paste0("~",PRS,"|",interact))
  trend <- emtrends(mod, specs = specs, var=PRS, infer=c(F,T))
  
  attributes(trend)$PRS <- PRS
  attributes(trend)$volume <- volume
  
  return(list("PRSEffect" = trend,
              "modDXInteract" = mod))
}

modGridQ2 <- subset(modGrid,DX.bl=="All",select=c(PRS,Volume,analysis))

#x <- models_interact("Whole_hippocampus","PRS_AD",interact="DX.bl")

auxQ2 <- mapply(FUN = models_interact,
                      volume = as.character(modGridQ2$Volume),
                      PRS = as.character(modGridQ2$PRS),
                      MoreArgs = list(interact="DX.bl"),
                      SIMPLIFY = F)

betaPRS_DXInt <- lapply(auxQ2,"[[","PRSEffect")
names(betaPRS_DXInt) <- paste(modGridQ2$Volume,modGridQ2$PRS,
                              sep=" ~ ") 

aux <- lapply(betaPRS_DXInt, function(x){
  df <- summary(x)
  #names(df)[3] <- "PRS_effect"
  betas <- df[,c(2:7)]
  names(betas)[c(2,6)] <- c("Effect","pvalue")
  
  n <- nrow(betas)
  res <- data.frame(Volume = rep(attributes(x)$volume, times = n),
                    PRS = rep(attributes(x)$PRS, time = n))

  res <- cbind(res,betas)
  return(res)
})

resultQ2 <- do.call(rbind,aux)
resultQ2$PRS <- factor(resultQ2$PRS,levels=c(PRSVars,snpVars))
resultQ2$DX.bl <- factor(resultQ2$DX.bl, levels=c("CN","MCI","AD"))
resultQ2$analysis <- modGridQ2$analysis[match(resultQ2$PRS,modGrid$PRS)]

resultQ2$prsVolDX <- paste(resultQ2$PRS,resultQ2$Volume,resultQ2$DX.bl,sep="|")
aux <- lapply(split(resultQ2,resultQ2$analysis), function(x){
  x$pvalue_corrected <- p.adjust(x$pvalue, method = "fdr")
  return(x[c("prsVolDX","pvalue_corrected")])
})
aux <- do.call(rbind,aux)
pos <- match(resultQ2$prsVolDX,aux$prsVolDX)
resultQ2$pvalue_corrected <- aux[pos,"pvalue_corrected"]
resultQ2$prsVolDX <- NULL

resultQ2[paste(names(resultQ2)[c(8,10)],"symbol",sep="_")] <-
  lapply(resultQ2[names(resultQ2[c(8,10)])], FUN=cut,
         breaks = c(-Inf,0.001,0.01,0.05,0.1,1),
         labels=c("***","**","*","·",""))

resultQ2.withAll <- resultQ2
aux.all <- res[res$DX.bl=="All",names(resultQ2.withAll)]
resultQ2.withAll <- rbind(resultQ2.withAll,aux.all)
resultQ2.withAll$DX.bl <- relevel(resultQ2.withAll$DX.bl,ref="All")
names(resultQ2.withAll)[3] <- "Group"

# Primary
plotData <- resultQ2.withAll[resultQ2.withAll$analysis=="Primary",]
upper.th <- quantile(plotData$Effect,1)
lower.th <- quantile(plotData$Effect,0)
plotData$Volume <- factor(plotData$Volume,levels=unique(plotData$Volume),
                          labels=c("Whole\nhippocampus","Subiculum","Pre-\nsubiculum","Para-\nsubiculum"))


heatmap <- ggplot(data=plotData,aes(x=Volume,y=PRS,
                                    fill=pmax(pmin(Effect,upper.th),lower.th))) + 
  geom_tile(color="grey") +
  geom_text(aes(label=pvalue_corrected_symbol),size=5)+
  scale_fill_gradient2(low="red4",mid = "white", high="#0066FF", midpoint = 0,
                       name="Effect")+
  facet_wrap(~Group)+
  ggstatsplot::theme_ggstatsplot() +
  theme(strip.background = element_blank(),
        plot.margin = margin(1,1,.5,1,"cm"),
        axis.title = element_blank(),
        axis.text = element_text(face="bold",size = 8)) +
  
  # ggtitle("Corrected significant associations between PRS or individual SNPs and
  #         hippocampal subfield volumes") +
  labs(caption="Genetic predictor linear main effects on hippocampal volumes. Pvalues are reported as \\*** <.01\\** <.01, \\* <.05 and ·<.1.<br/>
  after correcting for multiple comparisons. Effects adjusted for Age at baseline, intracranial volume, sex, diagnostic status,<br/>
       years of education and tesla scan parameter.") +
  theme(plot.caption = element_markdown(hjust = 0, lineheight = 1.5),
        plot.subtitle = element_markdown(size = 12, hjust = 0),
        plot.title = element_text(size = 16, hjust = 0))

heatmap.primary <- heatmap
heatmap.primary
#8.68x5.45 in  export pdf

### All: secondary analysis
plotData <- resultQ2.withAll[resultQ2.withAll$analysis=="Secondary",]
upper.th <- quantile(plotData$Effect,1)
lower.th <- quantile(plotData$Effect,0)
plotData$Volume <- factor(plotData$Volume,levels=unique(plotData$Volume),
                          labels=c("Whole\nhippocampus","Subiculum","Pre-\nsubiculum","Para-\nsubiculum"))


heatmap <- ggplot(data=plotData,aes(x=Volume,y=PRS,
                                    fill=pmax(pmin(Effect,upper.th),lower.th))) + 
  geom_tile(color="grey") +
  geom_text(aes(label=pvalue_corrected_symbol),size=5)+
  scale_fill_gradient2(low="red4",mid = "white", high="#0066FF", midpoint = 0,
                       name="Effect")+
  facet_wrap(~Group)+
  ggstatsplot::theme_ggstatsplot() +
  theme(strip.background = element_blank(),
        plot.margin = margin(1,1,.5,1,"cm"),
        axis.title = element_blank(),
        axis.text = element_text(face="bold",size = 8)) +
  
  # ggtitle("Corrected significant associations between PRS or individual SNPs and
  #         hippocampal subfield volumes") +
  labs(caption="Genetic predictor linear main effects on hippocampal volumes. Pvalues are reported as \\*** <.01\\** <.01, \\* <.05 and ·<.1.<br/>
  after correcting for multiple comparisons. Effects adjusted for Age at baseline, intracranial volume, sex, diagnostic status,<br/>
       years of education and tesla scan parameter.") +
  theme(plot.caption = element_markdown(hjust = 0, lineheight = 1.5),
        plot.subtitle = element_markdown(size = 12, hjust = 0),
        plot.title = element_text(size = 16, hjust = 0))

heatmap.secondary <- heatmap
heatmap.secondary
#8.68x6 in  export pdf


# Primary Whole hippocampus
plotData <- resultQ2.withAll[resultQ2.withAll$analysis=="Primary" &
                             resultQ2.withAll$Volume=="Whole_hippocampus",]
upper.th <- quantile(plotData$Effect,1)
lower.th <- quantile(plotData$Effect,0)


heatmap <- ggplot(data=plotData,aes(x=Group,y=PRS,
                                    fill=pmax(pmin(Effect,upper.th),lower.th))) + 
  geom_tile(color="grey") +
  geom_text(aes(label=pvalue_corrected_symbol),size=5)+
  scale_fill_gradient2(low="red4",mid = "white", high="#0066FF", midpoint = 0,
                       name="Effect")+
  ggstatsplot::theme_ggstatsplot() +
  theme(strip.background = element_blank(),
        plot.margin = margin(1,1,.5,1,"cm"),
        axis.title = element_blank(),
        axis.text = element_text(face="bold",size = 8)) +
  
  # ggtitle("Corrected significant associations between PRS or individual SNPs and
  #         hippocampal subfield volumes") +
  # labs(caption="Genetic predictor linear main effects on hippocampal volumes. Pvalues are reported as \\*** <.01\\** <.01, \\* <.05 and ·<.1.<br/>
  # after correcting for multiple comparisons. Effects adjusted for Age at baseline, intracranial volume, sex, diagnostic status,<br/>
  #      years of education and tesla scan parameter.") +
  theme(plot.caption = element_markdown(hjust = 0, lineheight = 1.5),
        plot.subtitle = element_markdown(size = 12, hjust = 0),
        plot.title = element_text(size = 16, hjust = 0))

heatmap.primary.WH <- heatmap
heatmap.primary.WH

# PDF 7ix3i


### Secondary analysis - Whole hippocampus
plotData <- resultQ2.withAll[resultQ2.withAll$analysis=="Secondary" & 
                             resultQ2.withAll$Volume=="Whole_hippocampus",]
upper.th <- quantile(plotData$Effect,1)
lower.th <- quantile(plotData$Effect,0)


heatmap <- ggplot(data=plotData,aes(x=Group,y=PRS,
                                    fill=pmax(pmin(Effect,upper.th),lower.th))) + 
  geom_tile(color="grey") +
  geom_text(aes(label=pvalue_corrected_symbol),size=5)+
  scale_fill_gradient2(low="red4",mid = "white", high="#0066FF", midpoint = 0,
                       name="Effect")+
  ggstatsplot::theme_ggstatsplot() +
  theme(strip.background = element_blank(),
        plot.margin = margin(1,1,.5,1,"cm"),
        axis.title = element_blank(),
        axis.text = element_text(face="bold",size = 8)) +
  
  # ggtitle("Corrected significant associations between PRS or individual SNPs and
  #         hippocampal subfield volumes") +
  # labs(caption="Genetic predictor linear main effects on hippocampal volumes. Pvalues are reported as \\*** <.01\\** <.01, \\* <.05 and ·<.1.<br/>
  # after correcting for multiple comparisons. Effects adjusted for Age at baseline, intracranial volume, sex, diagnostic status,<br/>
  #      years of education and tesla scan parameter.") +
  theme(plot.caption = element_markdown(hjust = 0, lineheight = 1.5),
        plot.subtitle = element_markdown(size = 12, hjust = 0),
        plot.title = element_text(size = 16, hjust = 0))

heatmap.secondary.WH <- heatmap
heatmap.secondary.WH


# Primary Subfields (subiculum region)
plotData <- resultQ2.withAll[resultQ2.withAll$analysis=="Primary" &
                               resultQ2.withAll$Volume!="Whole_hippocampus",]
upper.th <- quantile(plotData$Effect,1)
lower.th <- quantile(plotData$Effect,0)
plotData$Volume <- factor(plotData$Volume,levels=unique(plotData$Volume),
                          labels=c("Subiculum","Pre-\nsubiculum","Para-\nsubiculum"))


heatmap <- ggplot(data=plotData,aes(x=Volume,y=PRS,
                                    fill=pmax(pmin(Effect,upper.th),lower.th))) + 
  geom_tile(color="grey") +
  geom_text(aes(label=pvalue_corrected_symbol),size=5)+
  scale_fill_gradient2(low="red4",mid = "white", high="#0066FF", midpoint = 0,
                       name="Effect")+
  facet_wrap(~Group)+
  ggstatsplot::theme_ggstatsplot() +
  theme(strip.background = element_blank(),
        plot.margin = margin(1,1,.5,1,"cm"),
        axis.title = element_blank(),
        axis.text = element_text(face="bold",size = 8)) +
  
  # ggtitle("Corrected significant associations between PRS or individual SNPs and
  #         hippocampal subfield volumes") +
  # labs(caption="Genetic predictor linear main effects on hippocampal volumes. Pvalues are reported as \\*** <.01\\** <.01, \\* <.05 and ·<.1.<br/>
  # after correcting for multiple comparisons. Effects adjusted for Age at baseline, intracranial volume, sex, diagnostic status,<br/>
  #      years of education and tesla scan parameter.") +
  theme(plot.caption = element_markdown(hjust = 0, lineheight = 1.5),
        plot.subtitle = element_markdown(size = 12, hjust = 0),
        plot.title = element_text(size = 16, hjust = 0))

heatmap.primary.subfields <- heatmap
heatmap.primary.subfields

#8.7x5.5 in  export pdf

# Secondary Subfields (subiculum region)
plotData <- resultQ2.withAll[resultQ2.withAll$analysis=="Secondary" &
                               resultQ2.withAll$Volume!="Whole_hippocampus",]
upper.th <- quantile(plotData$Effect,1)
lower.th <- quantile(plotData$Effect,0)
plotData$Volume <- factor(plotData$Volume,levels=unique(plotData$Volume),
                          labels=c("Subiculum","Pre-\nsubiculum","Para-\nsubiculum"))


heatmap <- ggplot(data=plotData,aes(x=Volume,y=PRS,
                                    fill=pmax(pmin(Effect,upper.th),lower.th))) + 
  geom_tile(color="grey") +
  geom_text(aes(label=pvalue_corrected_symbol),size=5)+
  scale_fill_gradient2(low="red4",mid = "white", high="#0066FF", midpoint = 0,
                       name="Effect")+
  facet_wrap(~Group)+
  ggstatsplot::theme_ggstatsplot() +
  theme(strip.background = element_blank(),
        plot.margin = margin(1,1,.5,1,"cm"),
        axis.title = element_blank(),
        axis.text = element_text(face="bold",size = 8)) +
  
  # ggtitle("Corrected significant associations between PRS or individual SNPs and
  #         hippocampal subfield volumes") +
  # labs(caption="Genetic predictor linear main effects on hippocampal volumes. Pvalues are reported as \\*** <.01\\** <.01, \\* <.05 and ·<.1.<br/>
  # after correcting for multiple comparisons. Effects adjusted for Age at baseline, intracranial volume, sex, diagnostic status,<br/>
  #      years of education and tesla scan parameter.") +
  theme(plot.caption = element_markdown(hjust = 0, lineheight = 1.5),
        plot.subtitle = element_markdown(size = 12, hjust = 0),
        plot.title = element_text(size = 16, hjust = 0))

heatmap.secondary.subfields <- heatmap
heatmap.secondary.subfields

#8.7x5.5 in  export pdf


#-----------------------------------------------------------------------------
# c) Q3: Quantify the association between genetic predisposition and 
#     hippocampal volume according to diagnostic status
#     Stratified aanalysis
#-----------------------------------------------------------------------------

plotData <- subset(res,Volume!="Whole_hippocampus")
names(plotData)[3] <- "Group"
heatmapT1 %+%plotData +
  labs(caption="PRS linear effects on hippocampal volumes, which corresponds to beta1 coefficient of T.1 models. Pvalues are reported as \\*** <.001, \\** <.01,<br/>
  \\* <.05 and · <.1 after correcting for multiple comparisons. Effects adjusted for Age at baseline,<br/>
  sex, intracranial volume, years of education and tesla scan parameter.<br/>
  **Source**: ADNI project")
  
#-----------------------------------------------------------------------------
# d) Q4.1: Is the association between PRS and volumes, overall by 
# diagnostic group, different by sex?
#-----------------------------------------------------------------------------

# ## GLOBALLY ·······················································
# modGridQ4 <- subset(modGrid,DX.bl=="All",select=c(PRS,Volume))
# 
# #x <- models_interact("Whole_hippocampus","PRS_AD",interact="DX.bl")
# 
# betaPRS_Sex <- mapply(FUN = models_interact,
#                       volume = as.character(modGridQ4$Volume),
#                       PRS = as.character(modGridQ4$PRS),
#                       MoreArgs = list(interact="PTGENDER"),
#                       SIMPLIFY = F)
# betaPRS_Sex <- lapply(betaPRS_Sex,"[[","PRSEffect")
# names(betaPRS_Sex) <- paste(modGridQ4$Volume,modGridQ4$PRS,
#                             sep=" ~ ") 
# 
# 
# 
# aux <- lapply(betaPRS_Sex, function(x){
#   df <- summary(x)
#   betas <- df[,c(2,3,7)]
#   names(betas)[c(2,3)] <- c("Effect","Significance")
#   
#   
#   n <- nrow(betas)
#   res <- data.frame(Volume = rep(attributes(x)$volume, times = n),
#                     PRS = rep(attributes(x)$PRS, time = n))
#   
#   res <- cbind(res,betas)
#   return(res)
# })
# 
# resultQ4 <- do.call(rbind,aux)
# resultQ4$PRS <- factor(resultQ4$PRS,levels=PRSVars)
# resultQ4$PTGENDER <- factor(resultQ4$PTGENDER, levels=c("Female","Male"))
# 
# # aux <- lapply(split(resultQ4,resultQ4$Volume),
#               FUN=function(x)p.adjust(x$Significance,method="fdr"))
# resultQ4$Significance_corrected <- do.call(c,aux)
# 
## resultQ4[paste(names(resultQ4)[5:6],"symbol",sep="_")] <-
#   lapply(resultQ4[names(resultQ4[5:6])], FUN=cut,
#          breaks = c(-Inf,0.001,0.01,0.05,0.1,1),
#          labels=c("***","**","*","·",""))
# 
# 
# plotData <- subset(resultQ4,Volume!="Whole_hippocampus")
# names(plotData)[3] <- "Group"
# heatmapT1 %+%plotData
# subset(resultQ4, Significance_symbol !="",
#        c(Volume,PRS,PTGENDER,Effect,Significance_symbol,Significance_corrected_symbol))


#-----------------------------------------------------------------------------
# d) Q4.2: Is the association between PRS and volumes, stratified by 
#   diagnostic group, different by sex?
#-----------------------------------------------------------------------------

#------------------------------------------------------------------------------
# FUNCTION: effectPRS_strat_inter 
#   INPUT:
#     - Hippocampal volume
#     - PRS
#     - group: "CN","MCI","AD" 
#
#   TASKS
#     - A) Subset the data by group, if applicable.
#     - B) Fit the linear model with PRS interaction with sex, PRS*PTGENDER,
#          adjusting for baseline age (AGE), education (PTEDUCAT), 
#          and total intracranial volume (ICV).
#     - C) Retrieve the effect of PRS on volume and confidence interval, 
#          by sex (PTGENDER) and for the stratified data by DX.bl group.
#          Also return the significance of the Wald test for the 
#          interaction term.
#
#   OUTPUT
#     - Dataframe with 9 variables: 
#         * DX.bl: diagnostic group of the stratiieds model.
#
#         * Volume: hippocampal subfield with baseline volume as the response.
#
#         * PRS: polygenic risk score used as the explanatory variable.
#
#         * PTGENDER: male or female, depending on which 
#            sex the effect refers to.
#
#         * Effect: Effect: PRS effect on the response volume, considering 
#           the interaction with sex (PTGENDER). Calculated as:
#           i) beta_PRS_male = beta_PRS
#           ii) beta_PRS_female = beta_PRS + beta_prs:female
#         * ci_lwr, ci_upr: confidence interval limits for Effect at 95%.
#
#         * Significance: p-value for contrast H0: beta_effect=0. Or:
#             Male: H0: beta_effect = beta_PRS = 0
#             Female: H0: beta_effect = beta_PRS + beta_prs:female = 0
#
#         * pvalInteraction: p-value for significance of the Wald test 
#             on the interaction coefficient beta_prs:female.
#------------------------------------------------------------------------------


effectPRS_strat_inter <- function(volume,PRS,group){
  
  data_strata <- subset(basal,DX.bl == group)
  
  covars <- paste(c("AGE","ICV","PTEDUCAT","tesla","PTGENDER"),
                  collapse=" + ")
  
  
  expr_additiu <- paste(volume,
                        paste(c(covars,PRS), collapse=" + "),
                        sep = " ~ ")
  expr_inter <- paste(volume,
                        paste(c(covars,PRS), collapse=" * "),
                        sep = " ~ ")

  mod_add <- lm(formula = expr_additiu, data = data_strata)        
  mod_int <- lm(formula = expr_inter, data = data_strata)
  
  attributes(mod_int)$PRS <- PRS
  attributes(mod_int)$volume <- volume
  
  prs_pos <- which(names(coef(mod_add)) == PRS)
  mainEffectPRS <- summary(mod_add)$coefficient[prs_pos,c(1:4)]
  ci <- confint.default(mod_add)[prs_pos,]
  df <- summary(mod_add)$df[2]

  emm <- emtrends(mod_int,~PTGENDER,var=PRS)
  emm.df <- as.data.frame(summary(emm,infer=c(T,T)))
  contrast <- as.data.frame(pairs(emm))
  signif <- contrast$p.value <0.05
  
  res <- data.frame(DX.bl=group,
                    Volume = rep(volume,3),
                    PRS = rep(PRS,3),
                    PTGENDER = c("Global","Female","Male"),
                    Effect = c(mainEffectPRS[1],emm.df[,2]),
                    SE = c(mainEffectPRS[2],emm.df$SE),
                    df = c(df,emm.df$df),
                    t.ratio = c(mainEffectPRS[3],emm.df$t.ratio),
                    ci_lwr = c(ci[1],emm.df$lower.CL),
                    ci_upr = c(ci[2],emm.df$upper.CL),
                    Significance = c(mainEffectPRS[4],emm.df$p.value),
                    # pvalInteraction = c(NA,contrast$p.value,NA),
                    subset = as.logical(c(1,0,0) + signif*c(-1,1,1)))
  
  return(list("PRSEffect" = res,
              "modSxInteract" = mod_int))
}
#-----------------------------------------------------------------------------


modGrid_strata <- expand.grid(PRS = c(PRSVars,snpVars),
                              Volume = sort(volVars),
                              Dx.bl = levels(basal$DX.bl),
                              stringsAsFactors = F)

result_strat <- mapply(FUN = effectPRS_strat_inter,
                       volume = modGrid_strata$Volume,
                       PRS = modGrid_strata$PRS,
                       group = modGrid_strata$Dx.bl,
                       SIMPLIFY = F,USE.NAMES = F)

auxQ4 <- lapply(result_strat,"[[","PRSEffect")

auxQ4 <- lapply(auxQ4,FUN=subset, subset=subset)

effectPRS_Sx <- do.call(rbind,auxQ4)
effectPRS_Sx$subset <- NULL

effectPRS_Sx$PRS <- factor(effectPRS_Sx$PRS,
                        levels=c(PRSVars,snpVars))
effectPRS_Sx$DX.bl <- factor(effectPRS_Sx$DX.bl,
                          levels=c("AD","MCI","CN"))
effectPRS_Sx$PTGENDER <- factor(effectPRS_Sx$PTGENDER,
                             levels = c("Global","Female","Male"))

effectPRS_Sx$alpha <- with(effectPRS_Sx,ifelse(Significance<0.05,1,.9))

# iv) Afegir simbols de significació
# effectPRS[paste(c("Significance","Interact"),"symbol",sep="_")] <-
#   lapply(effectPRS[c("Significance","pvalInteraction")],
#          FUN=cut,breaks = c(-Inf,0.001,0.01,0.05,0.1,1),
#          labels=c("***","**","*","^",""))
# effectPRS[is.na(effectPRS$Interact_symbol),"Interact_symbol"] <- ""

head(effectPRS_Sx)

effectPRS_Sx$analysis <- modGrid$analysis[match(effectPRS_Sx$PRS,modGrid$PRS)]


  pos_dodge <- position_dodge(width=0.8) 
  volume.labs <- c("Whole\nhippocampus",
                   "Subiculum",
                   "Pre-\nsubiculum",
                   "Para-\nsubiculum")
  names(volume.labs) <- volVars
  
  PRS.labs <- c(sapply(strsplit(PRSVars,split="_"),"[",2),"APOE4")
  names(PRS.labs) <- c(PRSVars,"APOE4")
  PRS.labs[PRS.labs=="ADnoAPOE"] <- "AD\nno APOE"

plotData <- effectPRS_Sx[effectPRS_Sx$analysis=="Primary",]  
plotData$Volume <- factor(plotData$Volume, levels=volVars)  
ggplot(plotData,aes(x=DX.bl)) +
  geom_hline(yintercept=0,linetype="dashed",color="red")+
  geom_pointrange(aes(y=Effect , ymin=ci_lwr , ymax=ci_upr, 
                      color=PTGENDER, alpha=alpha),
                  position = pos_dodge,show.legend = F) +
#  geom_text(aes(y=Effect,label=Significance_symbol, color =PTGENDER),size=5,
#           position = position_dodge(width=0.1), show.legend = F)+
  facet_grid(rows=vars(PRS),cols=vars(Volume),scales = "free",
             labeller=labeller(Volume=volume.labs,PRS=PRS.labs))+
  scale_color_manual(values = c("#000000","#D95F02","#7570B3")) + 
  scale_alpha(range = c(0.3,1))+
  coord_flip()+
  
  theme_bw() +
  theme(strip.background = element_blank(),
        plot.margin = margin(1,1,.5,1,"cm"),
        axis.title = element_blank(),
        axis.text.x = element_text(angle=90,hjust=1)) +
  ggtitle("Association results for direct effects of polygenic risk scores on hippocampal volumes<br/>for female and male and stratified by diagnostic group.") +
  labs(subtitle = "Adjusted for Age at baseline, sex, intracranial volume, years of education and tesla scan parameter. <br/>
       <span style = 'color: #000000;'>**Global**</span> , <span style = 'color: #D95F02;'>**Female**</span> and <span style = 'color: #7570B3;'>**Male**</span>" ,
       caption="**Source**: ADNI project | **Units** Linear coefficient and 95% confidence intervals of PRS effect, for female and male when sex interaction with<br/>
       PRS is significant; or main effect otherwise in models T.3. Non-significant coeficients are shown with transparency, according to Wald-test<br/>
       p-value at level 0.05.") +
  theme(plot.caption = element_markdown(hjust = 0, lineheight = 1.5),
        plot.subtitle = element_markdown(size = 12, hjust = 0),
        plot.title = element_markdown(size = 16, hjust = 0)) -> primaryPanel

primaryPanel
# Export 9.86x7.58 landscape

PRS.labs <- gsub("\\.","\n",snpVars[2:7])
names(PRS.labs) <- snpVars[2:7]

effectPRS_Sx[grep("eQTL",effectPRS_Sx$PRS),"analysis"] <- "Secondary.eQTL"
plotData <- effectPRS_Sx[effectPRS_Sx$analysis=="Secondary.eQTL",]  
plotData$Volume <- factor(plotData$Volume, levels=volVars)  

ggplot(plotData, aes(x=DX.bl)) +
  geom_hline(yintercept=0,linetype="dashed",color="red")+
  geom_pointrange(aes(y=Effect , ymin=ci_lwr , ymax=ci_upr, 
                      color=PTGENDER, alpha=alpha),
                  position = pos_dodge,show.legend = F) +
  #  geom_text(aes(y=Effect,label=Significance_symbol, color =PTGENDER),size=5,
  #           position = position_dodge(width=0.1), show.legend = F)+
  facet_grid(rows=vars(PRS),cols=vars(Volume),scales = "free",
             labeller=labeller(Volume=volume.labs,PRS=PRS.labs))+
  scale_color_manual(values = c("#000000","#D95F02","#7570B3")) + 
  scale_alpha(range = c(0.3,1))+
  coord_flip()+
  
  theme_bw() +
  theme(strip.background = element_blank(),
        plot.margin = margin(1,1,.5,1,"cm"),
        axis.title = element_blank(),
        axis.text.x = element_text(angle=90,hjust=1)) +
  ggtitle("Association results for direct effects of polygenic risk scores on hippocampal<br/>volumes for female and male and stratified by diagnostic group.") +
  labs(subtitle = "Adjusted for Age at baseline, sex, intracranial volume, years of education and tesla scan parameter. <br/>
       <span style = 'color: #000000;'>**Global**</span> , <span style = 'color: #D95F02;'>**Female**</span> and <span style = 'color: #7570B3;'>**Male**</span>" ,
       caption="**Source**: ADNI project | **Units** Linear coefficient and 95% confidence intervals of PRS effect, for female and male when sex<br/>
       interaction with PRS is significant; or main effect otherwise in models T.3. Non-significant coeficients are shown with<br/>
       transparency, according to Wald-test p-value at level 0.05.") +
  theme(plot.caption = element_markdown(hjust = 0, lineheight = 1.5),
        plot.subtitle = element_markdown(size = 12, hjust = 0),
        plot.title = element_markdown(size = 16, hjust = 0)) -> secondaryPanel.eQTL

# Export 9.86x7.58 portrait
secondaryPanel.eQTL


PRS.labs <- gsub("\\.","\n",snpVars[2:7])
names(PRS.labs) <- snpVars[2:7]

effectPRS_Sx[grep("pQTL",effectPRS_Sx$PRS),"analysis"] <- "Secondary.pQTL"
plotData <- effectPRS_Sx[effectPRS_Sx$analysis=="Secondary.pQTL",]  
plotData$Volume <- factor(plotData$Volume, levels=volVars)  


ggplot(plotData, aes(x=DX.bl)) +
  geom_hline(yintercept=0,linetype="dashed",color="red")+
  geom_pointrange(aes(y=Effect , ymin=ci_lwr , ymax=ci_upr, 
                      color=PTGENDER, alpha=alpha),
                  position = pos_dodge,show.legend = F) +
  #  geom_text(aes(y=Effect,label=Significance_symbol, color =PTGENDER),size=5,
  #           position = position_dodge(width=0.1), show.legend = F)+
  facet_grid(rows=vars(PRS),cols=vars(Volume),scales = "free",
             labeller=labeller(Volume=volume.labs,PRS=PRS.labs))+
  scale_color_manual(values = c("#000000","#D95F02","#7570B3")) + 
  scale_alpha(range = c(0.3,1))+
  coord_flip()+
  
  theme_bw() +
  theme(strip.background = element_blank(),
        plot.margin = margin(1,1,.5,1,"cm"),
        axis.title = element_blank(),
        axis.text.x = element_text(angle=90,hjust=1)) +
  ggtitle("Association results for direct effects of polygenic risk scores on hippocampal<br/>
          volumesfor female and male and stratified by diagnostic group.") +
  labs(subtitle = "Adjusted for Age at baseline, sex, intracranial volume, years of education and tesla scan parameter. <br/>
       <span style = 'color: #000000;'>**Global**</span> , <span style = 'color: #D95F02;'>**Female**</span> and <span style = 'color: #7570B3;'>**Male**</span>" ,
       caption="**Source**: ADNI project | **Units** Linear coefficient and 95% confidence intervals of PRS effect, for female and male when sex<br/>
       interaction with PRS is significant; or main effect otherwise in models T.3. Non-significant coeficients are shown with<br/>
       transparency, according to Wald-test p-value at level 0.05.") +
  theme(plot.caption = element_markdown(hjust = 0, lineheight = 1.5),
        plot.subtitle = element_markdown(size = 12, hjust = 0),
        plot.title = element_markdown(size = 16, hjust = 0)) -> secondaryPanel.pQTL

secondaryPanel.pQTL
# 9.8x9 landscape

auxT3 <- lapply(result_strat,"[[","PRSEffect")
auxT3 <- do.call(rbind,auxT3)
head(auxT3)

prsEffectList <- list("Models T1" = subset(res,DX.bl=="All",c(2,1,4:8,11,12)),
                      "Models T2" = resultQ2[c(9,1:8,10)],
                      "Models T3" = auxT3[c(2,3,1,4,5:12)]
                      )

prsEffectList <- lapply(prsEffectList,function(x){
  res <- x
  res$model <- with(res,paste(Volume,PRS,sep=":"))
  res <- res[c("model",names(x))]
  rownames(res) <- NULL
  return(res)
})


# Build Excel worksheet
wb <- createWorkbook()
invisible(lapply(names(prsEffectList),function(x)addWorksheet(wb,x)))
invisible(
  lapply(1:length(prsEffectList),
         function(i) writeDataTable(wb = wb,
                                    x = prsEffectList[[i]],
                                    sheet = names(prsEffectList)[i],
                                    withFilter = T,
                                    tableStyle = "TableStyleLight1")
  )
)

path <- here("results",paramT,"2_Modelling","1_baselineVolumes_groupAdjustedDifferences",
             "taules","crossSecModels_prsEffect.xlsx")

saveWorkbook(wb,file=path,overwrite = T)


# ModelsList
modelsT1 <- lapply(auxQ1[modGrid$DX.bl=="All"],"[[","ModelGlobal")
modelsT2 <- lapply(auxQ2,"[[","modDXInteract")
modelsT3 <- lapply(result_strat,"[[","modSxInteract")

# Parameters
modelParameters <- lapply(list(modelsT1,modelsT2,modelsT3),
                          FUN = enambleTable_xsecModels)
names(modelParameters) <- paste0("Models T",1:3)

# Build Excel worksheet
wb <- createWorkbook()
invisible(lapply(names(modelParameters),function(x)addWorksheet(wb,x)))
invisible(
  lapply(1:length(modelParameters),
         function(i) writeDataTable(wb = wb,
                                    x = modelParameters[[i]],
                                    sheet = names(modelParameters)[i],
                                    withFilter = T,
                                    tableStyle = "TableStyleLight1")
  )
)

path <- here("results",paramT,"2_Modelling","1_baselineVolumes_groupAdjustedDifferences",
             "taules","crossSecModels_parameters.xlsx")

saveWorkbook(wb,file=path,overwrite = T)
```
