## Supplementary material for "Genetic drivers of hippocampal atrophy highlight the role of *APOE* functional variants and AD polygenicity in Mild Cognitive Impairment": modelling_longitudinalAnalysis_all.R

```
###############################################################################
### MODELLING                                                                 #
###   Longitudinal Brain trajectories Analysis                                #
###                                                                           #
###############################################################################
rm(list=ls())
source("Rscripts/libraries.R")
source(here(RscriptsDir,"3_modelling","modelling_auxFunctions.R"))
       
### SELECT DATASET: 1.5T, 3T OR ALL----------------------------

  #·························································
  load(here(cleandataDir,"dataForAnalysis_mixed.RData"))
  paramT <- "All"
  
  # Omit the only measure at m60 in AD individuals
  aux <- dd$DX.bl == "AD" & dd$VISCODE2=="m60"
  dd <- dd[!aux,]
  dd$PTID <- droplevels(dd$PTID)
  
  # Error in brain measurements
  #dd <- dd[!dd$PTID%in%c("4337","0558"),]
  
  #·························································


### WORKING VARIABLES + PREVIOUS TRANSFORMATIONS -------------------------------

  # variables: hippocampal volumes, PRS continuous, PRS categorical
  
  # volVars <- sort(names(dd)[33:45])
  # volVars <- volVars[volVars %in% c("Whole_hippocampus","subiculum","presubiculum","parasubiculum")]
  volVars <- c("Whole_hippocampus","subiculum","presubiculum","parasubiculum")
  PRSVars <- names(dd)[7:12][1:2]
  riskVars <- names(dd)[46:51][1:2]
  snpVars <- names(dd)[13:23]
  dd$tesla <- as.factor(dd$tesla)
  #save(volVars,riskVars,snpVars,file=here(cleandataDir,"vars.RData"))

  # Data: select variables of interest and tranform time from months to years
  dd2 <- dd[c("PTID","AGE","DX.bl","PTGENDER","PTEDUCAT","ICV",
             "VISCODE2","timelapse","time_age","time_visit","tesla",
             riskVars, snpVars, volVars)]
  dd2$time_visit <- dd2$time_visit/12
  dd2$timelapse <- dd2$timelapse/12
  
  # snp pqtlSF.rs7412_T: dominant model (0 alleles, 1-2 alleles)
  dd2$pQTLCSF.rs7412_T <- ifelse(dd2$pQTLCSF.rs7412_T==0,"0","1-2")
  dd2$pQTLCSF.rs7412_T <- factor(dd2$pQTLCSF.rs7412_T,levels=c("0","1-2"))

#########
# Select longitudinal measures up to 5 years. We will use variable time_visit
# as a measure of longitudinal time (as time after baseline visit in years)
# It will take values as 0, 0.5, 1, 2, 3, 4 and 5.
# It is comparable to use age of individual at a given visit and center their
# values to the baseline age (AGE, baseline age).
# Mesures: bl,m3,m6,m12,m24,m36,m48,m60
  
dd2 <- subset(dd2,VISCODE2 %in% c("bl","m03","m06","m12","m24","m36","m48","m60"))
rownames(dd2) <- NULL


#===============================================================================
# 1) Number of observations for each group
#===============================================================================
pp <- dd2
pp <- pp[!pp$PTID%in%c("4337","0558"),] 

aux <- summaryBy(PTID ~ DX.bl+VISCODE2,
                 FUN=length,data=pp)
names(aux)[3] <- "numMeasures"

numMeasures_wide <- reshape(aux, direction = "wide",
                            v.names = "numMeasures",
                            idvar = "VISCODE2",
                            timevar = "DX.bl")

names(numMeasures_wide) <- c("VISCODE2","CN","MCI","AD")
numMeasures_wide[is.na(numMeasures_wide)] <- 0

#numMeasures_wide

### ADD REAL TIME FROM BASELINE 'time_visit' : mean(sd) [min,max]
f <- function(x){
  stats <- c(m=mean(x),sd=sd(x),min(x),max(x))
  sprintf("%.2f (%.2f) [%.2f,%.2f]",stats[1],stats[2],stats[3],stats[4])
}

timelapseTab <- summaryBy(timelapse ~ VISCODE2,data=pp,FUN=f)

taulaLongi <- merge(numMeasures_wide,timelapseTab,by="VISCODE2")
names(taulaLongi) <- c("Time Point","CN","MCI","AD","Time from baseline")

taulaLongi$`Time Point` <- c("Baseline", "Month 3","Month 6",
                             paste("Year",1:5))


taulaLongi[1,"Time from baseline"] <- 0


### iii) NUMBER OF INDIVIDUALS AT BASELINE
x <- basal$DX.bl
n <- sprintf("N = %.i",table(x))
n_header <- paste(paste(levels(dd$DX.bl),n,sep=", "),collapse="; ")


### IV) HTML TABLE
tab <- kbl(taulaLongi,align = "lcccr",
           caption = paste0("Number and timing of scans per time point by diagnostic group (",n_header,")."))


tab <- kable_classic(tab,full_width=F,font="Cambria")
formattedTab <- footnote(tab,
                         general = c("Time from baseline (in years) is in mean (standard deviation); ranges are listed in square brackets.",
                                     "Diagnostic groups: Controls (CN), Mild Cognitive Impairment (MCI) and Alzheimer's Disease (AD) "),
                         title_format = c("bold"))

formattedTab_longiMeasures <- row_spec(formattedTab,0,bold=T)

file <- "longitudinalMeasures_descriptiveTable.html"
dir.create(here("results",paramT,"2_Modelling"),F)
dir.create(here("results",paramT,"2_Modelling",
                "2_longitudinalTrajectories"),F)
dir.create(here("results",paramT,"2_Modelling",
                "2_longitudinalTrajectories","taules"),F)

path <- here("results",paramT,"2_Modelling",
             "2_longitudinalTrajectories","taules")

if(!(any(grepl(file,list.files(path))))){
  save_kable(formattedTab_longiMeasures,file=here(path,file),zoom=2)
}

### NUMBER OF MEASUREMENTS BY INDIVIDUAL
aux <- sapply(split(dd2,dd2$PTID),nrow)
measures <- data.frame(PTID=names(aux),numObs = aux)
measures <- merge(measures,basal[c("PTID","DX.bl")],by="PTID")
obsTab <- summaryBy(numObs ~ DX.bl, data=measures, FUN=summary)
names(obsTab) <- c("Diagnostic group","Min","Q1","Q2","Mean","Q3","Max")
obsTab


#===============================================================================
# 2) TRAJECTORIES PLOTS 
#===============================================================================

# Mean trend
pos <- position_dodge(width=0.2)

plotData <- dd2[,c("PTID","DX.bl","time_visit",volVars)]
plotData <- plotData[!plotData$PTID%in%c("4337","0558"),]
plotData <- reshape(plotData,
                    varying = names(plotData[-c(1:3)]),
                    timevar = "subfield",
                    times = names(plotData[-c(1:3)]),
                    idvar = c("PTID","time_visit"),
                    v.names = "volume",
                    direction = "long")
rownames(plotData) <- NULL
plotData$subfield <- factor(plotData$subfield,levels = volVars)
plotData <- plotData[plotData$time_visit!=0.25,]

path <- here("results",paramT,"1_DescriptiveAnalysis",
             "2_longitudinalMeasurements/smoothedTrajectories.png")
png(path,width = 800,height = 480)
ggplot(plotData,aes(x=time_visit,y=volume,group=DX.bl)) +
  geom_smooth(aes(linetype=DX.bl),method = "loess",se=F,color="black") +
  stat_summary(fun.data= "mean_cl_boot",size=.4)+
  scale_y_continuous(expand=c(0,0)) +
  scale_linetype_discrete("Diagostic group")+
  theme_classic()+
  facet_wrap(~as.factor(subfield),scales = "free_y") +
  ggtitle("Smoothed Mean measurement trajectories for different hippocampal subfield volumes")+
  labs(y = "Hippocampal volume (mm3)",
       caption="Hippocampal subfield mean volume at each measurement time after baseline (years), and 95% confidence interval.
                Smoothed trajectory was plotted using loess curve for each<br/>diagnositc group.
                It should be taken into account that subjects at baseline have different age.<br/>
       **Source**: ADNI project | **Units** Average hippocampal volume (mm3) of subjects at each measurement time after baseline (in years)") +
  theme(plot.caption = element_markdown(hjust = 0, lineheight = 1.5),
        plot.subtitle = element_markdown(size = 12, hjust = 0),
        plot.title = element_text(size = 16, hjust = 0),
        axis.title.x = element_blank())

dev.off()


vollabs <- c("Whole hippocampus","Subiculum","Presubiculum","Parasubiculum")
names(vollabs) <- volVars

path <- here("results",paramT,"1_DescriptiveAnalysis",
             "2_longitudinalMeasurements/smoothedTrajectories.pdf")
pdf(path,width = 8.56,height = 5.28)
ggplot(plotData,aes(x=time_visit,y=volume,group=DX.bl)) +
  geom_smooth(aes(linetype=DX.bl),method = "loess",se=F,color="black") +
  stat_summary(fun.data= "mean_cl_boot",size=.4)+
  scale_y_continuous(expand=c(0,0)) +
  scale_linetype_discrete("Diagostic group")+
  theme_classic()+
  facet_wrap(~subfield,scales = "free_y",
             labeller=labeller(subfield=vollabs)) +
  labs(y = bquote('Hippocampal subfields volume  ' ~(mm^3)),
       x = "\nTime after baseline (years)")+
  theme(plot.caption = element_markdown(hjust = 0, lineheight = 1.5),
        plot.subtitle = element_markdown(size = 12, hjust = 0),
        plot.title = element_text(size = 16, hjust = 0),
        axis.title = element_text(size=16),
        axis.text = element_text(size=12),
        strip.text = element_text(size=12))

dev.off()


#===============================================================================
# 3) Hippocampal volume trajectory analyses
# --------------------------------------------------------------------------
#    OBJECTIVES
#
#     0) Check the diagnostic of residuals and detect outliers.
#
#     1) Check if there is a significant reduction in hippocampal volumes 
#       over time (Model 1).
#
#     2) See if this trajectory is different depending on the diagnostic 
#       group (Model 2).
#
#     3) Using Model 2 with fixed effects and random slope and intercept 
#       as random, test if the random slope improves the model.
#
#     4) Verify if in any of the volumes the quadratic term for time 
#      is significant, to consider non-linearity in the trajectory (Model 3).
#
#     5) Check if there is a specific effect of sex on the rate of reduction 
#      of hippocampal volumes that differs by diagnostic group.
#
#
#------------------------------------------------------------------------------
#   PROCEDURE:
#
#     Obj.0: Review the diagnostic plots and analyze subjects with observations 
#      that have high residuals.
#
#     Obj.1: Fit a mixed linear model for each volume with time_visit as the 
#            temporal variable, correcting for diagnostic group (DX.bl), 
#            baseline age (AGE), sex (PTGENDER), years of education (PTEDUCAT), 
#            and total intracranial volume in cm³ (ICV/1000).
#            To account for the correlation between measurements from the same 
#            subject, allow each individual to have their own starting point and 
#           temporal slope (random intercept and slope by PTID).
#            (Additive model, 1):
#
#         Vol_i ~ time_visit + DX.bl + Covariates + (1+time_visit|PTID)
#
#     Obj.2: Consider in the previous model that the effect of time_visit on 
#            the response volume depends on the diagnostic group, by adding an 
#            interaction term time_visit * DX.bl. (Diagnostic model, 2).
#
#         Vol_i ~ time_visit * DX.bl + Covariates + (1+time_visit|PTID)
#
#     Obj.3: Apply a likelihood ratio test to compare the nested models with 
#         and without random slope, based on a chi-square mixture statistic 
#         (using the ranova() function from lmerTest). Correct for multiple
#         comparisons using FDR at 5%.
#
#     Obj.4: Add the quadratic term of time_visit both as a main effect and 
#         with interaction with DX.bl. Perform the simultaneous contrast that 
#         all coefficients involving the quadratic term of time are 0. Retrieve 
#         the p-value of the contrast. If it is less than 0.05, it is considered 
#         that one of the coefficients of the quadratic term has an effect. 
#         (Model 3).
#
#         Vol_i ~ (time_visit + time_visit^2) * DX.bl + Covariates + 
#                  (1+time_visit|PTID)
#
#     Obj.5: Add an interaction term time_visit*PTGENDER to model 2 and 
#        test the significance of the coefficient.
#
#         Vol_i ~ time_visit * DX.bl + time_visit * PTGENDER + 
#           Covariates + (1+time_visit|PTID)
#
#
#    NOTE: Covariates are PTGENDER, PTEDUCAT, AGE, ICV/1000.
#    Random effects: random intercept and time_visit slope by PTID.
#    The quadratic effect of time_visit is not considered, as it was not found 
#    to be significant in section 1.
#===============================================================================

#--------------------------------------------------------------------------------
# FUNCTION timeEffect(): For each volume, perform the procedures for the four
#  previous objectives, and return a list of objects as the result to answer them.
#--------------------------------------------------------------------------------

volume <- "Whole_hippocampus"; data<-dd2

timeEffect <- function(volume,data){
  # OBJ1: 
  predictors <- "time_visit + DX.bl + AGE + I(ICV/1000) + PTGENDER + PTEDUCAT + tesla + (1+time_visit|PTID)"
  form <- paste(volume,predictors, sep = " ~ ")
  mod1 <- lme4::lmer(formula = form, data=data,
                     control = lmerControl(optimizer ="Nelder_Mead"))
  
  # OBJ2: 
  predictors <- "time_visit * DX.bl + AGE + I(ICV/1000) + PTGENDER + PTEDUCAT + tesla + (1+time_visit|PTID)"
  form <- paste(volume,predictors, sep = " ~ ")
  mod2 <- lmerTest::lmer(formula = form, data=data,
                     control = lmerControl(optimizer ="Nelder_Mead"))
  
  # OBJ3: 
  ranovaTest <- ranova(mod2)$`Pr(>Chisq)`[2]
  
  # OBJ4: 
  mod3 <- update(mod2,~.+I(time_visit^2)+I(time_visit^2):DX.bl)
  posTerms <- grep("time_visit\\^2",names(fixef(mod3)))
  terms <- names(fixef(mod3))[posTerms] # coeficients time_visit^2 
  
  # Test existence of quadratic time effect
  timeQuad <- car::linearHypothesis(mod3,terms,c(0,0,0))$`Pr(>Chisq)`[2]
  
  # OBJ5:
  predictors <- "time_visit * DX.bl + time_visit * PTGENDER + AGE + I(ICV/1000) + PTEDUCAT + tesla + (1+time_visit|PTID)"
  form <- paste(volume,predictors, sep = " ~ ")
  mod4 <- lme4::lmer(formula = form, data=data,
                     control = lmerControl(optimizer ="Nelder_Mead"))
  
  # OBJ0: Residuals
  #resid <- plot(mod2,main = paste("Diagnositc",volume))
  resid <- resid_panel(mod2)
  
  return(list(modelAdditiu = mod1,
              modelDiagEffect = mod2,
              modelSexEffect = mod4,
              ranovaTest = ranovaTest,
              efecteQuadratic = timeQuad,
              diagnostic = resid))
}
#--------------------------------------------------------------------------------

obj_is_vector = function(x){TRUE}
timeEffectModels <- lapply(volVars,FUN=timeEffect, data=dd2)
names(timeEffectModels) <- volVars


#-------------------------------------------------------------------------------
# A) OBJ0: model diagnosis
#  
#-------------------------------------------------------------------------------

lapply(timeEffectModels,"[[","diagnostic")

outliers <- lapply(timeEffectModels,function(x){
  car::outlierTest(x$modelDiagEffect)
})

aux <- lapply(outliers,function(x)as.integer(names(x$rstudent))) 
aux <- lapply(aux,function(x)as.character(unique(dd2[x,"PTID"]))) 
(tab <- table(do.call(c,aux))) 
(ptid_outlier <- names(sort(tab,decreasing = T))[1:2]) 

# Incoherent measures
dd2[dd2$PTID %in% ptid_outlier,c("PTID","DX.bl","time_visit",volVars)]

# Exclude individual and re-run again models.
dd2 <- subset(dd2, !(PTID %in% ptid_outlier))
dd2$PTID <- droplevels(dd2$PTID)
rownames(dd2) <- NULL
# length(unique(dd2$PTID))
# nrow(dd2)

timeEffectModels <- lapply(volVars,FUN=timeEffect, data=dd2)
names(timeEffectModels) <- volVars
lapply(timeEffectModels,"[[","diagnostic")

## Save diagnostics
dir.create(here("results",paramT,"2_Modelling",
                "2_longitudinalTrajectories","modelDiagnostics"),F)
lapply(names(timeEffectModels),FUN=function(x){
  path <- here("results",paramT,"2_Modelling","2_longitudinalTrajectories",
               "modelDiagnostics")
  file <- paste0(path,"/modelResiduals_",x,".png")  
  ggsave(filename = file, plot=timeEffectModels[[x]]$diagnostic,device = "png")
})

# saveRDS(dd2,here("data","cleandata","datasetForAnalysis.long.rds"))

#-------------------------------------------------------------------------------
# A) OBJ1
#    Is there a change over time in hippocampal volumes?
#--------------------------------------------------------------------------------
rateChangeVolumes <- 
  lapply(timeEffectModels,function(x){
    fixef <- extract_fixed_effects(x$modelAdditiu)[2,]
    effect <- sprintf("%.2f (%.2f,%.2f); %.3f", 
                      fixef$value,fixef$lower_2.5,fixef$upper_97.5,fixef$p_value)
    
    return(effect)
  })

rateChangeVolumes <- do.call(c,rateChangeVolumes)
rateChangeVolumes <- data.frame(timeEffect = rateChangeVolumes)

rateChangeVolumes


#-------------------------------------------------------------------------------
# B) OBJ2
#    Is there any difference in the change in volume according to the 
#    diagnostic group?
#-------------------------------------------------------------------------------
effectDXbl <- 
  sapply(timeEffectModels, function(x){
    pval_contrast <- car::linearHypothesis(x$modelDiagEffect,
                                           c("time_visit:DX.blMCI","time_visit:DX.blAD"),
                                           c(0,0))[2,"Pr(>Chisq)"]
    return(round(pval_contrast,3))
  })

effectDXbl 
round(p.adjust(effectDXbl,method = "fdr"),3)

volume <- timeEffectModels$CA1
timeTrendDx <- lapply(timeEffectModels,function(volume){
  mod <- volume$modelDiagEffect

  # timeTrend <- emtrends(mod,specs = "DX.bl",var="time_visit",
  #                       at=list(PTGENDER="Female"),pbkrtest.limit=4176)
  timeTrend <- emtrends(mod,specs = "DX.bl",var="time_visit",
                        at=list(PTGENDER="Female"),pbkrtest.limit=4176)
  contrast <- pairs(timeTrend)
  
  # Time trend
  aux <- t(summary(timeTrend)[,-1])
  slopes <- apply(aux,2,function(x){sprintf("%.2f (%.2f,%.2f)",x[1],x[4],x[5])})
  names(slopes) <- summary(timeTrend)[,1]
  
  #Contrast
  pvals <- summary(contrast)$p.value
  pvals <- ifelse(pvals<0.001,"<.001",sprintf("%.3f",pvals))
  names(pvals) <- summary(contrast)$contrast

  # Result
  res <-as.data.frame(t(matrix(c(slopes,pvals))))
  names(res) <- Reduce("c",names(c(slopes,pvals)))
  return(res)
})

timeTrendDx <- do.call(rbind,timeTrendDx)
timeTrendDx

tab <- kbl(timeTrendDx,align = "cccccc",
           caption = "Estimated mean annual volume changes by group diagnostic")


tab <- kable_classic(tab,full_width=F,font="Cambria")
tab <- add_header_above(tab,c(" " = 1,
                              "Estimated linear time trend"=3,
                              "Pairwise comparisons"=3), 
                        bold = T,line = T)
formattedTab <- footnote(tab,
                         general = c("Linear time trend (95% CI) as linear combination of time-related coefficients. It indicates mean annual volume change in mm3 estimated for a given diagnostic
                                     group and hippocampal region. Differences in time trend between diagnostic groups were assessed by Wald tests for linear contrasts. Pvalues are reported",
                                     "Diagnostic groups: Controls (CN), Mild Cognitive Impairment (MCI) and Alzheimer's Disease (AD) "),
                         title_format = c("bold"))

formattedTab_timeTrendDX <- row_spec(formattedTab,0,bold=T)
formattedTab_timeTrendDX

file <- "modelL0_timeTrends_byDXbl.html"
dir.create(here("results",paramT,"2_Modelling"),F)
dir.create(here("results",paramT,"2_Modelling",
                "2_longitudinalTrajectories"),F)
dir.create(here("results",paramT,"2_Modelling",
                "2_longitudinalTrajectories","taules"),F)

path <- here("results",paramT,"2_Modelling",
             "2_longitudinalTrajectories","taules")

if(!(any(grepl(file,list.files(path))))){
  save_kable(formattedTab_timeTrendDX,file=here(path,file),zoom=2)
}


#-------------------------------------------------------------------------------
# C) OBJ3
#    Does the random slope over time improve the models?
#-------------------------------------------------------------------------------

ranovaTest <- do.call(c,lapply(timeEffectModels,"[[","ranovaTest"))
p.adjust(ranovaTest,method = "fdr")

#-------------------------------------------------------------------------------
# D) OBJ4
#    Is there a quadratic effect of time?
#-------------------------------------------------------------------------------

quadraticTimeEffect <- do.call(c,lapply(timeEffectModels,"[[","efecteQuadratic"))
round(p.adjust(quadraticTimeEffect,method = "fdr"),3)

#-------------------------------------------------------------------------------
# E) OBJ5
#    Sex effects? beta_timeVisit:PTGENDER
#-------------------------------------------------------------------------------
# Per debuggin: x <- timeEffectModels[[1]]

timeTrendSx <- lapply(timeEffectModels,function(volum){
  mod <- volum$modelSexEffect
  
  timeTrend <- emtrends(mod,specs = "PTGENDER",var="time_visit",
                        pbkrtest.limit=4176)
  contrast <- pairs(timeTrend)
  
  # Time trend
  aux <- t(summary(timeTrend)[,-1])
  slopes <- apply(aux,2,function(x){sprintf("%.2f (%.2f,%.2f)",x[1],x[4],x[5])})
  names(slopes) <- summary(timeTrend)[,1]
  
  #Contrast
  pvals <- summary(contrast)$p.value
  pvals <- ifelse(pvals<0.001,"<.001",sprintf("%.3f",pvals))
  names(pvals) <- summary(contrast)$contrast
  
  # Result
  res <-as.data.frame(t(matrix(c(slopes,pvals))))
  names(res) <- Reduce("c",names(c(slopes,pvals)))
  return(res)
})

timeTrendSx <- do.call(rbind,timeTrendSx)
timeTrendSx


#===============================================================================
# 4) Analysis of the effect of genetic predisposition on the trajectory of 
#  hippocampal volume reduction.
#    --------------------------------------------------------------------------
#    OBJECTIVES:
#     1) check if genetic predisposition to neurodegenerative traits and aging 
#       influences the rate of hippocampal volume reduction.
#
#     2) If this genetic effect time_visit*PRS exists, is it different depending 
#       on the individual's state on the disease continuum, DX.bl?
#
#     3) Is this genetic effect different according to sex?
#
#------------------------------------------------------------------------------
#   PROCEDURE:
#     
#--------------------------------------------------------------------------------
# FUNCTION: prsTimeEffect(): for each volum and PRS performs obj1 procedure +
# adds a triple interaction.
#--------------------------------------------------------------------------------

#-------------------------------------------------------------------------------
prsTimeEffect <- function(i,data,modGrid){
  volume <- modGrid[i,"Volume"]
  PRS <- modGrid[i,"PRS"]
  
  # Formula ··································
  predictors <- paste("time_visit",c("DX.bl",PRS),sep=" * ")
  predictors_3intDX <- paste("time_visit","DX.bl",PRS,sep=" * ") #triple interact DX
  predictors_3intSx <- paste("time_visit","PTGENDER",PRS,sep=" * ") #triple interact Sex

#  covars <- c("AGE","I(ICV/1000)","DX.bl","PTGENDER","PTEDUCAT")
  covars <- c("AGE","I(ICV/1000)","DX.bl","PTGENDER","PTEDUCAT","tesla")
  random <- "(1 + time_visit|PTID)"
  
  form <- 
    sapply(list(predictors,predictors_3intDX,predictors_3intSx),function(x){
      res <- paste(c(x,covars,random),collapse= " + ")
      res <- paste(volume,res, sep = " ~ ")
      return(res)
    })
  
  ## OBJ1
  mod <- lme4::lmer(formula = form[1], data=data,
                    control = lmerControl(optimizer ="Nelder_Mead"))
  
  attributes(mod)$PRS <- PRS
  attributes(mod)$volume <- volume
  
  
  fixef <- extract_fixed_effects(mod)
  effect_pos <- grep(paste0("^time_visit:",PRS),fixef$term)

  res <- as.data.frame(fixef[effect_pos,c("value","se","lower_2.5",
                                          "upper_97.5","p_value")])
  
  rownames(res) <- paste(volume,PRS,sep=":")
  #······································································
  
  ## OBJ2
  mod_3IntDX <- lme4::lmer(formula = form[2], data=data,
                     control = lmerControl(optimizer ="Nelder_Mead"))
  
  #testDX <- drop1(mod_3IntDX, test="Chisq")[paste("time_visit","DX.bl",PRS,sep=":"),"Pr(Chi)"]
  testDX <- anova(mod_3IntDX,mod, test="LRT")[2,"Pr(>Chisq)"]
  attributes(mod_3IntDX)$PRS <- PRS
  attributes(mod_3IntDX)$volume <- volume
  
  #······································································
  
  ## OBJ3
  form[3] <- sub("DX.bl","time_visit * DX.bl",form[3])
  mod_3IntSx <- lme4::lmer(formula = form[3], data=data,
                     control = lmerControl(optimizer ="Nelder_Mead"))
  
  testSx <- anova(mod_3IntSx,mod, test="LRT")[2,"Pr(>Chisq)"]
  #testSx <- drop1(mod_3IntSx, test="Chisq")[paste("time_visit","PTGENDER",PRS,sep=":"),"Pr(Chi)"]
  attributes(mod_3IntSx)$PRS <- PRS
  attributes(mod_3IntSx)$volume <- volume
  
  return(list("modGlobal"= mod,
              "PRS_timeEffect" = res,
              "AnovaTest_3wayIntDX" = testDX,
              "mod_3wayIntDX" = mod_3IntDX,
              "AnovaTest_3wayIntSx" = testSx,
              "mod_3wayIntSx" = mod_3IntSx))
}
#--------------------------------------------------------------------------


#-------------------------------------------------------------------------------
# A) OBJ1
#    Effect of PRS on volume reduction?
#--------------------------------------------------------------------------------

### i) Matrix of PRS and hippocampal volume combinations.
modGrid <- expand.grid(PRS = c(riskVars,snpVars),
                       Volume = volVars,
                       stringsAsFactors = F)
# head(modGrid)
#prsTimeEffect(1,data=dd2,modGrid=modGrid)
#i <- 136
modGrid$analysis[modGrid$PRS %in% c(riskVars,"APOE4")] <- "Primary"
modGrid$analysis[modGrid$PRS %in% snpVars[-1]] <- "Secondary"
# modGrid$analysis[modGrid$PRS %in% snpVars[2:7]] <- "Univariate.brain"
# modGrid$analysis[modGrid$PRS %in% snpVars[8:11]] <- "Univariate.CSF"

### ii) Apply the function effectPRS_strat() on the GRID.
#library(parallel)
# result <- lapply(1:nrow(modGrid),
#                    FUN = prsTimeEffect,
#                    data=dd2,
#                    modGrid=modGrid)
# 
# names(result) <- paste(modGrid$Volume,modGrid$PRS,sep=":")
# saveRDS(result,file=here(cleandataDir,"results.rds"))
result <- readRDS(here(cleandataDir,"results.rds"))

# i arreglem el resultat...
res_prsTimeEffect <- lapply(result,"[[","PRS_timeEffect") # seleccionar efecte PRS de la list
res_prsTimeEffect <- do.call(rbind,res_prsTimeEffect) # consolidar en dataFrame
res_prsTimeEffect <- cbind(modGrid,res_prsTimeEffect) # Afegir PRS i volume 
rownames(res_prsTimeEffect) <- NULL # no rownames
res_prsTimeEffect$PRS <- factor(res_prsTimeEffect$PRS,levels=c(riskVars,snpVars)) 

head(res_prsTimeEffect,17)

### iii) FDR correction

res_prsTimeEffect$prsVol <- paste(res_prsTimeEffect$PRS,res_prsTimeEffect$Volume,sep="|")
aux <- lapply(split(res_prsTimeEffect,res_prsTimeEffect$analysis), function(x){
  x$pvalue_corrected <- p.adjust(x$p_value, method = "fdr")
  return(x[c("prsVol","pvalue_corrected")])
})
aux <- do.call(rbind,aux)
pos <- match(res_prsTimeEffect$prsVol,aux$prsVol)
res_prsTimeEffect$pvalue_corrected <- aux[pos,"pvalue_corrected"]
res_prsTimeEffect$prsVol <- NULL


# res_prsTimeEffect <- res_prsTimeEffect[res_prsTimeEffect$PRS%in%riskVars[1:6],]
# aux <- lapply(split(res_prsTimeEffect,res_prsTimeEffect$Volume),
#               FUN=function(x)p.adjust(x$p_value,method="fdr"))
# res_prsTimeEffect$pvalue_corrected <- do.call(c,aux)

### iv) Significance
res_prsTimeEffect[paste(names(res_prsTimeEffect)[8:9],"symbol",sep="_")] <-
  lapply(res_prsTimeEffect[names(res_prsTimeEffect[8:9])], FUN=cut,
         breaks = c(-Inf,0.001,0.01,0.05,0.1,1),
         labels=c("***","**","*","·",""))

head(res_prsTimeEffect)
(significant_effects <- subset(res_prsTimeEffect, pvalue_corrected_symbol!=""))

tab <- significant_effects[c(2,1,4:7,9)]
names(tab) <- c("Volume","PRS","Beta coef","SE","lower_CI","upper_CI","pvalue")
tab$pvalue <- ifelse(tab$pvalue<0.001,"<0.001",round(tab$pvalue,3))

tab <- kbl(tab,align = "llcccccc",row.names = F,
           caption = "Significant effects of PRS on time trend. Models L.1")


tab <- kable_classic(tab,full_width=F,font="Cambria")
formattedTab <- footnote(tab,
                         general = c("Significant linear coefficients for interaction between PRS and time. Pvalues were corrected for multiple comparisons using FDR method."),
                         title_format = c("bold"))

formattedTab_prsTimeEffect <- row_spec(formattedTab,0,bold=T)
formattedTab_prsTimeEffect
# save_kable(x = formattedTab_prsTimeEffect,
#            file = here("results",paramT,"2_Modelling",
#                        "2_longitudinalTrajectories","taules",
#                        "modelL1_betasPRSTime.png"),zoom=1.5)


tab <- res_prsTimeEffect
rownames(tab) <- NULL
tab <- split(tab,tab$Volume)

# Build Excel worksheet
wb <- createWorkbook()
invisible(lapply(names(tab),function(x)addWorksheet(wb,x)))
invisible(
  lapply(1:length(tab),
         function(i) writeDataTable(wb = wb,
                                    x = tab[[i]],
                                    sheet = names(tab)[i],
                                    withFilter = T,
                                    tableStyle = "TableStyleLight1")
  )
)

path <- here("results",paramT,"2_Modelling","2_longitudinalTrajectories",
             "taules","modelL1_interactionTimeVsGenetics.xlsx")
saveWorkbook(wb,file=path,overwrite = T)


### v) HEATMAP
plotData <- res_prsTimeEffect
plotData <- plotData[plotData$analysis=="Primary",]
upper.th <- quantile(plotData$value,1)
lower.th <- quantile(plotData$value,0.05)

plotData$Volume <- factor(plotData$Volume,levels=unique(plotData$Volume),
                          labels=c("Whole hippocampus","Subiculum","Presubiculum","Parasubiculum"))

heatMap <- 
  ggplot(data=plotData,
         aes(x=Volume, y=PRS, fill=pmax(pmin(value,upper.th),lower.th))) + 
   geom_tile(show.legend = T,color="grey") +
  geom_text(aes(label=pvalue_corrected_symbol),size=6)+
  scale_fill_gradient2(low="red4",mid = "white", high="#0066FF", midpoint = 0,
                       name="beta")+
  # facet_wrap(~DX.bl)+
  ggstatsplot::theme_ggstatsplot() +
  theme(strip.background = element_blank(),
        plot.margin = margin(1,1,.5,1,"cm"),
        axis.title = element_blank(),
        axis.text = element_text(face="bold",size = 8)) +
  
#  ggtitle("Corrected significant effect of PRS and individual SNPs<br>on hippocampal subfield volume's rate of change over time") +
  labs(caption="<br>Linear coefficient for interaction between time and AD PRS or APOE4 status. Effects adjusted for Age at baseline (years), intracranial<br/>
  volume(cm3), sex, diagnostic status, years of education and tesla parameter. Model allows a diferent volume rate of change depending  on<br/>
  diagnositc  status. Also, intercept and rate of change in time are included as random effects. Significant interactions are reported as Wald-test<br/>
  pvalues at levels 0.1(·), 0.05(\\*), 0.01(\\**) and 0.001(\\***) and corrected for multiple comparisons by FDR method. Non-significant differences<br/>
  are not shown. Colors intensity is saturated to 5 and 95% percentile values<br/>
  **Source**: ADNI project | **Units** Average increment in volume rate of change (mm3) associated with having high predisposition for the PRS trait.") +
  theme(plot.caption = element_markdown(hjust = 0, lineheight = 1.5, size=6),
        plot.subtitle = element_markdown(size = 12, hjust = 0),
        plot.title = element_text(size = 16, hjust = 0))

heatMap.primary <- heatMap
heatMap.primary


plotData <- res_prsTimeEffect
plotData <- plotData[plotData$analysis=="Secondary",]
upper.th <- quantile(plotData$value,.95)
lower.th <- quantile(plotData$value,0.07)

plotData$Volume <- factor(plotData$Volume,levels=unique(plotData$Volume),
                          labels=c("Whole hippocampus","Subiculum","Presubiculum","Parasubiculum"))

heatMap <- 
  ggplot(data=plotData,
         aes(x=Volume, y=PRS, fill=pmax(pmin(value,upper.th),lower.th))) + 
  geom_tile(show.legend = T,color="grey") +
  geom_text(aes(label=pvalue_corrected_symbol),size=6)+
  scale_fill_gradient2(low="red4",mid = "white", high="#0066FF", midpoint = 0,
                       name="beta")+
  # facet_wrap(~DX.bl)+
  ggstatsplot::theme_ggstatsplot() +
  theme(strip.background = element_blank(),
        plot.margin = margin(1,1,.5,1,"cm"),
        axis.title = element_blank(),
        axis.text = element_text(face="bold",size = 8)) +
  
  #  ggtitle("Corrected significant effect of PRS and individual SNPs<br>on hippocampal subfield volume's rate of change over time") +
  labs(caption="<br>Linear coefficient for interaction between time and AD PRS or APOE4 status. Effects adjusted for Age at baseline (years), intracranial<br/>
  volume(cm3), sex, diagnostic status, years of education and tesla parameter. Model allows a diferent volume rate of change depending  on<br/>
  diagnositc  status. Also, intercept and rate of change in time are included as random effects. Significant interactions are reported as Wald-test<br/>
  pvalues at levels 0.1(·), 0.05(\\*), 0.01(\\**) and 0.001(\\***) and corrected for multiple comparisons by FDR method. Non-significant differences<br/>
  are not shown. Colors intensity is saturated to 5 and 95% percentile values<br/>
  **Source**: ADNI project | **Units** Average increment in volume rate of change (mm3) associated with having high predisposition for the PRS trait.") +
  theme(plot.caption = element_markdown(hjust = 0, lineheight = 1.5, size=6),
        plot.subtitle = element_markdown(size = 12, hjust = 0),
        plot.title = element_text(size = 16, hjust = 0))

heatMap.secondary <- heatMap
heatMap.secondary

#7.86x3.63


#-------------------------------------------------------------------------------
# B) OBJ2
#    Does the effect of PRS on volume reduction depend on the diagnostic status?
#-------------------------------------------------------------------------------

# Interaction time_visit*PRS*DX.bl significant?
testDX <- sapply(result,"[[","AnovaTest_3wayIntDX")
cbind(testDX[testDX<0.05])


# Debbuging: x<-result$`CA1:AD_predisposition` #
trends <- lapply(result,function(x){
  mod <- x$mod_3wayIntDX
  PRS <- attributes(mod)$PRS
  specs <- as.formula(paste0("~",PRS,"*DX.bl"))
  at <- "list(PTGENDER='Female')"
  if(PRS %in% snpVars){
    levs <- paste0(names(table(dd2[,PRS])),collapse=",")
    at <- paste0("list(PTGENDER = 'Female', ",
                 PRS," = c(",levs,"))")
  }
  if(PRS == "pQTLCSF.rs7412_T"){
    at <- paste0("list(PTGENDER = 'Female', ",
                 PRS," = c('0','1-2'))")
  }
  
      
  trend <- emtrends(mod, specs = specs, "time_visit",
                    at = eval(parse(text=at)),
                    pbkrtest.limit=4186)
  
  return(trend)
})

trends.df <- lapply(trends,summary)

 trends.df <- lapply(trends.df,function(x){
  x$variable <- names(x)[1]
  names(x)[1] <- "PRS_value"
  x$PRS_value <- as.character(x$PRS_value)
  x <- x[c("variable","PRS_value",names(x)[-c(1,length(names(x)))])]
  return(x)
})

trends.df <- do.call(rbind,trends.df)
vol <- sapply(strsplit(rownames(trends.df),":"),"[[",1)
trends.df <- cbind(Volumes = vol,trends.df)
rownames(trends.df) <- NULL

contrastPRS <- lapply(trends,function(x){
  n <- names(x@levels)[1]
  s <- summary(contrast(x,"consec",simple=n))
  if(nrow(s)>3)
  {
    s <- do.call(rbind,lapply(split(s,s$DX.bl),"[",1,)) 
  }
  # Get volumes and PRS variables
  Volume <- as.character(x@model.info$call[[2]])[2]
  PRS <- n
  s <- cbind(Volume,PRS,s)
  return(s)
})

res_contrastPRS <- as.data.frame(do.call(rbind,contrastPRS))
rownames(res_contrastPRS) <- NULL


 # FDR
res_contrastPRS$analysis <- modGrid[match(res_contrastPRS$PRS,modGrid$PRS),"analysis"]
res_contrastPRS$prsVol <- paste(res_contrastPRS$PRS,
                                res_contrastPRS$Volume,
                                res_contrastPRS$DX.bl,
                                  sep="|")

aux <- lapply(split(res_contrastPRS,res_contrastPRS$analysis), function(x){
  x$pvalue_corrected <- p.adjust(x$p.value, method = "fdr")
  return(x[c("prsVol","p.value","pvalue_corrected")])
})
aux <- do.call(rbind,aux)
pos <- match(res_contrastPRS$prsVol,aux$prsVol)
res_contrastPRS$pvalue_corrected <- aux[pos,"pvalue_corrected"]
#res_contrastPRS$prsVol <- NULL

res_contrastPRS$pvalue_corrected_symbol <-
  cut(res_contrastPRS$pvalue_corrected,
      breaks = c(-Inf,0.001,0.01,0.05,0.1,1),
      labels=c("***","**","*","·",""))

head(res_contrastPRS)

contrast1_signif <- subset(res_contrastPRS,pvalue_corrected<0.05)
prsDiff <- with(contrast1_signif,paste(Volume,PRS,sep=":"))

contrast1_signif[c("prsVol","analysis")] <- NULL
contrast1_signif[5:8] <- lapply(contrast1_signif[5:8],round,2)
contrast1_signif[c(9:10)] <- lapply(contrast1_signif[c(9:10)],function(x){
  ifelse(x<0.001,"<0.001",sprintf("%.3f",x))
})

contrast1_signif$pvalue_corrected <- 
  with(contrast1_signif,paste0(pvalue_corrected,"(",pvalue_corrected_symbol,")"))

rownames(contrast1_signif) <- NULL
contrast1_signif$pvalue_corrected_symbol <- NULL

tab <- kbl(contrast1_signif,align = "lllccccccc",row.names = F,
           caption = "Significant effects of PRS on time trend. Models L.2, linear contrasts 1")


tab <- kable_classic(tab,full_width=F,font="Cambria")
formattedTab <- footnote(tab,
                         general = c("Wald test for $H_0: \\beta5_{time:High} + \\beta7_{time:High:MCI}=0$.", 
                                     "Pvalues were corrected for multiple comparisons using FDR method. Significance level: (*)<.05, (**)<.01, (***)<.001"),
                         title_format = c("bold"))

formattedTab_contrast1 <- row_spec(formattedTab,0,bold=T)
formattedTab_contrast1

# save_kable(x = formattedTab_contrast1,
#            file = here("results",paramT,"2_Modelling",
#                        "2_longitudinalTrajectories","taules",
#                        "modelL2_contrast1Signif.png"),zoom=1.5,)

  tab <- res_contrastPRS
  rownames(tab) <- NULL
  tab <- split(tab,tab$Volume)

  # Build Excel worksheet
  wb <- createWorkbook()
  invisible(lapply(names(tab),function(x)addWorksheet(wb,x)))
  invisible(
    lapply(1:length(tab),
           function(i) writeDataTable(wb = wb,
                                      x = tab[[i]],
                                      sheet = names(tab)[i],
                                      withFilter = T,
                                      tableStyle = "TableStyleLight1")
    )
  )

path <- here("results",paramT,"2_Modelling","2_longitudinalTrajectories",
             "taules","modelL2_contrast1_DXbl.xlsx")
saveWorkbook(wb,file=path,overwrite = T)

aux <- lapply(prsDiff,function(x){
  
  linfct <- contrast(trends[[x]],interaction=c("consec","pairwise"))@linfct
  mod <- result[[x]]$mod_3wayIntDX
  joinTest <- summary(glht(mod,linfct),test=Chisqtest())
  
  res <- data.frame(Volumes = attributes(mod)$volume,
                    variable = attributes(mod)$PRS,
                    W2_chisq = as.numeric(joinTest$test$SSH),
                    df = as.numeric(joinTest$test$df[[1]]),
                    Pval = joinTest$test$pvalue)
  res$symbolJoinTest <- ifelse(res$Pval<0.1,"*","")
  return(res)
})
contrast2 <- do.call(rbind,aux)

tab <- contrast2
tab$W2_chisq <- round(tab$W2_chisq,2)
tab$Pval <- ifelse(tab$Pval<0.001,"<0.001",sprintf("%.3f",tab$Pval))
tab$Pval <- with(tab,ifelse(symbolJoinTest=="*",paste0(Pval,"(*)"),Pval))
tab$symbolJoinTest <- NULL

tab <- kbl(tab,align = "llccc",row.names = F,
           caption = "Differences between diagnostic groups regarding PRS effects on volume rate of change. Models L2, linear contrasts 2")


tab <- kable_classic(tab,full_width=F,font="Cambria")
formattedTab <- footnote(tab,
                         general = c("Simultaneous Wald Test for 3 linear contrast. $H_0:\\beta7_{time:MCI:High}=0 ;\\beta7_{time:AD:High}=0; \\beta7_{time:AD:High}-\\beta7_{time:MCI:High}=0$",
                                     "Significance level: (*)<.05"),
                         title_format = c("bold"))

formattedTab_contrast2 <- row_spec(formattedTab,0,bold=T)
formattedTab_contrast2

tab <- contrast2
tab <- split(tab,tab$Volume)

# Build Excel worksheet
wb <- createWorkbook()
invisible(lapply(names(tab),function(x)addWorksheet(wb,x)))
invisible(
  lapply(1:length(tab),
         function(i) writeDataTable(wb = wb,
                                    x = tab[[i]],
                                    sheet = names(tab)[i],
                                    withFilter = T,
                                    tableStyle = "TableStyleLight1")
  )
)

path <- here("results",paramT,"2_Modelling","2_longitudinalTrajectories",
             "taules","modelL2_contrast2.xlsx")
saveWorkbook(wb,file=path,overwrite = T)

finalResult <- trends.df
finalResult$ident <- with(finalResult,
                          paste(variable,Volumes,DX.bl,sep="|"))


pos <- match(finalResult$ident,res_contrastPRS$prsVol)
finalResult$pvalue <- res_contrastPRS[pos,"p.value"]
finalResult$pvalue_corrected <- res_contrastPRS[pos,"pvalue_corrected"]


finalResult$alpha <- with(finalResult,ifelse(pvalue_corrected<0.05,1,.9))
finalResult$alpha2 <- with(finalResult,ifelse(pvalue_corrected<0.05,1,.4))

finalResult$ident <- with(finalResult,
                          paste(variable,Volumes,sep="|"))
pos <- match(finalResult$ident,paste(contrast2$variable,contrast2$Volumes,sep="|"))
finalResult$symbolJoinTest <- contrast2[pos,"symbolJoinTest"]

finalResult$symbolJoinTest[is.na(finalResult$symbolJoinTest)] <- ""

finalResult <- within(finalResult,{
  symbolJoinTest[symbolJoinTest=="*" & (PRS_value %in% c("Low-Intermediate","0","2") | 
                                          DX.bl!="MCI")] <- ""
})

finalResult$analysis <- modGrid[match(finalResult$variable,modGrid$PRS),"analysis"]

names(finalResult)[names(finalResult)=="DX.bl"] <- "Group"
finalResult$Group <-factor(finalResult$Group,levels = c("AD","MCI","CN"))

finalResult[grep("pQTLCSF",finalResult$variable),"analysis"] <- "univariate.pQTL"
finalResult[grep("eQTLBrain",finalResult$variable),"analysis"] <- "univariate.eQTL"


# PANEL PLOT PRIMARY ANALYSIS···············································
plotData.primary <- finalResult[finalResult$analysis=="Primary",]

# Auxiliar
  # Variable and levels to factor
plotData.primary$variable <- factor(plotData.primary$variable,
                                    levels=c(riskVars,"APOE4"))
plotData.primary$PRS_value <- factor(plotData.primary$PRS_value,
                                     levels=c("Low-Intermediate","High",
                                              "0","1","2"))
plotData.primary$Volumes <- factor(plotData.primary$Volumes,levels=volVars)  
  pos_dodge <- position_dodge(width=0.6) 

#  volume.labs <- volVars
  volume.labs <- c("Whole\nhippocampus","Subiculum",
                    "Pre-\nsubiculum","Para-\nsubiculum")
  names(volume.labs) <- volVars
  
  PRS.labs <- sapply(strsplit(c(riskVars,"APOE4"),split="_"),"[",1)
  names(PRS.labs) <- c(riskVars,"APOE4")
  PRS.labs[PRS.labs=="ADnoAPOE"] <- "AD\nno APOE"
  

#PLOT
panel <- 
  ggplot(plotData.primary,aes(x=Group)) +
  geom_hline(yintercept=0,linetype="dashed",color="red")+
  geom_pointrange(aes(y=time_visit.trend , ymin=lower.CL , ymax=upper.CL, 
                      color=as.factor(PRS_value), alpha=alpha),
                  position = pos_dodge, show.legend = F,size=.3,
                  linewidth=.5) +
  geom_text(aes(y=-diff(range(time_visit.trend))/40,
                label=symbolJoinTest),size=10)+
  facet_grid(rows=vars(variable),cols=vars(Volumes),scales = "free",
             labeller=labeller(Volumes=volume.labs, variable=PRS.labs))+
  scale_color_manual(values = c("#D95F02","#7570B3",
                                "#0033CC","#993366","#990000")) +
  scale_alpha(range = c(0.3,1))+
  coord_flip()+
  
  theme_bw() +
  theme(strip.background = element_blank(),
        plot.margin = margin(1,1,.5,1,"cm"),
        axis.title = element_blank(),
        axis.text.x = element_text(angle=90,hjust=1)) +
  ggtitle("APOE carriership and AD PRS effect over time rate of change of hippocampal volumes<br>by diagnostic group.") +
  labs(subtitle = "Adjusted for Age at baseline, sex, intracranial volume, years of education and tesla scan parameter. <br/>
       Trends for <span style = 'color: #D95F02;'>**Low-Intermediate**</span> and <span style = 'color: #7570B3;'>**High**</span> predisposition to PRS trait or disease<br/>
       or for <span style = 'color: #0033CC;'>**0**</span>,<span style = 'color: #993366;'>**1**</span> or
       <span style = 'color: #990000;'>**2**</span> effect alleles",
       caption="Linear coefficient and 95% confidence intervals for volumetric rate of change over time (mm3 change per year),by diagnostic group and level of predisposition<br>
       for the PRS trait/disease. Highlighted estimates indicate significance differences between High and Low-Intermediate predispositions for a particular diagnostic<br>
       group. Panels with an \\* present significant difference in PRS effect on volume change between groups. The red dashed line marks no time trend.<br>
       Estimates correspond to models L.2") +
  theme(plot.caption = element_markdown(hjust = 0, lineheight = 1.5),
        plot.subtitle = element_markdown(size = 12, hjust = 0),
        plot.title = element_markdown(size = 16, hjust = 0))

panel.primary <- panel
panel.primary
#9.86 x 7.58

# PANEL PLOT Univariate brain ANALYSIS···············································
plotData.univar.brain <- finalResult[finalResult$analysis=="univariate.eQTL",]

# Auxiliar
# Variable and levels to factor
plotData.univar.brain$variable <- factor(plotData.univar.brain$variable,
                                      levels=snpVars[2:7])
plotData.univar.brain$PRS_value <- factor(plotData.univar.brain$PRS_value,
                                     levels=c("0","1","2"))

plotData.univar.brain$Volumes <- factor(plotData.univar.brain$Volumes,levels=volVars)  

PRS.labs <- sapply(strsplit(snpVars[2:7],split="\\."),"[",2)
names(PRS.labs) <- snpVars[2:7]

panel.univariate.brain <- 
  ggplot(plotData.univar.brain,aes(x=Group)) +
  geom_hline(yintercept=0,linetype="dashed",color="red")+
  geom_pointrange(aes(y=time_visit.trend , ymin=lower.CL , ymax=upper.CL, 
                      color=as.factor(PRS_value)),
                  position = pos_dodge, show.legend = F,size=.3,
                  linewidth=.5,alpha=0.2) +
  geom_text(aes(y=max(time_visit.trend)+mean(time_visit.trend)/3,
                label=symbolJoinTest),size=10)+
  scale_color_manual(values = c("#0033CC","#993366","#990000")) +
  facet_grid(rows=vars(variable),cols=vars(Volumes),scales = "free",
             labeller=labeller(Volumes=volume.labs, variable=PRS.labs))+
  coord_flip()+
  theme_bw() +
  theme(strip.background = element_blank(),
        plot.margin = margin(1,1,.5,1,"cm"),
        axis.title = element_blank(),
        axis.text.x = element_text(angle=90,hjust=1)) +
  ggtitle("eQTL Brain individual SNPs effect over time rate of change of<br>
          hippocampal volumes by diagnostic group.") +
  labs(subtitle = "Adjusted for Age at baseline, sex, intracranial volume, years of education and tesla<br>
  scan parameter. <br/>
       Trends for <span style = 'color: #0033CC;'>**0**</span>,<span style = 'color: #993366;'>**1**</span> or
       <span style = 'color: #990000;'>**2**</span> effect alleles" ,
       caption="Linear coefficient and 95% confidence intervals for volumetric rate of change over time (mm3 change per year),by<br>
       diagnostic group and level of predisposition for the PRS trait/disease. Highlighted estimates indicate significance<br>
       differences between High and Low-Intermediate predispositions for a particular diagnostic group. Panels with an<br/>
       \\* present significant difference in PRS effect on volume change between groups. The red dashed line marks no<br/>
       time trend. Estimates correspond to models L.2.") +
  theme(plot.caption = element_markdown(hjust = 0, lineheight = 1.5),
        plot.subtitle = element_markdown(size = 12, hjust = 0),
        plot.title = element_markdown(size = 16, hjust = 0))

panel.univariate.brain
#9.86 x 7.58 in , portrait
  

# PANEL PLOT Univariate brain ANALYSIS···············································
plotData.univar.CSF <- finalResult[finalResult$analysis=="univariate.pQTL",]

# Variable and levels to factor
plotData.univar.CSF$variable <- factor(plotData.univar.CSF$variable,
                                         levels=snpVars[8:11])
plotData.univar.CSF$PRS_value <- factor(plotData.univar.CSF$PRS_value,
                                          levels=c("0","1","2","1-2"))

plotData.univar.CSF$Volumes <- factor(plotData.univar.CSF$Volumes,levels=volVars)  

PRS.labs <- sapply(strsplit(snpVars[8:11],split="\\."),"[",2)
names(PRS.labs) <- snpVars[8:11]

panel.univariate.CSF <- 
  panel %+% plotData.univar.CSF +
  scale_color_manual(values = c("#0033CC","#993366","#990000","#990000")) +
  facet_grid(rows=vars(variable),cols=vars(Volumes),scales = "free",
             labeller=labeller(Volumes=volume.labs, variable=PRS.labs)) +
  ggtitle("pQTL CSF individual SNPs effect over time rate of change of hippocampal volumes by diagnostic group.") +
  labs(subtitle = "Adjusted for Age at baseline, sex, intracranial volume, years of education and tesla scan parameter. <br/>
       Trends for <span style = 'color: #0033CC;'>**0**</span>,<span style = 'color: #993366;'>**1**</span> or
       <span style = 'color: #990000;'>**2**</span> effect alleles",
       caption="Linear coefficient and 95% confidence intervals for volumetric rate of change over time (mm3 change per year),by diagnostic group and level of<br>
       predisposition for the PRS trait/disease. Highlighted estimates indicate significance differences between High and Low-Intermediate predispositions<br>
       for a particular diagnostic group. Panels with an \\* present significant difference in PRS effect on volume change between groups. The red dashed<br>
       line marks no time trend. Estimates correspond to models L.2") +
  theme(text = element_text(size=15))
panel.univariate.CSF

#12 x 10 in , landscape


# path <- here("results",paramT,"2_Modelling","2_longitudinalTrajectories",
#              "modelL2_plots.RData")
# save(list = list(panel.primary,panel.univariate.brain, panel.univariate.CSF),
#      file = path)


# library(ggeffects)
# plotModels <- function(model){
#   
#   fit <- model$mod_3wayIntDX
#   prs <- attributes(fit)$PRS
#   volume <- attributes(fit)$volume
#   
#   # Plots
#   m <- ggpredict(fit,terms=c("time_visit",prs,"DX.bl"),
#                  condition = c(ICV=mean(dd2$ICV)))
#   
#   p <- plot(m,add.data = T,ci.style = "dash",dot.alpha = 0.2,dot.size = 1.5,
#             colors = c("#D95F02","#7570B3"),line.size = 1,use.theme = T)
#   
#   # Save
#   path <-here("results",paramT,"2_Modelling","2_longitudinalTrajectories",
#               "trajectories","time_visit")
#   filename <- paste0("modelTimeVisit_",volume,"_",prs,".png")
#   ggsave(plot=p,filename = filename,path=path,device = "png")
# 
#   # Return
#   return(plot=p)
# }
# 
# plots<- lapply(result,FUN=plotModels)


#-------------------------------------------------------------------------------
# B) OBJ2.bis
#    PRS effect on volume reduction, depending on disease status
#    (coeficient beta_timeVisit:PRS) 
#    Stratified models too.
#-------------------------------------------------------------------------------
volume <- "Whole_hippocampus" ; PRS <- "AD_predisposition" ; group <- "All"
data <- dd2

#--------------------------------------------------------------------------------

prsTimeEffect_strat <- function(volume,PRS,group = "All",data){
  data_strat <- data
  
  # i) Formula ··················································
  predictors <- paste("time_visit",c("DX.bl",PRS),sep=" * ")
  covars <- c("AGE","I(ICV/1000)","PTGENDER","PTEDUCAT","tesla")
  random <- "(1+time_visit|PTID)"
  
  if(group %in% c("CN","MCI","AD")){
    data_strat <- subset(data,DX.bl == group)
    predictors <- paste("time_visit",PRS,sep=" * ")
  }
  
  if(group %in% c("Female","Male")){
    data_strat <- subset(data,PTGENDER == group)
    covars <-covars[!(covars=="PTGENDER")]
  }
  
  
  form <- paste(c(predictors,covars,random),collapse= " + ")
  form <- paste(volume,form, sep = " ~ ")
  
  mod <- suppressMessages(
    lme4::lmer(formula = form, data=data_strat,
               control = lmerControl(optimizer ="Nelder_Mead")))
  

  if(isSingular(mod)){
    cat("Singular random effects covariance matrix for ",volume," and ",PRS," in ",group,"strata.
Removing random slope to simplify model...\n\n")
    random <- "(1|PTID)"
    form <- paste(c(predictors,covars,random),collapse= " + ")
    form <- paste(volume,form, sep = " ~ ")
    mod <- lme4::lmer(formula = form, data=data_strat,
                      control = lmerControl(optimizer ="Nelder_Mead"))
  }

  fixef <- extract_fixed_effects(mod)
  
  effect_pos <- grep(paste0("^time_visit:",PRS),fixef$term)
  
  res <- as.data.frame(fixef[effect_pos,c("value","se","lower_2.5",
                                          "upper_97.5","p_value")])
  
  rownames(res) <- paste(volume,PRS,group,sep="|")
  
  return(list("PRS_time"=res,"model"=mod))
}
#--------------------------------------------------------------------------

modGridDx <- expand.grid(PRS = c(riskVars,snpVars),
                         Volume = volVars,
                         Group = c("All",levels(dd2$DX.bl)),
                         stringsAsFactors = F)

modGridDx$analysis[modGridDx$PRS %in% c(riskVars,"APOE4")] <- "Primary"
modGridDx$analysis[modGridDx$PRS %in% snpVars[-1]] <- "Secondary"
# modGridDx$analysis[modGridDx$PRS %in% snpVars[2:7]] <- "Univariate.brain"
# modGridDx$analysis[modGridDx$PRS %in% snpVars[8:11]] <- "Univariate.CSF"

result_stratDx <- mapply(FUN = prsTimeEffect_strat,
                         volume = modGridDx$Volume,
                         PRS = modGridDx$PRS,
                         group = modGridDx$Group,
                         MoreArgs = list(data=dd2), 
                         SIMPLIFY = F,USE.NAMES = F)

res <- lapply(result_stratDx,"[[","PRS_time")
res <- do.call(rbind,res) # consolidar en dataFrame
res <- cbind(modGridDx,res) # Afegir PRS i volume 
rownames(res) <- NULL # no rownames
res$PRS <- factor(res$PRS,levels=c(riskVars,snpVars)) 
res$Group <- factor(res$Group,c("All","CN","MCI","AD"))
head(res)
res <- split(res,res$Group)


lr <- lapply(res, function(l){
  l$prsVol <- paste(l$PRS,l$Volume,sep="|")
  aux <- lapply(split(l,l$analysis), function(x){
    x$pvalue_corrected <- p.adjust(x$p_value, method = "fdr")
    return(x[c("prsVol","pvalue_corrected")])
  })
  aux <- do.call(rbind,aux)
  pos <- match(l$prsVol,aux$prsVol)
  l$pvalue_corrected <- aux[pos,"pvalue_corrected"]
  l$prsVol <- NULL
  return(l)
})
res <- do.call(rbind,lr)

res[paste(names(res)[9:10],"symbol",sep="_")] <-
  lapply(res[names(res[9:10])], FUN=cut,
         breaks = c(-Inf,0.001,0.01,0.05,0.1,1),
         labels=c("***","**","*","·",""))

res_prsTimeEffect_stratDx <- res
head(res_prsTimeEffect_stratDx)
(significant_effectsDx <- subset(res_prsTimeEffect_stratDx, 
                                 pvalue_corrected_symbol!=""))


### PLOT
# plotDataDx <- subset(res_prsTimeEffect_stratDx,
#                      Volume!="Whole_hippocampus" & res_prsTimeEffect_stratDx$Group!="All")
# plotDataDx <- subset(res_prsTimeEffect_stratDx,res_prsTimeEffect_stratDx$Group!="All")
# plotDataDx <- plotDataDx[plotDataDx$analysis=="Primary",]
# 
# upper.th <- quantile(plotDataDx$value,.9)
# lower.th <- quantile(plotDataDx$value,.1)
# 
# plotDataDx$Volume <- factor(plotDataDx$Volume,levels=unique(plotDataDx$Volume),
#                             labels=c("CA1","Presubiculum","Subiculum","Whole\nhippocampus"))
# 
# 
# heatMapDx <- 
#   ggplot(data=plotDataDx, aes(x=Volume, y=PRS, 
#                               fill=pmax(pmin(value,upper.th),lower.th))) + 
#   geom_tile(color="grey") +
#   geom_text(aes(label=pvalue_corrected_symbol),size=4)+
#   scale_fill_gradient2(low="red4",mid = "white", high="#0066FF", midpoint = 0,
#                       name="beta")+
#   facet_wrap(~Group,ncol=3)+
#   ggstatsplot::theme_ggstatsplot() +
#   theme(strip.background = element_blank(),
#         plot.margin = margin(1,1,.5,1,"cm"),
#         axis.title = element_blank(),
#         axis.text = element_text(face="bold",size = 8),
#         axis.text.x = element_text(angle=90,vjust = 0.6,hjust = 1)) +
#   
#   labs(caption="Linear coefficient for interaction between time and PRS. Effects adjusted for Age at baseline (years), intracranial volume(cm3), sex and years<br/>
#        of education. Model allows a diferent volume rate of change depending on diagnositc status. Also, intercept and rate of change in time<br/>
#   are included as random effects. Significant interactions are reported as Wald-test pvalues at levels 0.1(·), 0.05(\\*), 0.01(\\**) and 0.001(\\***),<br/>
#   corrected for multiple comparisons by FDR method. Non-significant differences are not shown.<br/>
#   **Source**: ADNI project | **Units** Average increment in volume rate of change (mm3) associated with having high predisposition for the PRS trait.") +
#   theme(plot.caption = element_markdown(hjust = 0, lineheight = 1.5),
#         plot.subtitle = element_markdown(size = 12, hjust = 0),
#         plot.title = element_text(size = 16, hjust = 0))
# 
# heatMapDx


# ALL
### PLOT 
# plotDataDx <- subset(res_prsTimeEffect_stratDx,
#                      Volume!="Whole_hippocampus" & res_prsTimeEffect_stratDx$Group!="All")
plotDataDx <- res_prsTimeEffect_stratDx
plotDataDx <- plotDataDx[plotDataDx$analysis=="Primary",]

upper.th <- quantile(plotDataDx$value,1)
lower.th <- quantile(plotDataDx$value,0.01)

plotDataDx$Volume <- factor(plotDataDx$Volume,levels=unique(plotDataDx$Volume),
                            labels=c("Whole\nhippocampus","Subiculum","Pre-\nsubiculum",
                                     "Para-\nsubiculum"))


heatMapDx <- 
  ggplot(data=plotDataDx, aes(x=Volume, y=PRS, 
                              fill=pmax(pmin(value,upper.th),lower.th))) + 
  geom_tile(color="grey") +
  geom_text(aes(label=pvalue_corrected_symbol),size=4)+
  scale_fill_gradient2(low="red4",mid = "white", high="#0066FF", midpoint = 0,
                       name="Effect")+
  facet_wrap(~Group,ncol=2)+
  ggstatsplot::theme_ggstatsplot() +
  theme(strip.background = element_blank(),
        plot.margin = margin(1,1,.5,1,"cm"),
        axis.title = element_blank(),
        axis.text = element_text(face="bold",size = 8)) +
  
  labs(caption="Linear coefficient for interaction between time and PRS. Effects adjusted for Age at baseline (years), intracranial volume(cm3),<br/>
  sex, years of education and tesla parameter. Model allows a diferent volume rate of change depending on diagnositc status.<br/>
  Also, intercept and rate of change in time are included as random effects. Significant interactions are reported as Wald-test<br/>
  pvalues at levels 0.1(·), 0.05(\\*), 0.01(\\**) and 0.001(\\***), corrected for multiple comparisons by FDR method. Non-significant<br/>
  differences are not shown.<br/>
  **Units** Average increment in volume rate of change (mm3) associated with having high predisposition for the PRS trait.") +
  theme(plot.caption = element_markdown(hjust = 0, lineheight = 1.5),
        plot.subtitle = element_markdown(size = 12, hjust = 0),
        plot.title = element_text(size = 16, hjust = 0))


heatMapDx.primary <-heatMapDx
heatMapDx.primary
#8.92x5.80

# Secondary
plotDataDx <- res_prsTimeEffect_stratDx
plotDataDx <- plotDataDx[plotDataDx$analysis=="Secondary",]

upper.th <- quantile(plotDataDx$value,.99)
lower.th <- quantile(plotDataDx$value,0.01)

plotDataDx$Volume <- factor(plotDataDx$Volume,levels=unique(plotDataDx$Volume),
                            labels=c("Whole\nhippocampus","Subiculum","Pre-\nsubiculum",
                                     "Para-\nsubiculum"))


heatMapDx <- 
  ggplot(data=plotDataDx, aes(x=Volume, y=PRS, 
                              fill=pmax(pmin(value,upper.th),lower.th))) + 
  geom_tile(color="grey") +
  geom_text(aes(label=pvalue_corrected_symbol),size=4)+
  scale_fill_gradient2(low="red4",mid = "white", high="#0066FF", midpoint = 0,
                       name="Effect")+
  facet_wrap(~Group,ncol=2)+
  ggstatsplot::theme_ggstatsplot() +
  theme(strip.background = element_blank(),
        plot.margin = margin(1,1,.5,1,"cm"),
        axis.title = element_blank(),
        axis.text = element_text(face="bold",size = 8)) +
  
  labs(caption="Linear coefficient for interaction between time and SNP. Effects adjusted for Age at baseline (years), intracranial volume(cm3),<br/>
  sex, years of education and tesla parameter. Model allows a diferent volume rate of change depending on diagnositc status.<br/>
  Also, intercept and rate of change in time are included as random effects. Significant interactions are reported as Wald-test<br/>
  pvalues at levels 0.1(·), 0.05(\\*), 0.01(\\**) and 0.001(\\***), corrected for multiple comparisons by FDR method. Non-significant<br/>
  differences are not shown. All SNP effects are parameterized according to additive model, except for rs_7412_T with a dominant<br/>
  model. **Units** Average increment in volume rate of change (mm3) associated with an increase of 1 alternative allele.") +
  theme(plot.caption = element_markdown(hjust = 0, lineheight = 1.5),
        plot.subtitle = element_markdown(size = 12, hjust = 0),
        plot.title = element_text(size = 16, hjust = 0))


heatMapDx.secondary <-heatMapDx
heatMapDx.secondary
#8.92x6.80

### Plot primary - Whole hippocampus
plotDataDx <- res_prsTimeEffect_stratDx
plotDataDx <- plotDataDx[plotDataDx$analysis=="Primary" &
                         plotDataDx$Volume=="Whole_hippocampus" ,]

upper.th <- quantile(plotDataDx$value,1)
lower.th <- quantile(plotDataDx$value,0)


heatMapDx <- 
  ggplot(data=plotDataDx, aes(x=Group, y=PRS, 
                              fill=pmax(pmin(value,upper.th),lower.th))) + 
  geom_tile(color="grey") +
  geom_text(aes(label=pvalue_corrected_symbol),size=4)+
  scale_fill_gradient2(low="red4",mid = "white", high="#0066FF", midpoint = 0,
                       name="Effect")+
  ggstatsplot::theme_ggstatsplot() +
  theme(strip.background = element_blank(),
        plot.margin = margin(1,1,.5,1,"cm"),
        axis.title = element_blank(),
        axis.text = element_text(face="bold",size = 8)) +
  
  # labs(caption="Linear coefficient for interaction between time and PRS. Effects adjusted for Age at baseline (years), intracranial volume(cm3),<br/>
  # sex, years of education and tesla parameter. Model allows a diferent volume rate of change depending on diagnositc status.<br/>
  # Also, intercept and rate of change in time are included as random effects. Significant interactions are reported as Wald-test<br/>
  # pvalues at levels 0.1(·), 0.05(\\*), 0.01(\\**) and 0.001(\\***), corrected for multiple comparisons by FDR method. Non-significant<br/>
  # differences are not shown.<br/>
  # **Units** Average increment in volume rate of change (mm3) associated with having high predisposition for the PRS trait.") +
  theme(plot.caption = element_markdown(hjust = 0, lineheight = 1.5),
        plot.subtitle = element_markdown(size = 12, hjust = 0),
        plot.title = element_text(size = 16, hjust = 0))


heatMapDx.primary.WH <-heatMapDx
heatMapDx.primary.WH

# Export 7ix3i 


# Secondary - WH
plotDataDx <- res_prsTimeEffect_stratDx
plotDataDx <- plotDataDx[plotDataDx$analysis=="Secondary" & 
                           plotDataDx$Volume == "Whole_hippocampus",]

upper.th <- quantile(plotDataDx$value,.99)
lower.th <- quantile(plotDataDx$value,0.01)

# plotDataDx$Volume <- factor(plotDataDx$Volume,levels=unique(plotDataDx$Volume),
#                             labels=c("Whole\nhippocampus","Subiculum","Pre-\nsubiculum",
#                                      "Para-\nsubiculum"))


heatMapDx <- 
  ggplot(data=plotDataDx, aes(x=Group, y=PRS, 
                              fill=pmax(pmin(value,upper.th),lower.th))) + 
  geom_tile(color="grey") +
  geom_text(aes(label=pvalue_corrected_symbol),size=4)+
  scale_fill_gradient2(low="red4",mid = "white", high="#0066FF", midpoint = 0,
                       name="beta")+
  # facet_wrap(~Group,ncol=2)+
  ggstatsplot::theme_ggstatsplot() +
  theme(strip.background = element_blank(),
        plot.margin = margin(1,1,.5,1,"cm"),
        axis.title = element_blank(),
        axis.text = element_text(face="bold",size = 8)) +
  
  # labs(caption="Linear coefficient for interaction between time and SNP. Effects adjusted for Age at baseline (years), intracranial volume(cm3),<br/>
  # sex, years of education and tesla parameter. Model allows a diferent volume rate of change depending on diagnositc status.<br/>
  # Also, intercept and rate of change in time are included as random effects. Significant interactions are reported as Wald-test<br/>
  # pvalues at levels 0.1(·), 0.05(\\*), 0.01(\\**) and 0.001(\\***), corrected for multiple comparisons by FDR method. Non-significant<br/>
  # differences are not shown. All SNP effects are parameterized according to additive model, except for rs_7412_T with a dominant<br/>
  # model. **Units** Average increment in volume rate of change (mm3) associated with an increase of 1 alternative allele.") +
  theme(plot.caption = element_markdown(hjust = 0, lineheight = 1.5),
        plot.subtitle = element_markdown(size = 12, hjust = 0),
        plot.title = element_text(size = 16, hjust = 0))


heatMapDx.secondary.WH <-heatMapDx
heatMapDx.secondary.WH
#7x3 landscape


### Plot primary - subfields

plotDataDx <- res_prsTimeEffect_stratDx
plotDataDx <- plotDataDx[plotDataDx$analysis=="Primary" &
                         plotDataDx$Volume !="Whole_hippocampus",]

upper.th <- quantile(plotDataDx$value,1)
lower.th <- quantile(plotDataDx$value,0)

plotDataDx$Volume <- factor(plotDataDx$Volume,levels=unique(plotDataDx$Volume),
                            labels=c("Subiculum","Pre-\nsubiculum","Para-\nsubiculum"))


heatMapDx <- 
  ggplot(data=plotDataDx, aes(x=Volume, y=PRS, 
                              fill=pmax(pmin(value,upper.th),lower.th))) + 
  geom_tile(color="grey") +
  geom_text(aes(label=pvalue_corrected_symbol),size=4)+
  scale_fill_gradient2(low="red4",mid = "white", high="#0066FF", midpoint = 0,
                       name="Effect")+
  facet_wrap(~Group,ncol=2)+
  ggstatsplot::theme_ggstatsplot() +
  theme(strip.background = element_blank(),
        plot.margin = margin(1,1,.5,1,"cm"),
        axis.title = element_blank(),
        axis.text = element_text(face="bold",size = 8)) +
  
  # labs(caption="Linear coefficient for interaction between time and PRS. Effects adjusted for Age at baseline (years), intracranial volume(cm3),<br/>
  # sex, years of education and tesla parameter. Model allows a diferent volume rate of change depending on diagnositc status.<br/>
  # Also, intercept and rate of change in time are included as random effects. Significant interactions are reported as Wald-test<br/>
  # pvalues at levels 0.1(·), 0.05(\\*), 0.01(\\**) and 0.001(\\***), corrected for multiple comparisons by FDR method. Non-significant<br/>
  # differences are not shown.<br/>
  # **Units** Average increment in volume rate of change (mm3) associated with having high predisposition for the PRS trait.") +
  theme(plot.caption = element_markdown(hjust = 0, lineheight = 1.5),
        plot.subtitle = element_markdown(size = 12, hjust = 0),
        plot.title = element_text(size = 16, hjust = 0))


heatMapDx.primary.subfields <-heatMapDx
heatMapDx.primary.subfields
#8.7x5.5 in  export pdf

# Plot secondary subfields
plotDataDx <- res_prsTimeEffect_stratDx
plotDataDx <- plotDataDx[plotDataDx$analysis=="Secondary" & 
                         plotDataDx$Volume!="Whole_hippocampus",]

upper.th <- quantile(plotDataDx$value,1)
lower.th <- quantile(plotDataDx$value,00)

plotDataDx$Volume <- factor(plotDataDx$Volume,levels=unique(plotDataDx$Volume),
                            labels=c("Subiculum","Pre-\nsubiculum","Para-\nsubiculum"))


heatMapDx <- 
  ggplot(data=plotDataDx, aes(x=Volume, y=PRS, 
                              fill=pmax(pmin(value,upper.th),lower.th))) + 
  geom_tile(color="grey") +
  geom_text(aes(label=pvalue_corrected_symbol),size=4)+
  scale_fill_gradient2(low="red4",mid = "white", high="#0066FF", midpoint = 0,
                       name="Effect")+
  facet_wrap(~Group,ncol=2)+
  ggstatsplot::theme_ggstatsplot() +
  theme(strip.background = element_blank(),
        plot.margin = margin(1,1,.5,1,"cm"),
        axis.title = element_blank(),
        axis.text = element_text(face="bold",size = 8)) +
  
  # labs(caption="Linear coefficient for interaction between time and SNP. Effects adjusted for Age at baseline (years), intracranial volume(cm3),<br/>
  # sex, years of education and tesla parameter. Model allows a diferent volume rate of change depending on diagnositc status.<br/>
  # Also, intercept and rate of change in time are included as random effects. Significant interactions are reported as Wald-test<br/>
  # pvalues at levels 0.1(·), 0.05(\\*), 0.01(\\**) and 0.001(\\***), corrected for multiple comparisons by FDR method. Non-significant<br/>
  # differences are not shown. All SNP effects are parameterized according to additive model, except for rs_7412_T with a dominant<br/>
  # model. **Units** Average increment in volume rate of change (mm3) associated with an increase of 1 alternative allele.") +
  theme(plot.caption = element_markdown(hjust = 0, lineheight = 1.5),
        plot.subtitle = element_markdown(size = 12, hjust = 0),
        plot.title = element_text(size = 16, hjust = 0))


heatMapDx.secondary.subfields <-heatMapDx
heatMapDx.secondary.subfields
#8.7x5.5 in  export pdf

# PRS/SNP·time_visit
tab <- res_prsTimeEffect_stratDx
rownames(tab) <- NULL
tab <- split(tab,tab$Group)
tab <- lapply(tab,function(x)x[,!names(x)=="Group"])

# Build Excel worksheet
wb <- createWorkbook()
invisible(lapply(names(tab),function(x)addWorksheet(wb,x)))
invisible(
  lapply(1:length(tab),
         function(i) writeDataTable(wb = wb,
                                    x = tab[[i]],
                                    sheet = names(tab)[i],
                                    withFilter = T,
                                    tableStyle = "TableStyleLight1")
  )
)

path <- here("results",paramT,"2_Modelling","2_longitudinalTrajectories",
             "taules","modelL1_interactionTimeVsGenetics_DXstratified.xlsx")
saveWorkbook(wb,file=path,overwrite = T)


#-------------------------------------------------------------------------------
# B) OBJ3
#    Does the effect of PRS on volume reduction depend on sex?
#-------------------------------------------------------------------------------
##### STRATIFIED BY SEX
# modGridSx <- expand.grid(PRS = c(riskVars,snpVars),
#                          Volume = sort(volVars),
#                          Group = c("All",levels(dd2$PTGENDER)),
#                          stringsAsFactors = F)
# 
# modGridSx$analysis[modGridSx$PRS %in% riskVars[1:4]] <- "Primary"
# modGridSx$analysis[modGridSx$PRS %in% c(riskVars[5:6],"APOE4")] <- "Secondary"
# modGridSx$analysis[modGridSx$PRS %in% snpVars[2:7]] <- "Univariate.brain"
# modGridSx$analysis[modGridSx$PRS %in% snpVars[8:11]] <- "Univariate.CSF"
# 
# 
# result_stratSx <- mapply(FUN = prsTimeEffect_strat,
#                          volume = modGridSx$Volume,
#                          PRS = modGridSx$PRS,
#                          group = modGridSx$Group,
#                          MoreArgs = list(data=dd2), 
#                          SIMPLIFY = F,USE.NAMES = F)
# 
# # i arreglem el resultat...
# res <- lapply(result_stratSx,"[[","PRS_time")
# res <- do.call(rbind,res) # consolidar en dataFrame
# res <- cbind(modGridSx,res) # Afegir PRS i volume 
# rownames(res) <- NULL # no rownames
# res$PRS <- factor(res$PRS,levels=c(riskVars,snpVars)) 
# res$Group <- factor(res$Group,c("All","Female","Male"))
# head(res)
# res <- split(res,res$Group)
# 
# lr <- lapply(res, function(l){
#   l$prsVol <- paste(l$PRS,l$Volume,sep="|")
#   aux <- lapply(split(l,l$analysis), function(x){
#     x$pvalue_corrected <- p.adjust(x$p_value, method = "fdr")
#     return(x[c("prsVol","pvalue_corrected")])
#   })
#   aux <- do.call(rbind,aux)
#   pos <- match(l$prsVol,aux$prsVol)
#   l$pvalue_corrected <- aux[pos,"pvalue_corrected"]
#   l$prsVol <- NULL
#   return(l)
# })
# res <- do.call(rbind,lr)
# 
# res[paste(names(res)[9:10],"symbol",sep="_")] <-
#   lapply(res[names(res[9:10])], FUN=cut,
#          breaks = c(-Inf,0.001,0.01,0.05,0.1,1),
#          labels=c("***","**","*","·",""))
# 
# 
# res_prsTimeEffect_stratSx <- res
# head(res_prsTimeEffect_stratSx)
# (significant_effectsSx <- subset(res_prsTimeEffect_stratSx, 
#                                  pvalue_corrected_symbol!=""))
# 
# 
# ### PLOT 
# plotDataSx <- subset(res_prsTimeEffect_stratSx,
#                      Volume!="Whole_hippocampus" & res_prsTimeEffect_stratSx$Group!="All")
# 
# heatMapSx <- 
#   ggplot(data=plotDataSx, aes(x=Volume, y=PRS, fill=value)) + 
#   geom_tile() +
#   geom_text(aes(label=pvalue_corrected_symbol),size=4)+
#   scale_fill_gradient2(low="red4",mid = "white", high="#0066FF", midpoint = 0)+
#   facet_wrap(~Group,ncol=2)+
#   ggstatsplot::theme_ggstatsplot() +
#   theme(strip.background = element_blank(),
#         plot.margin = margin(1,1,.5,1,"cm"),
#         axis.title = element_blank(),
#         axis.text = element_text(face="bold",size = 8),
#         axis.text.x = element_text(angle=90,vjust = 0.6,hjust = 1)) +
#   
#   ggtitle("Corrected significant effect of investigated PRS on hippocampal\nsubfield volume's rate of change") +
#   labs(subtitle="Stratified by sex",
#        caption="Linear coefficient for interaction between time and PRS. Effects adjusted for Age at baseline (years), intracranial volume(cm3), diagnostic status and years<br/>
#        of education. Model allows a diferent volume rate of change depending on diagnositc status. Also, intercept and rate of change in time<br/>
#   are included as random effects. Significant interactions are reported as Wald-test pvalues at levels 0.1(·), 0.05(\\*), 0.01(\\**) and 0.001(\\***),<br/>
#   corrected for multiple comparisons by FDR method. Non-significant differences are not shown.<br/>
#   **Source**: ADNI project | **Units** Average increment in volume rate of change (mm3) associated with having high predisposition for the PRS trait.") +
#   theme(plot.caption = element_markdown(hjust = 0, lineheight = 1.5),
#         plot.subtitle = element_markdown(size = 12, hjust = 0),
#         plot.title = element_text(size = 16, hjust = 0))
# 
# heatMapSx
# 
# 
# tab <- res_prsTimeEffect_stratSx
# rownames(tab) <- NULL
# tab <- split(tab,tab$Group)
# tab <- lapply(tab,function(x)x[,!names(x)=="Group"])
# 
# # Build Excel worksheet
# wb <- createWorkbook()
# invisible(lapply(names(tab),function(x)addWorksheet(wb,x)))
# invisible(
#   lapply(1:length(tab),
#          function(i) writeDataTable(wb = wb,
#                                     x = tab[[i]],
#                                     sheet = names(tab)[i],
#                                     withFilter = T,
#                                     tableStyle = "TableStyleLight1")
#   )
# )
# 
# path <- here("results",paramT,"2_Modelling","2_longitudinalTrajectories",
#              "taules","modelL1_interactionTimeVsGenetics_Sxstratified.xlsx")
# saveWorkbook(wb,file=path,overwrite = T)

# rm(list=ls())
# source("Rscripts/libraries.R")
# source(here(RscriptsDir,"3_modelling","modelling_auxFunctions.R"))
# dd2 <- readRDS(here(cleandataDir,"datasetForAnalysis.long.rds"))
# result <- readRDS(here(cleandataDir,"results.rds"))
# load(here(cleandataDir,"vars.RData"))

testSx <- sapply(result,"[[","AnovaTest_3wayIntSx")
cbind(testSx[testSx<0.05])

### LINEAR TRENDSlow or high genetic predisposition
# Debbuging:x<-result$`Hippocampal_tail:AD_predisposition` 


trendsSx <- lapply(result,function(x){
  mod <- x$mod_3wayIntSx
  PRS <- attributes(mod)$PRS
  specs <- as.formula(paste0("~",PRS,"*PTGENDER"))
  #at <- "list(DX.bl = 'MCI')"
  at <- list()
  if(PRS %in% snpVars){
    levs <- paste0(names(table(dd2[,PRS])),collapse=",")
    at <- paste0("list(DX.bl = 'MCI', ",
                 PRS," = c(",levs,"))")
  }
  if(PRS == "pQTLCSF.rs7412_T"){
    at <- paste0("list(DX.bl = 'MCI', ",
                 PRS," = c('0','1-2'))")
  }
  
  trend <- emtrends(mod, specs = specs, "time_visit",
                    at = eval(parse(text=at)),
                    pbkrtest.limit=4186,weights="prop")
  
  return(trend)
})


trendsSx.df <- lapply(trendsSx,summary)

trendsSx.df <- lapply(trendsSx.df,function(x){
  x$variable <- names(x)[1]
  names(x)[1] <- "PRS_value"
  x$PRS_value <- as.character(x$PRS_value)
  x <- x[c("variable","PRS_value",names(x)[-c(1,length(names(x)))])]
  return(x)
})

trendsSx.df <- do.call(rbind,trendsSx.df)
vol <- sapply(strsplit(rownames(trendsSx.df),":"),"[[",1)
trendsSx.df <- cbind(Volumes = vol,trendsSx.df)
rownames(trendsSx.df) <- NULL


contrastPRS.Sx <- lapply(trendsSx,function(x){
  n <- names(x@levels)[1]
  s <- summary(contrast(x,"consec",simple=n))
  if(nrow(s)>2)
  {
    s <- do.call(rbind,lapply(split(s,s$PTGENDER),"[",1,)) 
  }
  # Get volum and PRS variable
  Volume <- as.character(x@model.info$call[[2]])[2]
  PRS <- n
  s <- cbind(Volume,PRS,s)
  return(s)
})

res_contrastPRS.Sx <- as.data.frame(do.call(rbind,contrastPRS.Sx))
rownames(res_contrastPRS.Sx) <- NULL


res_contrastPRS.Sx$analysis <- modGrid[match(res_contrastPRS.Sx$PRS,modGridDx$PRS),"analysis"]
res_contrastPRS.Sx$prsVol <- paste(res_contrastPRS.Sx$PRS,
                                   res_contrastPRS.Sx$Volume,
                                   res_contrastPRS.Sx$PTGENDER,
                                   sep="|")

aux <- lapply(split(res_contrastPRS.Sx,res_contrastPRS.Sx$analysis), function(x){
  x$pvalue_corrected <- p.adjust(x$p.value, method = "fdr")
  return(x[c("prsVol","p.value","pvalue_corrected")])
})
aux <- do.call(rbind,aux)
pos <- match(res_contrastPRS.Sx$prsVol,aux$prsVol)
res_contrastPRS.Sx$pvalue_corrected <- aux[pos,"pvalue_corrected"]
#res_contrastPRS$prsVol <- NULL


res_contrastPRS.Sx$pvalue_corrected_symbol <-
  cut(res_contrastPRS.Sx$pvalue_corrected,
      breaks = c(-Inf,0.001,0.01,0.05,0.1,1),
      labels=c("***","**","*","·",""))

head(res_contrastPRS.Sx)

contrast1_signif.Sx <- subset(res_contrastPRS.Sx,pvalue_corrected<0.05)
prsDiff.Sx <- with(contrast1_signif.Sx,paste(Volume,PRS,sep=":"))


contrast1_signif.Sx[c("prsVol","analysis")] <- NULL
contrast1_signif.Sx[5:8] <- lapply(contrast1_signif.Sx[5:8],round,2)
contrast1_signif.Sx[c(9:10)] <- lapply(contrast1_signif.Sx[c(9:10)],function(x){
  ifelse(x<0.001,"<0.001",sprintf("%.3f",x))
})

contrast1_signif.Sx$pvalue_corrected <- 
  with(contrast1_signif.Sx,paste0(pvalue_corrected,"(",pvalue_corrected_symbol,")"))

rownames(contrast1_signif.Sx) <- NULL
contrast1_signif.Sx$pvalue_corrected_symbol <- NULL

tab <- kbl(contrast1_signif.Sx,align = "lllccccccc",row.names = F,
           caption = "Significant effects of PRS on time trend. Models L.3, linear contrasts 1")


tab <- kable_classic(tab,full_width=F,font="Cambria")
formattedTab <- footnote(tab,
                         general = c("Wald test for $H_0: \\beta5_{time:High} + \\beta7_{time:High:MCI}=0$.", 
                                     "Pvalues were corrected for multiple comparisons using FDR method. Significance level: (*)<.05, (**)<.01, (***)<.001"),
                         title_format = c("bold"))

formattedTab_contrast1.Sx <- row_spec(formattedTab,0,bold=T)
formattedTab_contrast1.Sx

# save_kable(x = formattedTab_contrast1,
#            file = here("results",paramT,"2_Modelling",
#                        "2_longitudinalTrajectories","taules",
#                        "modelL2_contrast1Signif.png"),zoom=1.5,)

# Exportar excel contrastos 1
tab <- res_contrastPRS.Sx
rownames(tab) <- NULL
tab <- split(tab,tab$Volume)

# Build Excel worksheet
wb <- createWorkbook()
invisible(lapply(names(tab),function(x)addWorksheet(wb,x)))
invisible(
  lapply(1:length(tab),
         function(i) writeDataTable(wb = wb,
                                    x = tab[[i]],
                                    sheet = names(tab)[i],
                                    withFilter = T,
                                    tableStyle = "TableStyleLight1")
  )
)

path <- here("results",paramT,"2_Modelling","2_longitudinalTrajectories",
             "taules","modelL3_contrast1_Sex.xlsx")
saveWorkbook(wb,file=path,overwrite = T)


aux <- lapply(prsDiff.Sx,function(x){
  
  linfct <- contrast(trendsSx[[x]],interaction=c("consec","pairwise"))@linfct
  mod <- result[[x]]$mod_3wayIntSx
  joinTest <- summary(glht(mod,linfct),test=Chisqtest())
  
  res <- data.frame(Volumes = attributes(mod)$volume,
                    variable = attributes(mod)$PRS,
                    W2_chisq = as.numeric(joinTest$test$SSH),
                    df = as.numeric(joinTest$test$df[[1]]),
                    Pval = joinTest$test$pvalue)
  res$symbolJoinTest <- ifelse(res$Pval<0.1,"*","")
  return(res)
})
contrast2.Sx <- do.call(rbind,aux)

tab <- contrast2.Sx
tab$W2_chisq <- round(tab$W2_chisq,2)
tab$Pval <- ifelse(tab$Pval<0.001,"<0.001",sprintf("%.3f",tab$Pval))
tab$Pval <- with(tab,ifelse(symbolJoinTest=="*",paste0(Pval,"(*)"),Pval))
tab$symbolJoinTest <- NULL

tab <- kbl(tab,align = "llccc",row.names = F,
           caption = "Differences between diagnostic groups regarding PRS effects on volume rate of change. Models L2, linear contrasts 2")


tab <- kable_classic(tab,full_width=F,font="Cambria")
formattedTab <- footnote(tab,
                         general = c("Simultaneous Wald Test for 3 linear contrast. $H_0:\\beta7_{time:MCI:High}=0 ;\\beta7_{time:AD:High}=0; \\beta7_{time:AD:High}-\\beta7_{time:MCI:High}=0$",
                                     "Significance level: (*)<.05"),
                         title_format = c("bold"))

formattedTab_contrast2.Sx <- row_spec(formattedTab,0,bold=T)
formattedTab_contrast2.Sx

tab <- contrast2.Sx
tab <- split(tab,tab$Volume)

# Build Excel worksheet
wb <- createWorkbook()
invisible(lapply(names(tab),function(x)addWorksheet(wb,x)))
invisible(
  lapply(1:length(tab),
         function(i) writeDataTable(wb = wb,
                                    x = tab[[i]],
                                    sheet = names(tab)[i],
                                    withFilter = T,
                                    tableStyle = "TableStyleLight1")
  )
)

path <- here("results",paramT,"2_Modelling","2_longitudinalTrajectories",
             "taules","modelL3_contrast2.xlsx")
saveWorkbook(wb,file=path,overwrite = T)


finalResultSx <- trendsSx.df
finalResultSx$ident <- with(finalResultSx,
                          paste(variable,Volumes,PTGENDER,sep="|"))

pos <- match(finalResultSx$ident,res_contrastPRS.Sx$prsVol)
finalResultSx$pvalue <- res_contrastPRS.Sx[pos,"p.value"]
finalResultSx$pvalue_corrected <- res_contrastPRS.Sx[pos,"pvalue_corrected"]


finalResultSx$alpha <- with(finalResultSx,ifelse(pvalue_corrected<0.05,1,.9))
#finalResultSx$alpha2 <- with(finalResultSx,ifelse(pvalue<0.05,1,.9))


# JoinTest
finalResultSx$ident <- with(finalResultSx,
                          paste(variable,Volumes,sep="|"))
pos <- match(finalResultSx$ident,paste(contrast2.Sx$variable,contrast2.Sx$Volumes,sep="|"))
finalResultSx$symbolJoinTest <- contrast2.Sx[pos,"symbolJoinTest"]

finalResultSx$symbolJoinTest[is.na(finalResultSx$symbolJoinTest)] <- ""

finalResultSx <- within(finalResultSx,{
  symbolJoinTest[symbolJoinTest=="*" & (PRS_value %in% c("Low-Intermediate","0","2") | 
                                          PTGENDER!="Female")] <- ""
})

# Analysis
finalResultSx$analysis <- modGrid[match(finalResultSx$variable,modGridDx$PRS),"analysis"]

finalResultSx[grep("pQTLCSF",finalResultSx$variable),"analysis"] <- "univariate.pQTL"
finalResultSx[grep("eQTLBrain",finalResultSx$variable),"analysis"] <- "univariate.eQTL"

# PTGENDER to group
names(finalResultSx)[names(finalResultSx)=="PTGENDER"] <- "Group"
finalResultSx$Group <-factor(finalResultSx$Group,levels = c("Female","Male"))


# PANEL PLOT PRIMARY ANALYSIS···············································
plotData.primary <- finalResultSx[finalResultSx$analysis=="Primary",]

# Auxiliar
# Variable and levels to factor
plotData.primary$variable <- factor(plotData.primary$variable,
                                    levels=c(riskVars,"APOE4"))
plotData.primary$PRS_value <- factor(plotData.primary$PRS_value,
                                     levels=c("Low-Intermediate","High",
                                              "0","1","2"))

plotData.primary$Volumes <- factor(plotData.primary$Volumes,levels=volVars)  

pos_dodge <- position_dodge(width=0.6) 
volume.labs <- c("Whole\nhippocampus","Subiculum",
                 "Pre-\nsubiculum","Para-\nsubiculum")
names(volume.labs) <- volVars


PRS.labs <- sapply(strsplit(c(riskVars,"APOE4"),split="_"),"[",1)
names(PRS.labs) <- c(riskVars,"APOE4")
PRS.labs[PRS.labs=="ADnoAPOE"] <- "AD\nno APOE"

#PLOT
panel <- 
  ggplot(plotData.primary,aes(x=Group)) +
  geom_hline(yintercept=0,linetype="dashed",color="red")+
  geom_pointrange(aes(y=time_visit.trend , ymin=lower.CL , ymax=upper.CL, 
                      color=as.factor(PRS_value), alpha=alpha),
                  position = pos_dodge, show.legend = F,size=.3,
                  linewidth=.5) +
  geom_text(aes(y=max(time_visit.trend)+mean(time_visit.trend)/3,
                label=symbolJoinTest))+
  facet_grid(rows=vars(variable),cols=vars(Volumes),scales = "free",
             labeller=labeller(Volumes=volume.labs, variable=PRS.labs))+
  scale_color_manual(values = c("#D95F02","#7570B3",
                                "#0033CC","#993366","#990000")) +
  scale_alpha(range = c(0.3,1))+
  coord_flip()+
  
  theme_bw() +
  theme(strip.background = element_blank(),
        plot.margin = margin(1,1,.5,1,"cm"),
        axis.title = element_blank(),
        axis.text.x = element_text(angle=90,hjust=1)) +
  ggtitle("Genetic effect over time rate of change of hippocampal volumes by sex.") +
  labs(subtitle = "Adjusted for Age at baseline, diagnostic group, intracranial volume, years of education and tesla scan parameter. <br/>
       Trends for <span style = 'color: #D95F02;'>**Low-Intermediate**</span> and <span style = 'color: #7570B3;'>**High**</span> predisposition to PRS trait or disease<br/>
       or for <span style = 'color: #0033CC;'>**0**</span>,<span style = 'color: #993366;'>**1**</span> or
       <span style = 'color: #990000;'>**2**</span> effect alleles" ,
       caption="Linear coefficient and 95% confidence intervals for volumetric rate of change over time (mm3 change per year),by sex group and level of predisposition for<br/>
       the PRS AD or APOE4 carriership. Highlighted estimates indicate significance differences between High and Low-Intermediate predispositions or number<br/>
       of effect alleles for a particular sex group. Panels with an \\* present significant difference in effect on volume change between groups. The red dashed line<br/>
       marks no time trend. Estimates correspond to models L.3") +
  theme(plot.caption = element_markdown(hjust = 0, lineheight = 1.5),
        plot.subtitle = element_markdown(size = 12, hjust = 0),
        plot.title = element_markdown(size = 16, hjust = 0))

panel.primary.Sx <- panel
panel.primary.Sx
# 9.86x7.58 i landscape


# PANEL PLOT Univariate brain ANALYSIS···············································
plotData.univar.brain <- finalResultSx[finalResultSx$analysis=="univariate.eQTL",]

# Auxiliar
# Variable and levels to factor
plotData.univar.brain$variable <- factor(plotData.univar.brain$variable,
                                         levels=snpVars[2:7])
plotData.univar.brain$PRS_value <- factor(plotData.univar.brain$PRS_value,
                                          levels=c("0","1","2"))

plotData.univar.brain$Volumes <- factor(plotData.univar.brain$Volumes,levels=volVars)  

PRS.labs <- sapply(strsplit(snpVars[2:7],split="\\."),"[",2)
names(PRS.labs) <- snpVars[2:7]


panel.univariate.brain.Sx <- 
  ggplot(plotData.univar.brain,aes(x=Group)) +
  geom_hline(yintercept=0,linetype="dashed",color="red")+
  geom_pointrange(aes(y=time_visit.trend , ymin=lower.CL , ymax=upper.CL, 
                      color=as.factor(PRS_value)),
                  position = pos_dodge, show.legend = F,size=.3,
                  linewidth=.5, alpha=0.2) +
  geom_text(aes(y=max(time_visit.trend)+mean(time_visit.trend)/3,
                label=symbolJoinTest))+
  facet_grid(rows=vars(variable),cols=vars(Volumes),scales = "free",
             labeller=labeller(Volumes=volume.labs, variable=PRS.labs))+
  scale_color_manual(values = c("#0033CC","#993366","#990000")) +
  coord_flip()+
  
  theme_bw() +
  theme(strip.background = element_blank(),
        plot.margin = margin(1,1,.5,1,"cm"),
        axis.title = element_blank(),
        axis.text.x = element_text(angle=90,hjust=1)) +
  ggtitle("eQTL Brain individual SNPs effect over time rate of change of<br>
          hippocampal volumes by sex.") +
  labs(subtitle = "Adjusted for Age at baseline, diagnostic group, intracranial volume, years of education<br>
  and tesla scan parameter. <br/>
       Trends for <span style = 'color: #0033CC;'>**0**</span>,<span style = 'color: #993366;'>**1**</span> or
       <span style = 'color: #990000;'>**2**</span> effect alleles" ,
       caption="Linear coefficient and 95% confidence intervals for volumetric rate of change over time (mm3 change per year),by<br>
       diagnostic group and level of predisposition for the PRS trait/disease. Highlighted estimates indicate significance<br>
       differences between High and Low-Intermediate predispositions for a particular sex group. Panels with an<br/>
       \\* present significant difference in PRS effect on volume change between groups. The red dashed line marks no<br/>
       time trend. Estimates correspond to models L.3.") +
  theme(plot.caption = element_markdown(hjust = 0, lineheight = 1.5),
        plot.subtitle = element_markdown(size = 12, hjust = 0),
        plot.title = element_markdown(size = 16, hjust = 0))

panel.univariate.brain.Sx

# 9.86x7.58 portrati

# PANEL PLOT Univariate brain ANALYSIS···············································
plotData.univar.CSF <- finalResultSx[finalResultSx$analysis=="univariate.pQTL",]

# Auxiliar
# Variable and levels to factor
plotData.univar.CSF$variable <- factor(plotData.univar.CSF$variable,
                                       levels=snpVars[8:11])
plotData.univar.CSF$PRS_value <- factor(plotData.univar.CSF$PRS_value,
                                        levels=c("0","1","2","1-2"))

plotData.univar.CSF$Volumes <- factor(plotData.univar.CSF$Volumes,levels=volVars)  

PRS.labs <- sapply(strsplit(snpVars[8:11],split="\\."),"[",2)
names(PRS.labs) <- snpVars[8:11]

# aux <- with(plotData.univar.CSF,
#             ident %in% c("pQTLCSF.rs429358_C|presubiculum",
#                          "pQTLCSF.rs75627662_T|subiculum",
#                          "pQTLCSF.rs429358_C|Whole_hippocampus"))
# plotData.univar.CSF[aux,]

panel.univariate.CSF.Sx <- 
  panel %+% plotData.univar.CSF +
  scale_color_manual(values = c("#0033CC","#993366","#990000","#990000")) +
  facet_grid(rows=vars(variable),cols=vars(Volumes),scales = "free",
             labeller=labeller(Volumes=volume.labs, variable=PRS.labs)) +
  ggtitle("pQTL CSF individual SNPs effect over time rate of change of hippocampal volumes by sex.") +

  labs(subtitle = "Adjusted for Age at baseline, diagnostic group, intracranial volume, years of education and tesla scan parameter. <br/>
       Trends for <span style = 'color: #0033CC;'>**0**</span>,<span style = 'color: #993366;'>**1**</span> or
       <span style = 'color: #990000;'>**2**</span> effect alleles",
       caption="Linear coefficient and 95% confidence intervals for volumetric rate of change over time (mm3 change per year),by diagnostic group and level of<br>
       predisposition for the PRS trait/disease. Highlighted estimates indicate significance differences between High and Low-Intermediate predispositions<br>
       for a particular diagnostic group. Panels with an \\* present significant difference in PRS effect on volume change between groups. The red dashed line<br>
       marks no time trend. Estimates correspond to models L.3")

panel.univariate.CSF.Sx

# 9.86x7.58 landscape

# EXPORT EXCEL TIME TRENDS MODELS L2 i L3

# Get table for parameter estimates

timeTrendList <- list(finalResult,finalResultSx)
names(timeTrendList) <- paste0("Models L",2:3)
timeTrendList <- lapply(timeTrendList,function(x){
  res <- x
  res$model <- with(x,paste(Volumes,variable,sep=":"))
  res <- res[c("model",names(x)[c(1:9,11,12)])]
  return(res)
})

# Build Excel worksheet
wb <- createWorkbook()
invisible(lapply(names(timeTrendList),function(x)addWorksheet(wb,x)))
invisible(
  lapply(1:length(timeTrendList),
         function(i) writeDataTable(wb = wb,
                                    x = timeTrendList[[i]],
                                    sheet = names(timeTrendList)[i],
                                    withFilter = T,
                                    tableStyle = "TableStyleLight1")
  )
)

path <- here("results",paramT,"2_Modelling","2_longitudinalTrajectories",
             "taules","longitudinalModels_timeTrends.xlsx")

saveWorkbook(wb,file=path,overwrite = T)


# EXPROT LONGITUDINAL MODEL PARAMETERS

  # Get table for parameter estimates

  modelParams <- lapply(c("modGlobal","mod_3wayIntDX","mod_3wayIntSx"),
                        FUN = ensambleParamsTable, data=result)
  
  names(modelParams) <- paste0("Models L",1:3)
  
  
  # Build Excel worksheet
  wb <- createWorkbook()
  invisible(lapply(names(modelParams),function(x)addWorksheet(wb,x)))
  invisible(
    lapply(1:length(modelParams),
           function(i) writeDataTable(wb = wb,
                                 x = modelParams[[i]],
                                 sheet = names(modelParams)[i],
                                 withFilter = T,
                                 tableStyle = "TableStyleLight1")
    )
  )

  
path <- here("results",paramT,"2_Modelling","2_longitudinalTrajectories",
             "taules","longitudinalModels_parameters.xlsx")

saveWorkbook(wb,file=path,overwrite = T)
```
