## Supplementary material for "Genetic drivers of hippocampal atrophy highlight the role of *APOE* functional variants and AD polygenicity in Mild Cognitive Impairment": Supplemental methods.docx

**SUPPLEMENTARY METHODS**

***Modelling of Hippocampal trajectories***

We analyzed the association genetic predisposition to AD and changes in hippocampal volumes repeatedly assessed over a 5-years period using linear mixed-effect models with random intercept for individuals:

$Y_{ij}^{\left( h \right)}=\beta_{0}+\beta_{1}t_{ij}+\beta_{2}PRS_{i}^{\left( g \right)}+\beta_{3}DX_{i}+\beta_{4}t_{ij}*DX_{i}+\beta_{5}t_{ij}*PRS_{i}^{\left( g \right)}+\sum_{k=6}^{9} \beta_{k}X_{ki}+b_{1i}+b_{2i}t_{ij}+\epsilon_{ij} \left( Eq.1 \right)$

where $Y_{ij}$ corresponds to hippocampal volume measured in participant *i* at visit number *j*; $PRS_{i}^{\left( g \right)}$ refers to the degree of predisposition of the participant for the trait or disease; $b_{1i}$ represents the random intercept for participant *i*, assumed to be normally distributed with mean 0 and variance $\sigma_{1}^{2}$; $b_{2i}$ corresponds to the random slope of time for participant *i*, assumed to be normally distributed with mean 0 and variance $\sigma_{2}^{2}$; i $\epsilon_{ij}$ denotes the residuals of the model, assuming a normal distribution with mean 0 and variance $\sigma_{e}^{2}$. The variable $t_{ij}$ represents the time in years elapsed between the baseline visit (*j*=0) and visit *j*, and can be considered as the participant's age at visit j centered around the age at the baseline visit. Finally, $\sum_{k=6}^{9} \beta_{k}X_{ki}$, corresponds to the model terms for the covariates. Disease status-specific trajectories were also assessed by including a three-way interaction term in the model:

$Y_{ij}=\beta_{0}+\beta_{1}t_{ij}+\beta_{2}PRS_{i}^{\left( g \right)}+\beta_{3}DX_{i}+\beta_{4}t_{ij}*DX_{i}+\beta_{5}t_{ij}*PRS_{i}^{\left( g \right)}+\beta_{6}DX_{i}*PRS_{i}^{\left( g \right)}+\beta_{7}t_{ij}*DX_{i}*PRS_{i}\left( g \right)$

$+\sum_{k=6}^{8} \beta_{k}X_{ki}+b_{1i}+b_{2i}t_{ij}+\epsilon_{ij} \left( Eq.2 \right)$

where the annual average volume variation was calculated according to the diagnostic status (CU, MCI, AD) and the genetic predisposition to the trait or disease (Low-Intermediate/High):

${\Delta Vol}_{CU|Low-Int}=\beta_{1}$	${\Delta Vol}_{CU|High}=\beta_{1}+\beta_{5:High}$

${\Delta Vol}_{MCI|Low-Int}=\beta_{1}+\beta_{4:MCI}$	${\Delta Vol}_{MCI|High}=\beta_{1}+\beta_{5:High}+\beta_{4:MCI}+\beta_{7:MCI:High}$

${\Delta Vol}_{AD|Low-Int}=\beta_{1}+\beta_{4:AD}$	${\Delta Vol}_{MCI|High}=\beta_{1}+\beta_{5:High}+\beta_{4:AD}+\beta_{7:AD:High}$
