## Supplementary figures and images for "Genetic drivers of hippocampal atrophy highlight the role of *APOE* functional variants and AD polygenicity in Mild Cognitive Impairment"

### SuplFig1.pdf

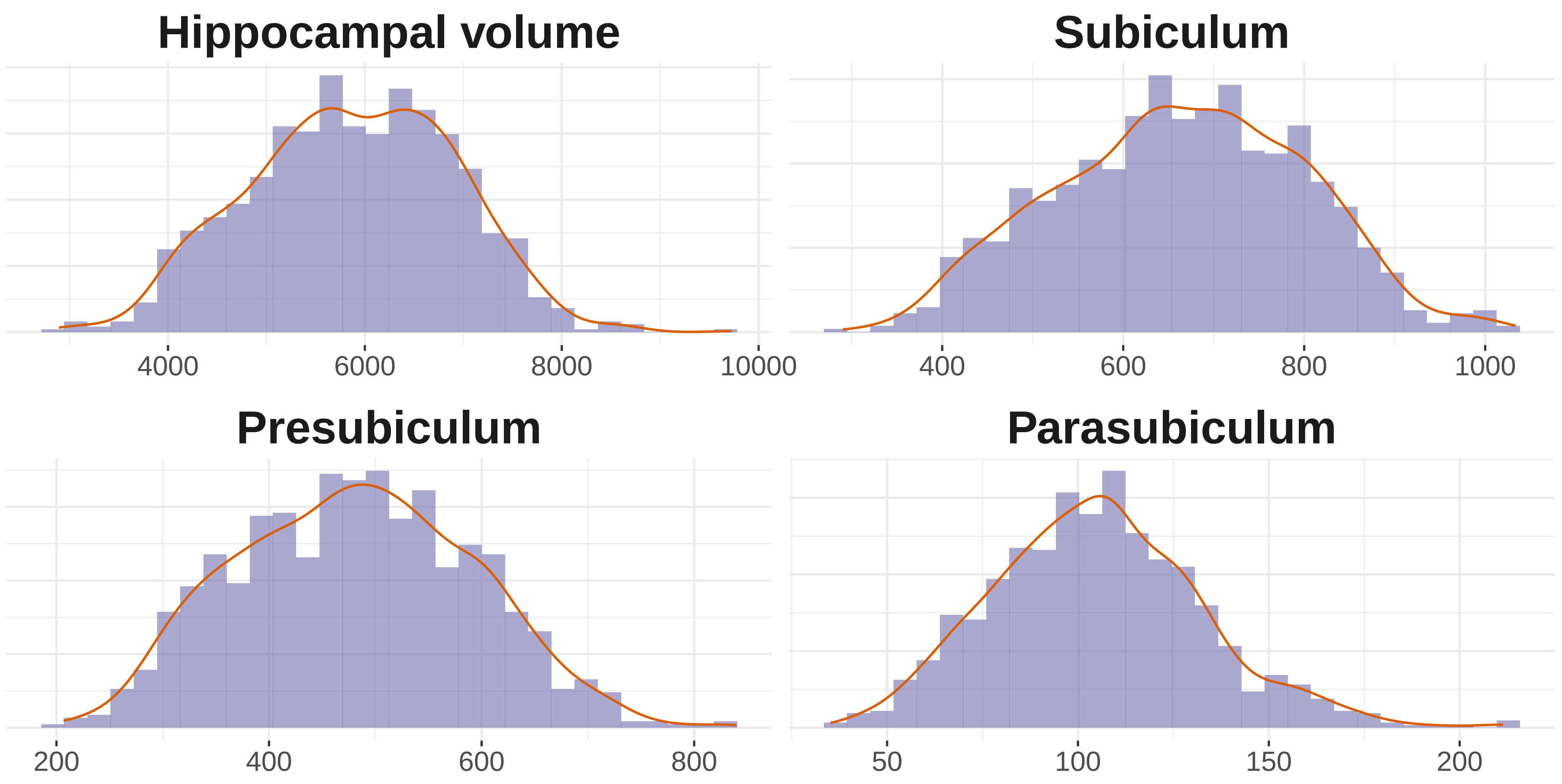

### SuplFig2.pdf

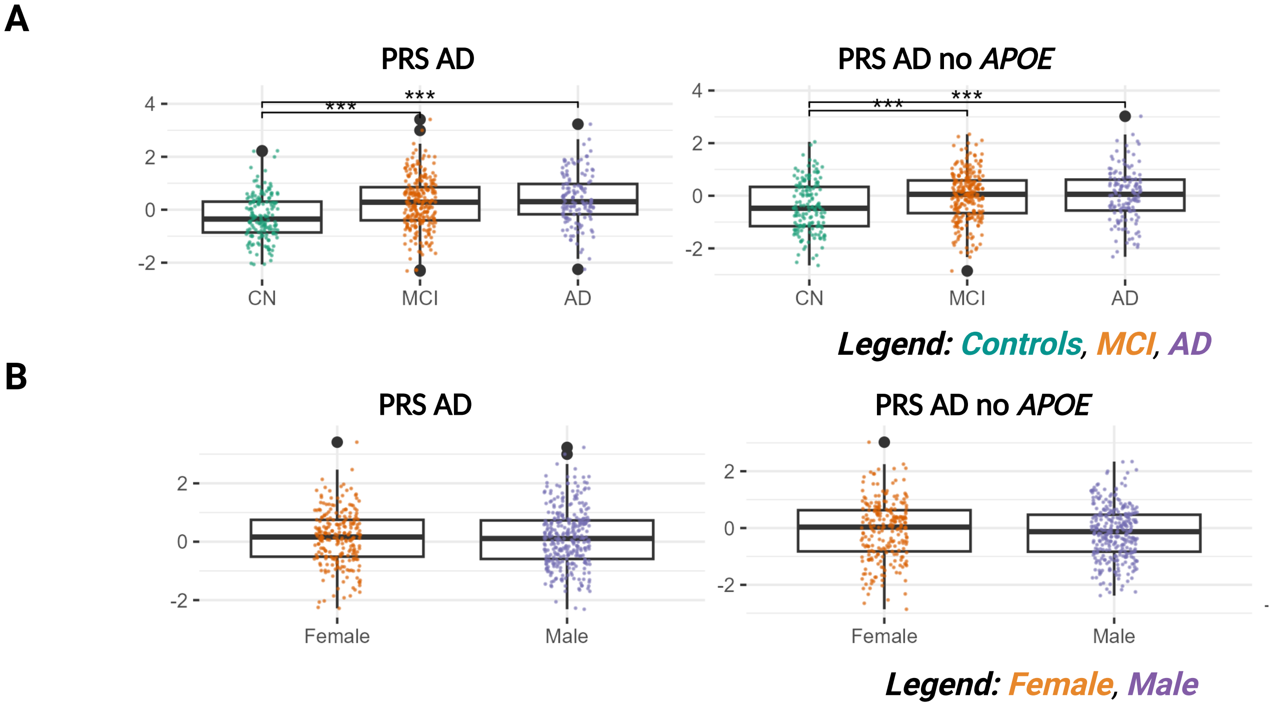

### SuplFig3.pdf

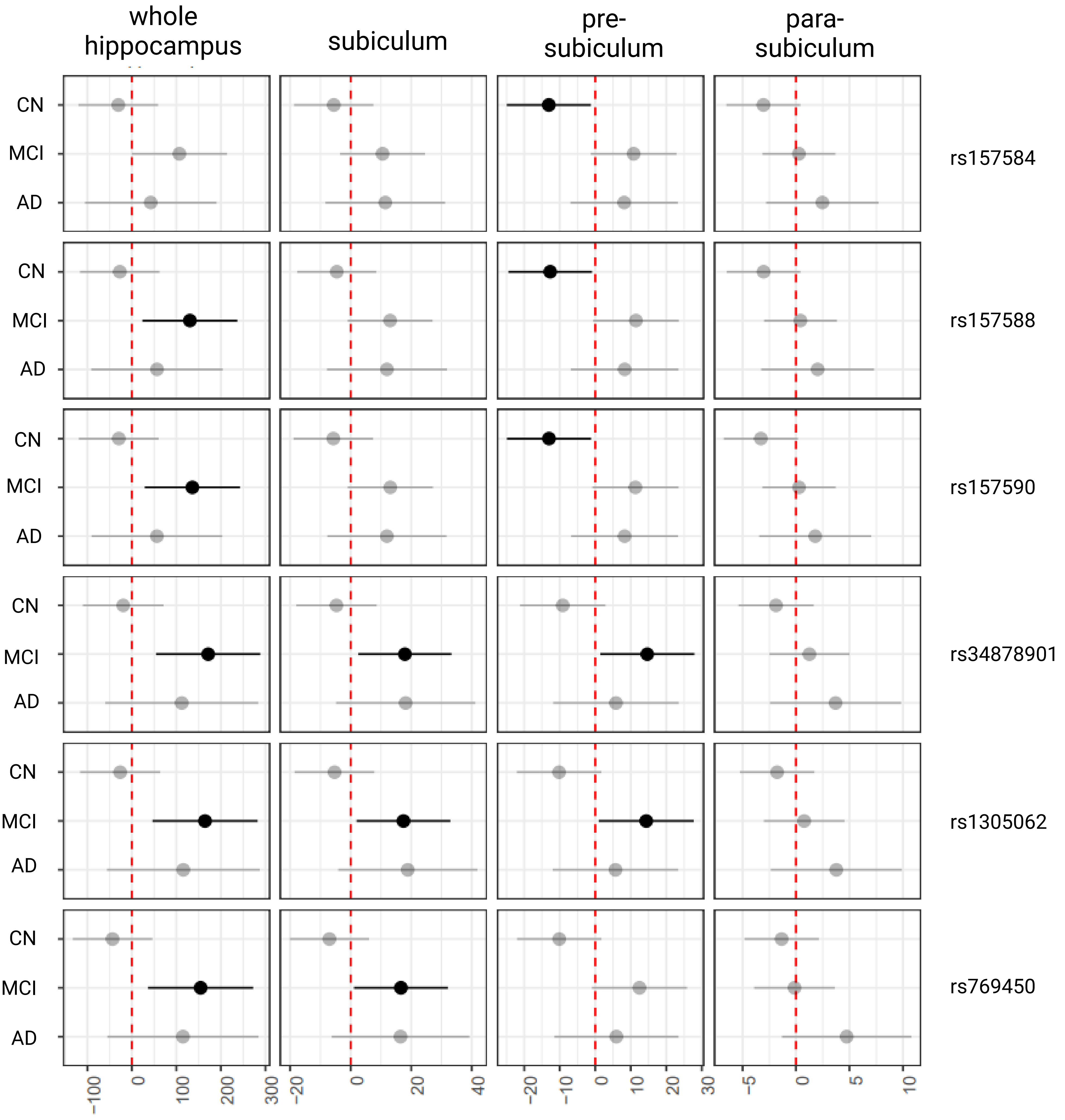


*Legend: Trends for **0**, **1** or **2** counts of effect alleles.*

### SuplFig4.pdf

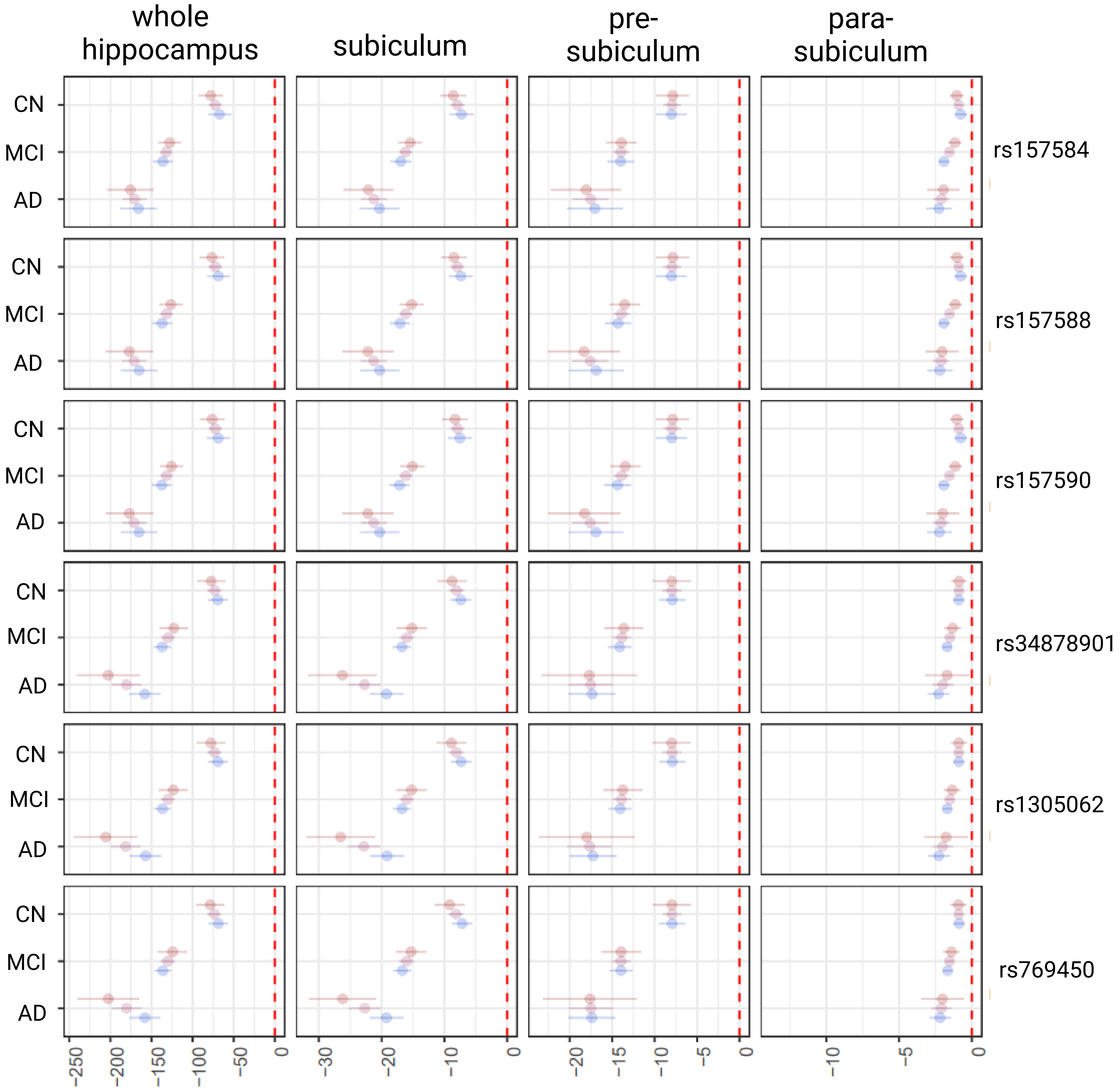


*Legend: Trends for* ***0***, ***1*** *or* ***2*** *counts of effect alleles.*

### SuplFig5.pdf

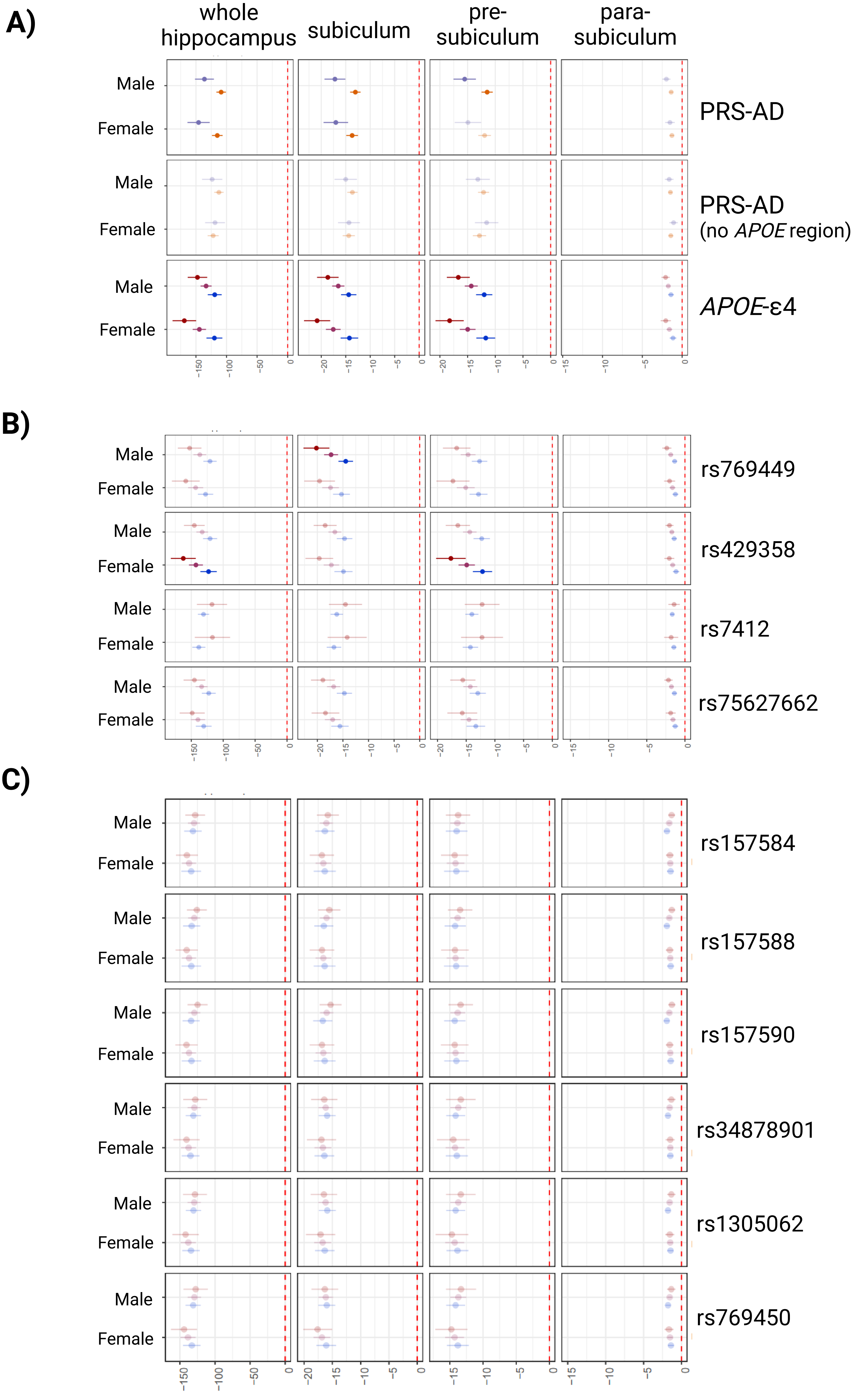


*Legend:* ***Global****,* ***Low genetic risk****,* ***High genetic risk*** ***or trends for 0,1,2 effect alleles***
